# Characterizing the genetic basis of Cardio-Renal-Metabolic multimorbidity using multivariate genomic modelling

**DOI:** 10.64898/2026.06.16.26355643

**Authors:** Abhiram D. B., Vignesh Arunachalam, Rodney A Lea, Kim Ngan Tran, Rohit Mishra, Prasad Challa, Sree K Venuthurupalli, Samuel Sidharta, Shivashankar H. Nagaraj

## Abstract

Cardio-renal-metabolic multimorbidity (CRMM) denotes co-occurring cardiovascular, kidney, and metabolic diseases. The shared genetic architecture of CRMM across diverse ancestries remains incompletely understood. Using ancestry-specific GWAS-summary statistics from >3.5 million participants across seven biobanks, we applied genomic structural equation modelling to characterise shared genetic architecture of hypertension, heart failure, ischaemic heart disease, chronic kidney disease, and type 2 diabetes in European (EUR), East Asian (EAS), and African (AFR) populations. Conditional analysis of the hierarchical latent-factor model identified 148, 16, and 138 independent CRMM-associated loci in EUR, AFR, and EAS populations, respectively. Integrative analyses implicated neurological pathways and brain-tissue enrichment, nominating *ATXN2L* as a putative functional gene. Drug repurposing analyses highlighted existing diabetes and hypertension therapies as potential therapeutic candidates, while Mendelian randomisation identified adiposity, cholesterol, and other traits as candidate causal risk factors for CRMM. These findings delineate cross-ancestry and ancestry-specific CRMM genetics, informing therapeutic targets and precision medicine.

## Introduction

Cardiovascular disease (CVD), chronic kidney disease (CKD), and type 2 diabetes (T2D) form a triad of interconnected conditions that rarely occur in isolation. Their co-occurrence, cardio-renal-metabolic multimorbidity (CRMM), represents one of the most formidable global health challenges in the twenty-first century.^1^ All three diseases share pathophysiological pathways and amplify each other’s progression: T2D is a leading cause of CKD and is associated with a two- to four-fold increase in CVD risk, while CKD accelerates heart failure (HF) and risk for cardiovascular events and death.^2,3^ Together, the seven key components of CRMM accounted for more than 600 million disability-adjusted life years worldwide in 2021, highlighting their substantial contribution to the global burden of disease.^4^

Genome-wide association studies (GWAS) have identified hundreds of loci associated independently with cardiometabolic conditions, including approximately 241 for coronary artery disease,^5^ 1,026 for CKD,^6^ and 1,289 for T2D.^7^ Despite the well-established clinical co-occurrence and temporal progression of these disorders, the shared genetic architecture underlying their overlap remains incompletely characterized, particularly in non-European populations. A major limitation of existing GWAS is their strong European ancestry (EUR) bias, limiting the generalizability of findings to other populations.^8^ The availability of large-scale, deeply phenotyped biobanks now offer an unprecedented opportunity to investigate the shared genetic architecture of CRMM across ancestrally diverse populations.

CVD, CKD, and T2D exhibit strong bidirectional and causal relationships,^9–11^ frequently co-occurring and progressing sequentially over a patient’s life. Observational studies have identified multiple shared clinical risk factors associated with CRMM, including age,^12^ sex,^13^ family history,^14^ body mass index,^15^ alcohol consumption, smoking,^16^ and psychosocial stress.^17^ However, very few studies have systematically examined which of these factors exert causal effects on CRMM outcomes. Moreover, methodological challenges, such as jointly modelling multiple correlated conditions, investigating genetic and clinical determinants, and accounting for population diversity in multi-ancestry settings, have limited comprehensive investigation of CRMM etiology.

To date, few GWASs have examined the genetic architecture of CRMM, where existing attempts have used two distinct approaches. Two studies modelled a latent CRMM factor derived from six cardiometabolic traits, identifying 2,212 genome-wide significant variants and 1,599 risk loci, respectively.^18,19^ A third study used biomarker-based topic modelling to identify five candidate CRMM subtypes and 57 genome-wide significant, predominantly subtype-specific variants in ∼52,700 individuals.^20^ However, these studies have three key limitations. First, the two latent-factor studies fitted a single common factor without testing factorability or comparing alternative factor structures, so the validity of the genetic construct remains unclear.^18,19^ Second, the genetic analyses were based mainly or entirely on individuals of European ancestry, limiting transferability to other populations. Third, the reported associations have not been independently replicated or validated across ancestries, although polygenic scores for three of the five candidate subtypes were associated with CRMM-related diagnoses in UK Biobank.^20^ Together, these shortcomings underscore the need for rigorous, independently replicated, multi-ancestry analyses that define and validate the genetic structure of CRMM.

To address outstanding questions regarding the shared genetic architecture of CRMM, its generalisability across diverse ancestries, and the lack of large-scale replication of multivariate findings, we leveraged genomic data from seven population biobanks: FinnGen,^21^ UK Biobank (UKB),^22^ the Million Veteran Program (MVP),^23^ China Kadoorie Biobank (CKB),^24^ the Taiwan Precision Medicine Initiative (TPMI),^25^ BioBank Japan (BBJ),^26^ and All of Us (AoU),^27^ spanning four ancestry groups (EUR, African [AFR], Admixed American [AMR], and East Asian [EAS]). Our primary aims were to characterise the genetic architecture of CRMM across ancestries, evaluate whether a shared latent genetic factor underlies its component diseases, and identify genetic variants contributing to multimorbidity risk. To achieve this, we applied genomic structural equation modelling (Genomic SEM) to construct a latent CRMM factor from hypertension (HP), ischaemic heart disease (IHD), HF, CKD, and T2D, followed by multivariate genome-wide association analyses. We subsequently performed functional and gene-based analyses, drug repurposing analyses, replication studies, ancestry-specific polygenic risk score (PRS) evaluations, and two-sample Mendelian randomization (2SMR) analyses to investigate the biological and clinical implications of identified genetic associations.

## Results

### Genetic overlap in CRMM

We first meta-analysed GWAS summary statistics for each of the five CRMM components (HP, HF, IHD, T2D, and CKD) across the contributing biobanks (FinnGen, UKB, MVP, CKB, TPMI, BBJ, and AoU), separately within each ancestry group. The ancestry-specific meta-analyses yielded effective sample sizes across the five CRMM components ranging from 323,911 to 1,132,967 in the European (EUR) population, 62,937 to 95,582 in the African (AFR) population, and 78,519 to 347,774 in the East Asian (EAS) population after quality control measures (Methods). For the American (AMR) population, where only a single cohort (AoU) was available, effective sample sizes ranged from 30,385 to 31,957 (Supplementary Tables 1–3). Bivariate LD score regression intercepts indicated minimal sample overlap between contributing cohorts, with all pairwise intercept estimates <0.2 (Supplementary Table 4).

To characterise the shared genetic architecture of CRMM components across ancestries, we evaluated pairwise genetic correlations between the five traits. Profound genetic correlations (r_g_ = 0.44–0.87) were observed among HP, HF, and IHD across EUR, AFR, and EAS ancestries. CKD was also strongly correlated with HP (r_g_ = 0.56–0.91 across ancestries), and CKD and T2D were significantly correlated in EUR and EAS populations (r_g_ > 0.5), whereas the correlation in AFR was lower (r_g_ = 0.32), which suggests that the genetic overlap between CKD and T2D may differ by ancestry (Fig. 1A–C and Supplementary Table 5). Factorability assessment supported downstream factor analysis in EUR and AFR populations, with Kaiser-Meyer-Olkin (KMO) Measure of Sampling Adequacy (MSA)^28^ values of 0.84 and 0.77, respectively, and significant Bartlett’s tests (*P*<0.05),^29^ confirming factorability. Parallel analyses^30^ suggested two, three, and two latent factors in EUR, AFR, and EAS ancestries, respectively (Supplementary Table 6; Supplementary Fig. 2). Comparison of competing exploratory factor analysis (EFA) solutions supported a parsimonious two-factor model, which explained 65%, 62%, and 72% of the variance in EUR, AFR, and EAS cohorts, respectively (Supplementary Table 7). Furthermore, hierarchical clustering supported this structure, grouping HP, HF, and IHD into a CVD latent factor, representing a shared genetic liability, which together with T2D and CKD formed a higher-order CRMM latent factor (Fig. 1D–F).

**Fig. 1.**
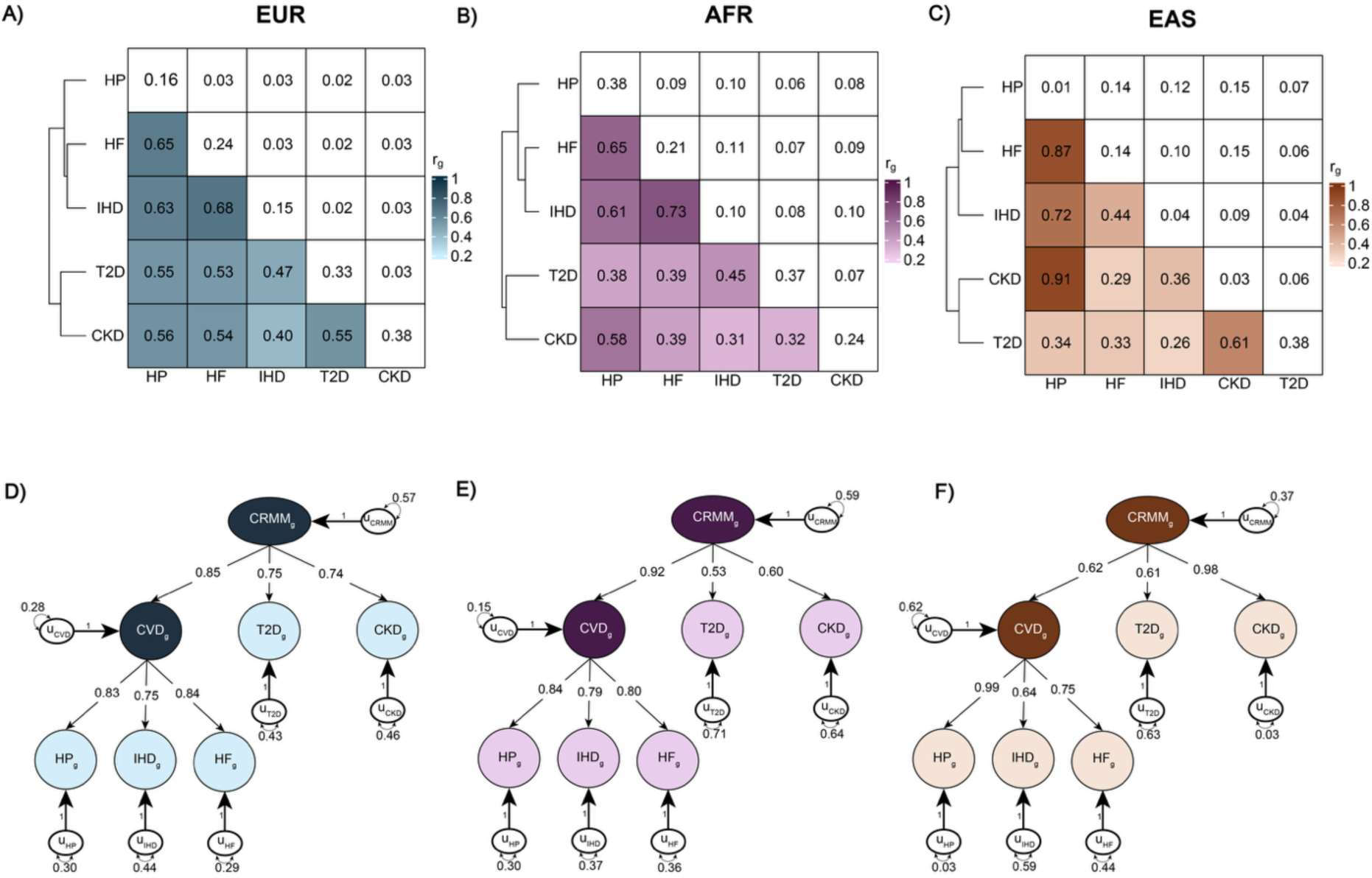
Genetic correlations among CRMM components and the derived CRMM latent factor. **A–C**, SNP-based heritability and pairwise genetic correlations among the five CRMM components estimated using LD score regression. For each ancestry (EUR, AFR, and EAS, shown in that order), the lower-left triangle displays pairwise genetic correlations, the upper-right triangle shows the corresponding standard errors, and the diagonal elements represent SNP-based heritability estimates. **D–F**, Path diagram of the hierarchical model estimated using Genomic SEM, with standardized factor loadings. EUR: European; AFR: African; EAS: East Asian; HP: hypertension; HF: heart failure; IHD: ischemic heart disease; T2D: type 2 diabetes; CKD: chronic kidney disease; CRMM: cardio-renal-metabolic multimorbidity; *u*: residual variance not explained by the latent factor; subscript g: indicates that the model is based on genetic covariances.

To confirm the latent CRMM structure identified by exploratory factor analysis (EFA), we performed a confirmatory factor analysis (CFA). A single common factor model met the fit criteria in EUR and AFR but not in EAS (Supplementary Table 8). A correlated second-order hierarchical model met the fit criteria in all three ancestry groups and consistently outperformed the single-factor model in EUR, AFR, and EAS (Supplementary Tables 9). Model fit was good in EUR (*χ*^2^ = 97.60, P = 3.18 × 10^-20^; Akaike information criterion (AIC) = 119.60; comparative fit index (CFI) = 0.98; standardized root mean square residual (SRMR) = 0.028), good fit in AFR (*χ*^2^ = 27.04, P = 1.95 × 10^-5^; AIC = 49.04; CFI = 0.96; SRMR = 0.059), and adequate fit in EAS (*χ*^2^ = 7.11, P = 0.13; AIC = 29.11; CFI = 0.99; SRMR = 0.062). These findings support a hierarchical, rather than a common, genetic architecture for CRMM in which correlated lower-order traits define a higher-order latent CRMM factor (Fig. 1D–F).

### Ancestry-specific pleiotropic loci

We performed ancestry-specific multivariate GWAS of the CRMM latent factor using Genomic SEM across EUR, AFR, and EAS populations to identify the SNPs responsible for the CRMM latent factor (Fig. 2A–C; Supplementary Fig. 3). A total of 8,509,536 SNPs in EUR, 14,602,226 SNPs in AFR, and 6,997,191 SNPs in EAS were retained for multivariate GWAS following quality control (QC). SNPs exhibiting significant Q_SNP_ heterogeneity (indicating effects that were trait-specific rather than mediated through the latent CRMM factor) were excluded (see Methods), resulting in 8,405,257 SNPs in EUR, 14,600,283 SNPs in AFR, and 6,991,910 SNPs in EAS for post-GWAS analyses. The effective sample size (N_eff_) of the CRMM latent factor GWAS was estimated at 353,130 for EUR, 75,436 for AFR, and 164,373 for EAS, with corresponding SNP-based heritability estimates (h^2^_SNP_) of 0.18, 0.23, and 0.12, respectively (Supplementary Table 10).

**Fig. 2.**
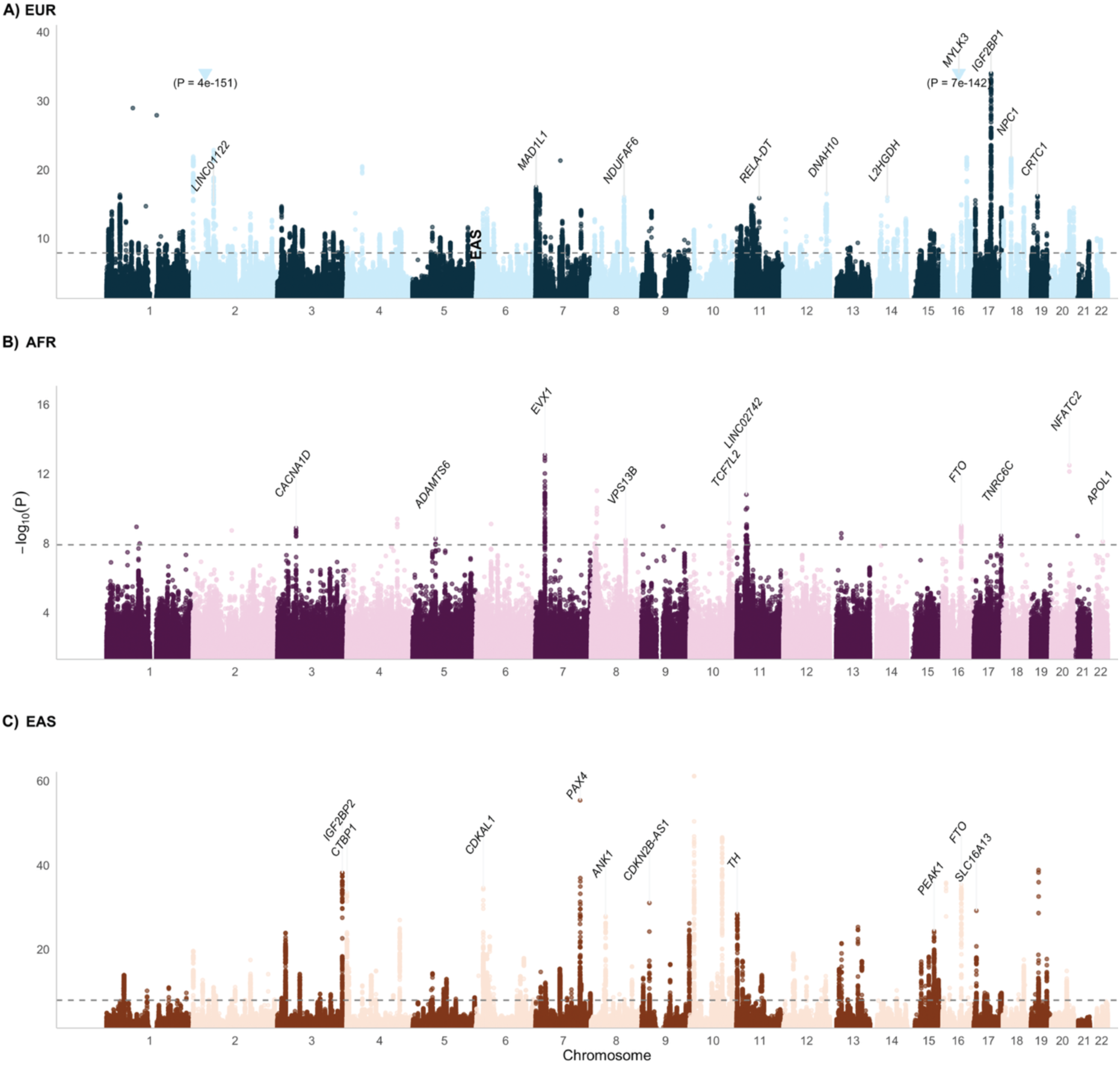
Manhattan plots of genome-wide significant loci for CRMM across ancestries. Manhattan plots showing genome-wide association results for CRMM in European (EUR), African (AFR), and East Asian (EAS) ancestries (top to bottom). The horizontal dashed line indicates the genome-wide significance threshold (*P* = 1.25 × 10^−8^). The top 10 lead SNPs are highlighted and annotated with their corresponding mapped genes.

We identified 11,484, 165, and 7,732 genome-wide significant (GWS; *P* < 1.25 × 10^-8^) SNPs associated with CRMM in EUR, AFR, and EAS populations, respectively. Cross-ancestry comparison revealed 719 shared genome-wide significant (GWS) SNPs between EUR and EAS and 24 between AFR and EAS, whereas no GWS SNPs were shared across all three ancestry groups. LD-based clumping (see Methods) identified 287, 30, and 202 lead SNPs in EUR, AFR, and EAS, respectively. Among these, 39, 2, and 27 overlapped with previously reported CRMM loci from Lu, et al. ^18^. Furthermore, four EUR and three EAS GWS SNPs replicated associations reported by Wang, et al. ^19^, and one EUR GWS SNP overlapped with a locus identified by Donker, et al. ^20^.

To identify conditionally independent signals beyond the lead SNPs, we performed conditional and joint (COJO) analysis. COJO analysis identified 148 independent SNPs in EUR, 16 in AFR, and 139 in EAS. Cross-ancestry heterogeneity analysis further showed that 77 of the EUR COJO SNPs and 20 of the EAS COJO SNPs did not exhibit statistically significant heterogeneity across EUR, AFR, and EAS ancestries (BH-adjusted (*P* > 0.05)), suggesting that their effect estimates were not significantly different across the three ancestries. Gene mapping using Ensembl Variant Effect Predictor (VEP) mapped these COJO SNPs to 113 genes in EUR, 11 genes in AFR, and 91 genes in EAS (Supplementary Fig. 4). Among the COJO SNPs we identified three SNPs shared between EUR and EAS populations (rs80196932 [*NUS1*], rs1426371 [*WSCD2*], and rs2925979 [*CMIP*]), with concordant directions of effect across the two populations. Functional annotation identified six missense variants in EUR and ten in EAS. Of these, five SNPs (three in EUR, one in AFR and one in EAS) demonstrated a RegulomeDB score of 1a, indicating strong evidence of regulatory activity (Supplementary Fig. 5). These variants mapped to five genes, including *HSF2BP* (rs2051407), *COMT* (rs165722), and *HTT* (rs362270) in EUR, *ZNF318* (rs1214761) in AFR, and *H3-3B* (rs1060113) in EAS. A total of 58 EUR, 13 AFR and 54 EAS COJO SNPs were previously unreported in the NHGRI-EBI GWAS Catalog v1.0 (Supplementary Tables 11–13).

We then quantified the similarity of the genetic architecture of CRMM across ancestry groups by estimating cross-ancestry genetic-effect correlations using Popcorn.^31^ We observed high genetic-effect correlations between EUR and AFR (ρ_ge_=0.97, S.E. = 0.10) and between AFR and EAS (ρ_ge_=0.82, S.E. = 0.12). The EUR-EAS comparison also showed strong concordance (ρ_ge_=1); however, Popcorn was unable to estimate a corresponding standard error, likely due to numerical instability during model fitting (Supplementary Table 14).

### Functional annotation and gene prioritization

We performed a series of post-GWAS analyses to investigate the biological mechanisms underlying the CRMM latent factor. Specifically, we conducted gene-based association analyses using Multi-marker Analysis of GenoMic Annotation (MAGMA) to identify genes associated with CRMM across the three ancestral populations. MAGMA gene-based analysis identified 497 significant genes in the EUR population (*P_Bonferroni_* < 0.05), compared with 23 and 153 significant genes in the AFR and EAS populations, respectively (Supplementary Tables 15–17). Notably, *TCF7L2* and *BDNF* demonstrated significant associations across all three ancestries.

To gain insight into the tissue-specific processes underlying CRMM susceptibility, we performed MAGMA^32^ gene-property analysis to assess whether genes associated with CRMM showed preferential expression across a range of human tissues. We tested 30 broad and 54 detailed GTEx tissue categories^33^ for enrichment of CRMM-associated genes. Among the broad tissue types, four showed significant (*P* < 1.67 × 10^-3^; Bonferroni-corrected threshold) enrichment in the EUR population, including blood vessel, brain, pituitary, and uterine tissues (Fig. 3A and Supplementary Tables 18). Analysis of the detailed tissue categories identified significant enrichment in the cerebellum, cerebellar hemisphere, pituitary, and uterus in EUR individuals (*P* < 9.26 × 10^-4^; Bonferroni-corrected threshold), while enrichment in EAS was observed for the cerebellum and cerebellar hemisphere (Supplementary Tables 19).

**Fig. 3.**
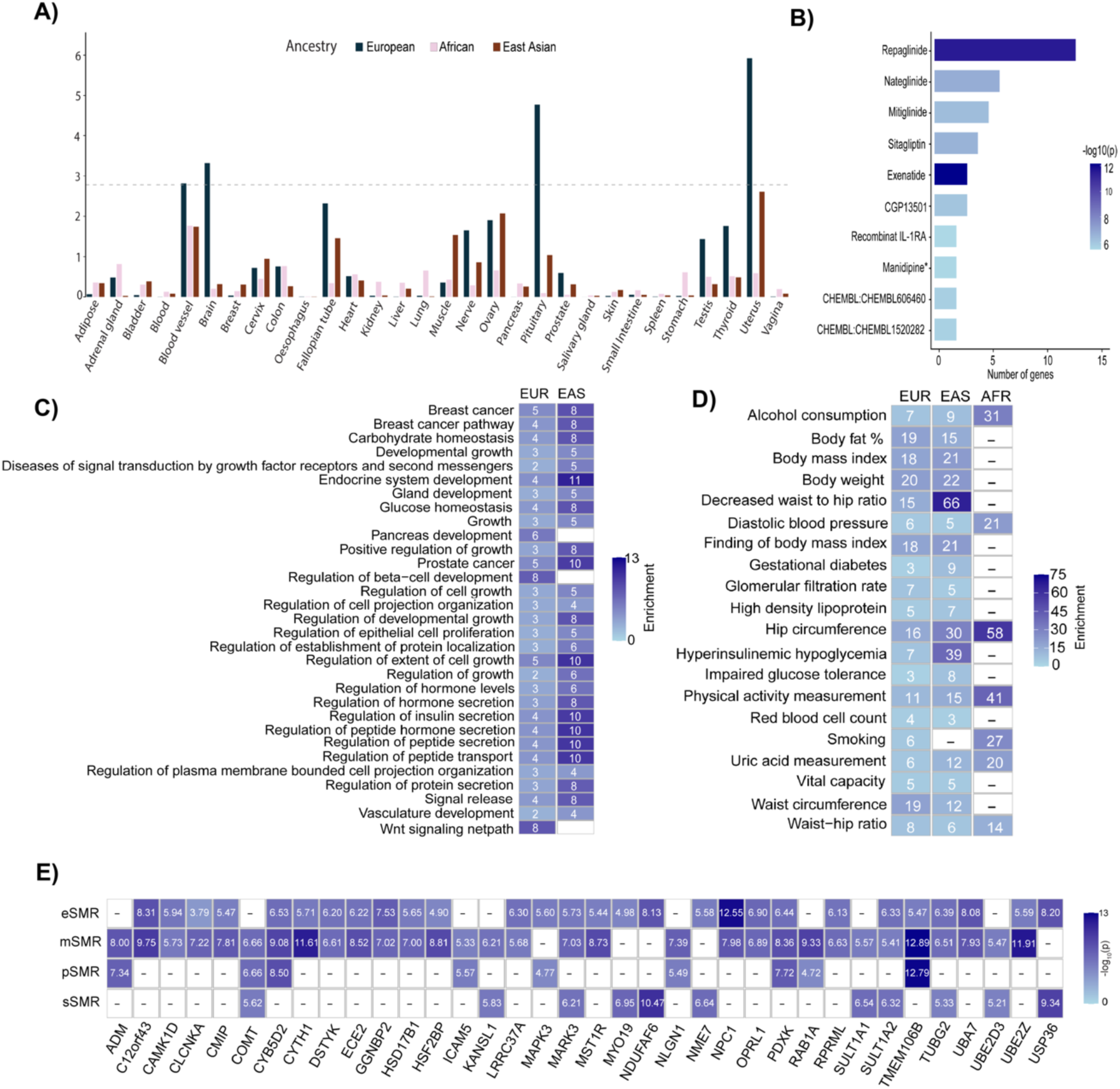
Functional enrichment and gene property analyses of CRMM across ancestries. A, Multiple bar plot showing gene-property analysis results from MAGMA across EUR, AFR, and EAS ancestries. The x-axis represents the 30 general tissue categories, and the y-axis represents the corresponding gene-property P values. Bars are colour-coded by ancestry. The horizontal dotted line indicates the Bonferroni-corrected significance threshold (*P_Bonferroni_*<0.05). Tissues exceeding the significance threshold are marked with an asterisk (*). **B,** Bar plot showing the top 10 statistically significant drugs identified from the drug-repurposing analysis (*P_Bonferroni_*<0.05).The x-axis represents the number of genes involved in each gene-drug interaction, and colour indicates the observed P value. An asterisk (*) denotes a drug identified in the AFR ancestry; the remaining drugs were identified in EAS. **C,** Biological pathway enrichment analysis of CRMM using Metascape, presented as a heat map. Pathways that remained significant after FDR correction and were common to at least two ancestries are shown. **D,** Disease pathway enrichment analysis of CRMM using Metascape, presented as a heat map. The top 20 significant pathways (P_FDR_ < 0.05) are shown and were common to at least two ancestries are shown. **E,** Heat map showing genes identified through SMR analysis in the EUR ancestry across eQTL, mQTL, pQTL, and sQTL datasets. The x-axis represents genes, and the y-axis represents the molecular QTL databases. Colour intensity indicates statistical significance on a -log_10_(P) scale. Only genes showing significant associations in at least two molecular QTL datasets are shown.

We further investigated the biological pathways underlying the gene-based associations using two complementary enrichment approaches. First, we performed gene-set enrichment analysis using the Molecular Signatures Database (MSigDB),^34^ which evaluates enrichment of genome-wide gene-level association statistics against predefined biological gene sets. This analysis identified significant enrichment of one, three, and six curated pathways in EUR, AFR, and EAS, respectively. The enriched gene sets highlighted processes related to oligodendrocyte differentiation (EUR; MSigDB: M2369), neuroinflammation (AFR; MSigDB: M39895), fibroblast growth factor receptor signalling (AFR; MSigDB: M17776), insulin receptor substrate activation (EAS; MSigDB: M27707), and maturity-onset diabetes of the young (EAS; MSigDB: M18312). GO analysis additionaly supported enrichment of regulation of developmental growth (EUR; MSigDB: M10722), regulation of cell differentiation (EUR; MSigDB: M4547), and regulation of RNA metabolic process (EUR; MSigDB: M29344), carbohydrate homeostasis (EAS; MSigDB: M14445) and intracellular glucose homeostasis (EAS; MSigDB: M14255; Supplementary Tables 20–21).

Second, we applied Metascape^35^ to the significant genes (*P_Bonferroni_* < 0.05) identified through MAGMA’s SNP wise mean model gene-based analysis (497 in EUR, 23 in AFR, and 153 in EAS). Unlike the MSigDB analysis, which evaluates enrichment across predefined gene sets using genome-wide gene-level association statistics, Metascape provides broader functional annotation of the prioritized genes across multiple biological and disease-related resources. Metascape identified 224 enriched biological processes in EUR, 7 in AFR, and 82 in EAS (Fig. 3C). Among the most prominently enriched pathways were positive regulation of mesenchymal stem cell differentiation (GO:2000741), embryonic skeletal system development (GO:0048706), and the 6q16 copy-number variation pathway (WP5400). The 6q16 copy-number variation pathway was primarily driven by *SIM1*, a transcription factor involved in neurogenesis and leptin-melanocortin signalling. (Supplementary Tables 22–23).

Metascape disease enrichment analysis identified 791 significantly enriched disease annotations across ancestries (332 in EUR, 17 in AFR, and 442 in EAS; Fig. 3D). Diabetes mellitus was among the strongest disease-related enrichments in EUR (Enrichment score [ES] 62; GO: C1852092), while beta-cell dysfunction was prominent in EAS (ES 100; GO: C1969875), consistent with the established role of impaired pancreatic beta-cell function in T2D^36^. In AFR, granulocytic sarcoma showed the strongest disease enrichment (ES 140; GO: C0152276), followed by systolic blood pressure-related annotations (ES 73; GO: C1306620; Supplementary Tables 24). The convergence of MSigDB and Metascape enrichment on metabolic and developmental processes provides complementary evidence for the biological relevance of these pathways to CRMM, while the broader Metascape annotations further contextualise the prioritised genes across disease and functional categories.

To prioritize genes and molecular features associated with the CRMM latent factor, we integrated CRMM GWAS summary statistics with eQTL, mQTL, pQTL, and sQTL gene expression, methylation, protein abundance, and splicing summary data, respectively, using summary data using summary-data-based Mendelian randomization (SMR). Across the four molecular layers, SMR identified 47 eQTL genes, 118 mQTL genes, 19 pQTL genes and 16 sQTL genes at the predefined significance threshold (Fig. 3E and Supplementary Tables 25–28). The identified associations were further evaluated using the heterogeneity in dependent instruments (HEIDI) test to assess whether the observed associations were consistent with a shared genetic signal rather than linkage. Among the sQTL results, *ATXN2L* emerged as one of the strongest prioritized genes, with evidence of association with the CRMM latent factor (*β*_SMR_ =-0.01; *P*_SMR_= 4.05× 10^-7^; *P*_HEIDI_ = 0.3). The *ATXN2L* association was supported by four variants (rs8046271, rs153106, rs4788084, and rs12446550) and was observed across tissues relevant to CRMM, including the heart, kidney, pancreas and adrenal gland (Supplementary Fig. 6 and Supplementary Fig. 7). The direction and magnitude of the association were consistent across the relevant tissue-specific eQTL datasets (*β*SMR = −0.01; *P*_SMR_ range = 4.05× 10^-7^–1.1× 10^-5^), providing evidence for a genetically regulated expression signal underlying the association. Previous studies have linked *ATXN2L* to T2D^37^ and reported its expression in fetal kidney tissue,^38^ supporting its potential relevance to the metabolic and renal components of CRMM. *NPC1* was the next prominent eQTL-prioritized gene, with evidence of association with the CRMM latent factor (*β*_SMR_ =-0.05; *P*_SMR_= 2.8× 10^-13^; *P*_HEIDI_ = 0.2). The association was driven by variants including rs4800495, rs7239575, and rs12964689, with the strongest signal observed in tissues including the heart, artery and pancreas (*P*_SMR_ = 2.8× 10^-13^, 1.2× 10^-11^, and 1.1× 10^-09^, respectively). *NPC1* has previously been implicated in neurodegenerative disorders.^39^ The pQTL-based analysis identified 19 prioritized genes (22 gene-protein associations), several of which showed functional relevance to the nervous system. *NAGLU* was among the strongest pQTL-SMR associations (*β*_SMR_ =-0.04; *P*_SMR_= 1.6 × 10^-07^; *P*_HEIDI_ = 0.1) and showed prominent expression across cardiovascular, haematopoietic and endocrine tissues. Other prioritized protein-coding genes showed similar evidence of neurological and cardiometabolic tissue expression (n = 22 gene-protein associations; *P*_SMR_< 2.9× 10^-05^). The mQTL analysis yielded the set of prioritized molecular features, with 118 genes demonstrating association with the CRMM latent factor (*P*_SMR_< 4.8× 10^-06^). Among these, eight methylation-associated genes (*BGLAP*, *CNTNAP1*, *COMT*, *HSF2BP*, *KIAA0040*, *OR10AD1*, *RAB1A,* and *TPSG1*) could be mapped to diabetes-related phenotypes in the EWAS Atlas.^40^ Two additional prioritized genes, *MVP* and *SLC25A44*, were linked through EWAS Atlas^40^ annotations to cardiovascular phenotypes including dilated cardiomyopathy and ischaemic stroke, respectively. Together, these multi-omic analyses establish functional relevance to the latent genetic signals responsible for driving CRMM.

### Drug Repurposing Opportunities for CRMM

Existing drugs may be repurposed to treat CRMM. We used DRUGSETS,^41^ a genetically informed drug repositioning framework, to identify 10 therapeutic candidates (drugs) with potential relevance to CRMM. In AFR, manidipine, an antihypertensive calcium-channel blocker, showed significant target-gene enrichment (*P* < 3.26 × 10^-6^), with the signal primarily driven by *CACNA1C* and *CACNA1D*, which encode subunits of L-type voltage-gated calcium channels. Manidipine also showed nominal enrichment in EUR (*P* < 0.05; Fig. 3B and Supplementary Table 29), but not in EAS. As a third-generation dihydropyridine calcium-channel blocker, manidipine is used to treat hypertension. These findings support the potential relevance of calcium-channel blockade, particularly manidipine, as a potential therapeutic target in CRMM.

In EAS, nine drug-related gene sets showed significant enrichment, predominantly involving therapies used in diabetes mellitus. Exenatide, a glucagon-like peptide-1 (GLP-1) receptor agonist used to treat T2D, showed the strongest enrichment (*P* = 6.87 × 10^-13^), followed by repaglinide, an insulin secretagogue (*P* = 3.08 × 10^-12^), nateglinide (*P* = 1.73 × 10^-7^), sitagliptin (*P* = 2.56 × 10^-7^), and mitiglinide (*P* = 5.53 × 10^-7^). The enrichment of multiple insulin secretagogues, including repaglinide, nateglinide and mitiglinide, points to glucose-lowering therapies as a potential pharmacological class relevant to the genetic architecture of CRMM in EAS (Supplementary Table 29).

### Replication of CRMM Lead SNPs

To demonstrate the validity of our results, we replicated our experiment in an independent AoU cohort using multimorbid CRMM phenotypes in an attempt to isolate the same lead CRMM-associated SNPs identified in the discovery cohorts (283 in EUR, 30 in AFR, and 202 in EAS; Supplementary Note and Supplementary Table 30). After correction for multiple testing (P_BH_ < 0.05), no lead SNP was associated with the main multimorbid phenotype (all five conditions), but five SNPs in EUR (rs9272330, rs7454108, rs2866277, rs13233731, and rs440401) were associated with an alternative definition (CKD and T2D with any cardiovascular condition). These variants were located on chromosomes 6, 7, and 16, mapping to genomic regions including *HLA-DQA1*, *AUTS2*, *LOC105375508*, and *ARHGDIG* (Fig. 4 and Supplementary Table 31).

**Fig. 4.**
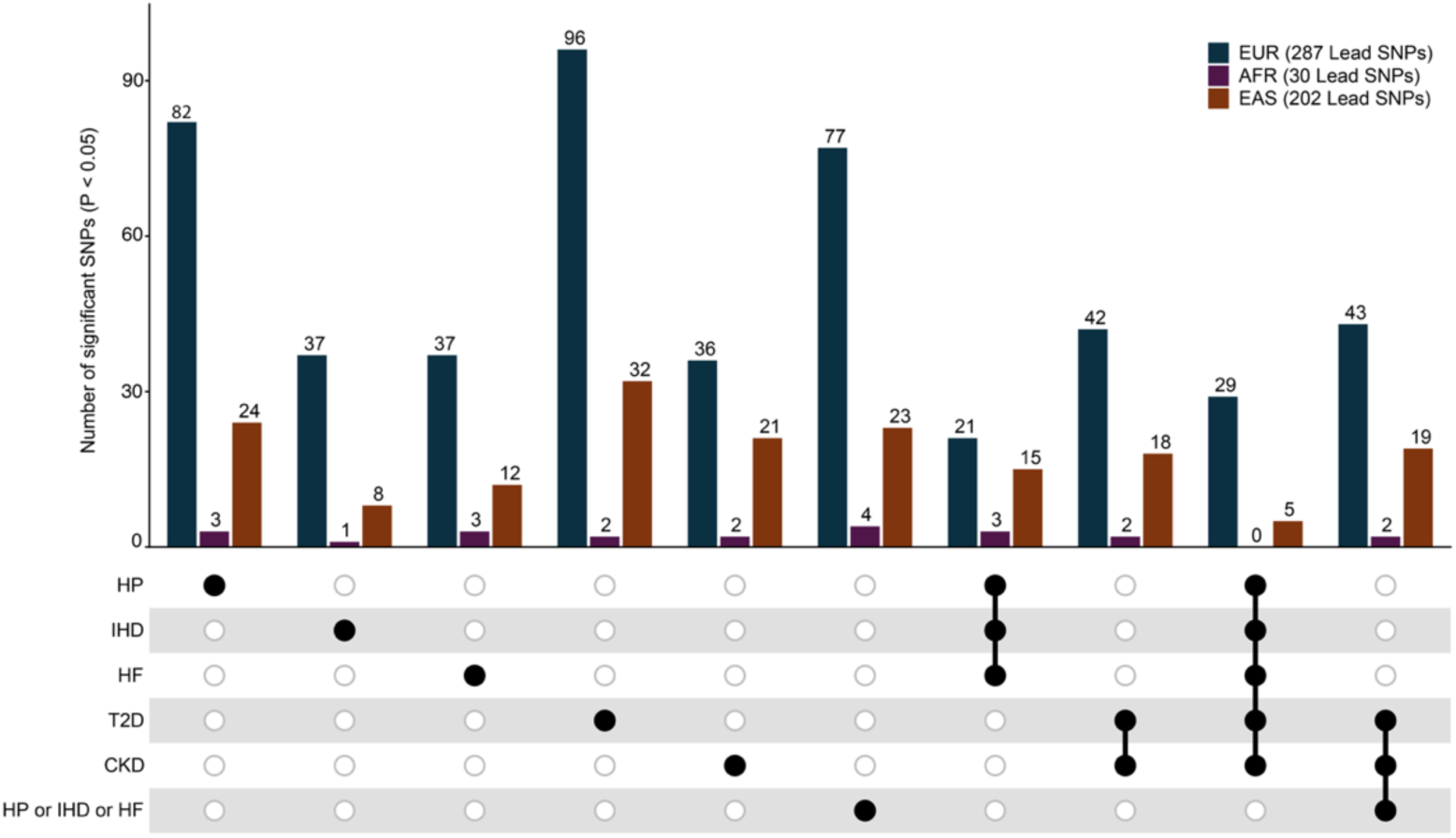
UpSet plot of significant SNPs across CRMM and its component diseases in the replication analysis. The plot shows the number and overlap of lead SNPs identified in the discovery GWAS that were significantly associated (*P* < 0.05) with CRMM and its individual components in an independent replication cohort. Bars are stratified by ancestry (EUR, AFR and EAS), with hatched segments indicating SNPs with concordant effect directions between discovery and replication analyses. The asterisk indicates that 201 EAS lead SNPs were available for CKD&T2D&HF&IHD&HP, compared with 202 for the other categories. Connected dots indicate the disease combinations represented by each intersection; combinations joined by “or” indicate the presence of at least one of the specified conditions. EUR, European; AFR, African; EAS, East Asian; CRMM, cardiometabolic and renal multimorbidity; HP, hypertension; HF, heart failure; IHD, ischaemic heart disease; T2D, type 2 diabetes; CKD, chronic kidney disease.

At a suggestive significance threshold (*P* < 0.05), 29 (10%) lead SNPs in EUR and five (3%) lead SNPs in EAS were associated with the multimorbid CRMM phenotype. When considering an alternative multimorbidity definition we identified 43 (15%), 2 (7%), and 19 (9%) significant SNPs in EUR, AFR, and EAS populations, respectively. At the level of individual CRMM components, most replicated associations in EUR were observed for T2D (n = 96; 33%), followed by HP (n = 82; 29%) and any cardiovascular condition (HF, IHD, or HP; n = 77; 27%). Among SNPs associated at *P* < 0.05, effect directions were concordant between discovery and replication for 26 of 29 and 41 of 43 SNPs in EUR for the main and alternative CRMM definitions, respectively, compared with 2 of 5 and 11 of 19 SNPs in EAS and 2 of 2 SNPs in AFR for the alternative definition (Fig. 4 and Supplementary Table 31).

### Causal Risk Factors for CRMM

We performed 2SMR using the TwoSampleMR^43^ framework to evaluate the potential causal effects of a broad range of exposures on CRMM, including 261 exposures (health conditions and clinical measurements) in EUR, 114 in AFR, and 199 in EAS populations. In the EUR ancestry, 60 exposures (23%) demonstrated significant causal associations with CRMM after multiple-testing correction (*P_BH_* < 0.05), encompassing neuropsychiatric, metabolic, endocrine, anthropometric, and organ-specific traits (Fig. 5, Supplementary Table 32, and Supplementary Fig. 8). Among the strongest risk factors were high cholesterol (odds ratio [OR] = 2.4, 95% confidence interval [CI] = 1.8–3.2), body fat (OR = 1.8, 95% CI = 1.6–2.0), and hypothyroidism (OR = 1.5, 95% CI = 1.2–1.74). In contrast, longer sleep duration (OR = 0.76, 95% CI = 0.67–0.86) and higher birth weight (OR = 0.86, 95% CI = 0.82–0.90) were associated with reduced CRMM risk. Notably, several neuropsychiatric traits, including attention-deficit hyperactivity disorder, autism spectrum disorder, dementia with Lewy bodies, and depression, exhibited positive causal effects on CRMM (OR > 1). These findings are consistent with our tissue specificity and pathway enrichment analyses, which highlighted the brain as a key implicated tissue and identified significant enrichment of gene sets related to brain development, neuronal function, and inflammatory processes (MSigDB: M4258, M11351, and M39895). In AFR populations, only two exposures (2%) were associated with CRMM, neck circumference (OR = 1.3, 95% CI = 1.2–1.4) and whole-body fat mass (OR = 0.81, 95% CI = 0.73–0.89); however, each estimate rests on a single instrument, so pleiotropy could not be assessed, and the inverse association with fat mass is inconsistent with the EUR and EAS results and should be interpreted with caution (Supplementary Table 33). In EAS populations, nine exposures (5%) were significantly associated with CRMM (Supplementary Fig. 9 and Supplementary Table 34). These associations were predominantly related to anthropometric measurements, laboratory biomarkers, autoimmune diseases, and sensory system conditions. Among the identified risk factors, body fat percentage (OR = 2.1, 95% CI = 1.8–2.3) and body mass index (OR = 1.5, 95% CI = 1.4–1.6) demonstrated the strongest positive causal effects on CRMM.

**Fig. 5.**
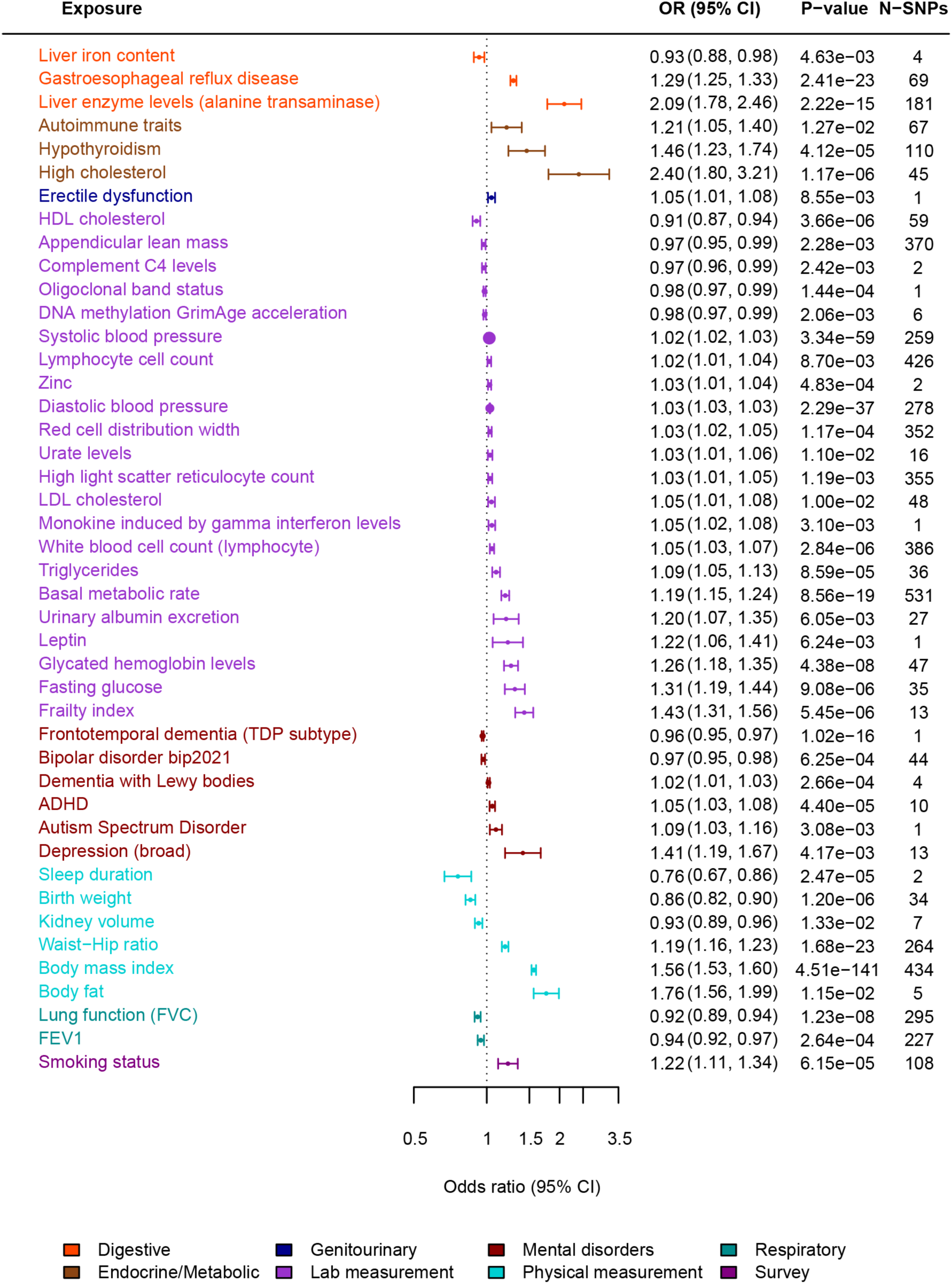
Genetically inferred causal relationships between CRMM and Clinical Phenotypes in European ancestry. Forest plot showing causal effect estimates of a range of clinical phenotypes on CRMM. Traits are grouped and colour-coded by domain, including digestive, endocrine/metabolic, genitourinary, lab measurement, mental disorders, physical measurements, respiratory, and survey. Points represent odds ratio (OR) estimates derived from the best causal method, with error bars indicating 95% confidence interval (CI). Corresponding P-values and the number of instrumental variables (N-SNPs) used for each estimate are shown. Only associations that remained significant after Benjamini-Hochberg FDR correction are presented.

### Ancestry-Specific Polygenic Risk Scores

We generated genome-wide PRS for CRMM using LDpred2-auto based on the multivariate CRMM GWAS and evaluated their predictive performance in ancestry-specific cohorts comprising 97,935 individuals of EUR, 24,966 individuals of AFR, and 4,240 individuals of EAS ancestry. Onset of CRMM was defined as the presence of all five component conditions or the co-occurrence of CKD and T2D with at least one CVD condition. Across the study populations, 719 EUR, 438 AFR, and fewer than 40 EAS participants met the criteria for CRMM, whereas 67,852 EUR, 15,061 AFR, and 3,527 EAS individuals were classified as controls, defined as being free of all five CRMM conditions (Supplementary Note and Supplementary Table 30).

The CRMM PRS was significantly associated (*P* < 0.05) with prevalent CRMM and multiple component traits across ancestries (Fig. 6). For CRMM status, the PRS explained an additional 5% of phenotypic variance in EUR populations, compared with 3% in AFR and 1% in EAS populations. Across individual CRMM component diseases, the incremental variance explained ranged from 1% to 5% in EUR, 0.2% to 3% in AFR, and 0.4% to 1% in EAS. Among the individual conditions, the CRMM PRS consistently demonstrated the strongest predictive performance for T2D across all ancestry groups. Consistent with the variance explained estimates, higher CRMM PRS was associated with increased risk of CRMM across ancestries. Each standard deviation increase in PRS corresponded to an OR of 1.84 (95% CI: 1.60–2.11) for the multimorbid phenotype in EUR populations and OR = 1.54 (95% CI: 1.28–1.85) in AFR populations; the EAS analysis could not be performed because fewer than 40 participants had all five conditions. For individual CRMM component traits, effect sizes (OR) ranged from 1.1 to 1.5 across ancestries (Supplementary Table 35). Although the estimated effect size in EAS was comparable to those observed in EUR and AFR populations, the wider confidence intervals, likely reflecting the smaller sample size and reduced number of CRMM cases, resulted in weaker statistical evidence for association.

**Fig. 6.**
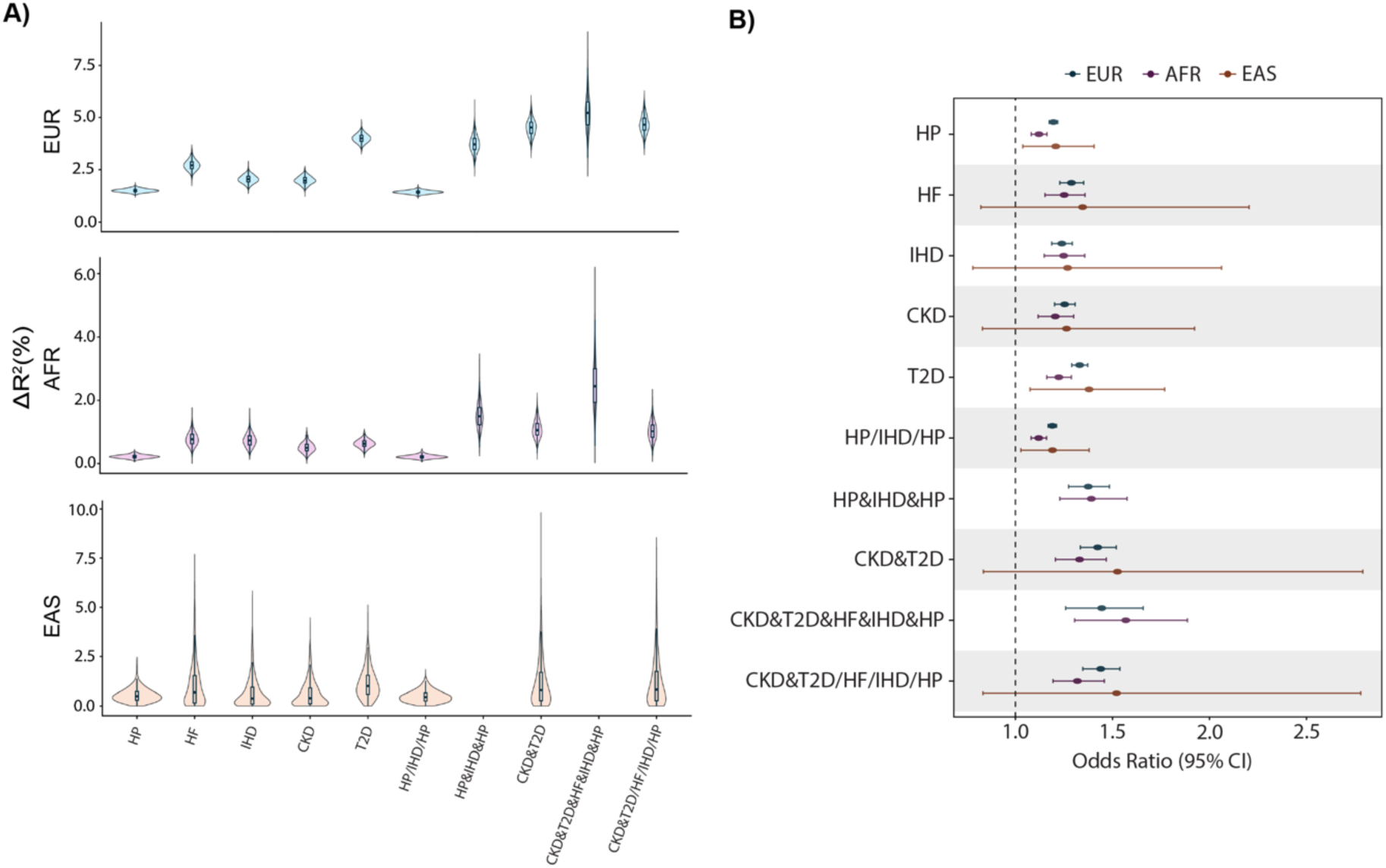
CRMM PRS performance and odds ratio estimates across European, African, and East Asian ancestries. A, Violin plots illustrating the incremental proportion of variance explained (*Δ*R^2^) by polygenic risk scores for CRMM and its component traits. B, Forest plot showing OR estimates across EUR, AFR, and EAS ancestries for CRMM and its components. Error bars represent 95% CIs and the dashed vertical line indicates an OR of 1. Disease combinations connected by “&” denote the co-occurrence of both conditions (e.g., CKD & T2D), whereas diseases separated by “/” denote the presence of any of the listed conditions (e.g., HP/IHD/HF). EUR: European; AFR: African; EAS: East Asian; HP: hypertension; HF: heart failure; IHD: ischemic heart disease; T2D: type 2 diabetes; CKD: chronic kidney disease; odds ratio (OR); confidence interval (CI).

## Discussion

To our knowledge, this is the first multi-ancestry genomic structural equation model of CRMM. Here, we show that HP, HF, IHD, CKD, and T2D share a hierarchical genetic architecture across EUR, AFR, and EAS populations, supporting CRMM as a genetically coordinated biological entity rather than a simple aggregation of independent diseases. Specifically, HP, HF, and IHD form a cardiovascular domain that, together with T2D and CKD, contributes to a higher-order CRMM factor. Modelling this shared genetic liability using Genomic SEM substantially increased discovery power, enabling the identification of novel susceptibility loci (58 in EUR, 13 in AFR, and 54 in EAS), biological pathways, therapeutic targets, and putative causal risk factors. Our findings further reveal both shared and ancestry-specific genetic determinants, highlighting the heterogeneous genetic basis of cardiometabolic multimorbidity across populations. Together, these results provide a framework for elucidating the biological mechanisms underlying multimorbidity, improving genetic risk prediction, and informing preventive and therapeutic strategies across diverse ancestral groups.

Several previous studies have leveraged Genomic SEM to investigate the shared genetic architecture underlying multimorbidity and multiple related disease conditions.^18,19,44–47^ Many of these studies have primarily adopted a common-factor or first-order latent factor model to capture shared genetic liability across disease domains, including psycho-cardiometabolic multimorbidity,^44^ psychiatric disorders,^45^ and cardiovascular-kidney-metabolic conditions.^18,19^ In contrast, other multivariate GWAS studies have implemented hierarchical latent factor structures to model both shared and domain-specific genetic components, as demonstrated in studies of frailty^46^ and metabolic syndrome.^47^ However, previous applications of these hierarchical approaches have largely focused on a single ancestry and predominantly EUR populations. Our study extends this framework by applying a hierarchical latent factor model across three ancestries (EUR, AFR, and EAS) thereby enabling the investigation of the shared genetic architecture of CRMM while accounting for ancestry-specific genetic variation.

A major finding of this study is the robust genetic correlation among CRMM components across diverse ancestries (EUR, AFR, and EAS), consistent with findings from previous single-trait genetic studies.^48^ Moreover, the underlying latent structure was further supported by factorability assessment and hierarchical clustering approaches. These results support the hypothesis that CRMM represents a biologically interconnected disease spectrum rather than a simple co-occurrence of independent conditions. By performing multivariate GWAS on the CRMM construct, we identified 148 genome-wide significant (GWS) SNPs in EUR, 16 in AFR, and 139 in EAS, even after conditional analysis, indicating both shared and ancestry-specific genetic contributions. Of our lead SNPs, 68 overlapped with loci reported by Lu, et al. ^18^ and seven of our GWS SNPs overlapped with associations reported by Wang, et al. ^19^ and one with a locus reported by Donker, et al. ^20^ which is consistent with a shared genetic architecture.

One of the most notable findings of this study was the consistent involvement of neurological biology across multiple layers of analysis. Several prioritized genes including *BDNF,*^49^ *TENM2,*^50^ and *TCF7L2,*^51^ have previously been implicated in neurological disorders, while tissue-specific analyses demonstrated enrichment in brain-related tissues and pathway analyses highlighted neuronal and neurobiological processes. These observations suggest that the biological mechanisms contributing to CRMM may extend beyond traditional cardiometabolic pathways. Increasing evidence supports extensive crosstalk between metabolic,^47^ vascular,^52^ and nervous systems through mechanisms including neuroinflammation, autonomic regulation, insulin signalling and cerebrovascular dysfunction.

Further support for this hypothesis was provided by the sQTL-SMR analyses, which prioritized *ATXN2L* as a putative functional gene underlying CRMM susceptibility. *ATXN2L* is expressed across several organs that play central roles in cardiometabolic regulation, including the heart, kidney, pancreas and adrenal gland. Previous studies have implicated this gene in obesity,^53^ type 2 diabetes,^37^ and other metabolic traits,^54^ while genes within the *ATXN* family have also been linked to RNA processing and cellular stress-response pathways that are increasingly recognized in neurodegenerative diseases^55^. Although the precise mechanism through which *ATXN2L* contributes to multimorbidity remains unclear, our findings suggest that genetically regulated splicing of this gene may represent a shared biological pathway influencing multiple CRMM components simultaneously.

The neurological enrichment observed in our genomic analyses is further supported by epidemiological evidence linking CRMM to adverse neurological outcomes. Recent cohort studies have demonstrated that increasing CRMM burden is associated with a substantially elevated risk of dementia and cognitive decline.^56^ Similarly, our Mendelian randomization analyses identified several neuropsychiatric traits as potentially causally associated with CRMM, supporting the hypothesis that shared biological mechanisms may contribute to the observed clinical overlap between these conditions. Collectively, these findings support the emerging view that cardiometabolic and neurological diseases are interconnected manifestations of overlapping biological processes rather than entirely independent pathological entities.

Notably, 24 GWS variants mapped to the *FTO* locus were shared across EAS and AFR and were also significant in EUR. However, these variants were excluded from the EUR-specific downstream analysis because they showed significant heterogeneity in EUR based on the Q_SNP_ test. Among these, rs1421085 T>C is a well-established obesity-associated variant that has been repeatedly identified in GWAS^57–59^ and supported by functional studies, including mouse models.^60,61^ Its identification within the CRMM-associated locus provides biological support for the contribution of established obesity-related genetic variation to the shared genetic architecture of CRMM.

Our genetically informed analysis identified 10 potential repurposed drugs across ancestries, including agents targeting HP (manidipine) and T2D (repaglinide, nateglinide, mitiglinide, exenatide, and sitagliptin). In the AFR population, the enrichment of manidipine,^62^ a calcium channel blocker used in HP, is consistent with established mechanisms of blood pressure regulation and underscores the role of calcium signaling pathways in CRMM risk. Notably, a previous 12-week clinical trial reported that manidipine, compared with amlodipine, not only reduced blood pressure but also improved insulin sensitivity and reduced albuminuria, suggesting potential metabolic and renal benefits beyond blood-pressure lowering^42^. In contrast, the EAS population demonstrated strong enrichment for drugs involved in insulin secretion and incretin-related pathways. Significant signals were observed for meglitinide-class agents,^63^ such as mitiglinide, repaglinide, and nateglinide, as well as GLP-1 receptor-based therapies including exenatide^64^ and the dipeptidyl peptidase 4 inhibitor sitagliptin.^65^ These findings point to a prominent role of *β*-cell function and incretin signaling in the genetic architecture of CRMM in EAS, emphasizing the importance of glucose homeostasis pathways.

Previous studies have shown that CRMM and its individual component diseases are associated with a wide range of adverse health outcomes across multiple organ systems.^66^ Building on these observations, we used 2SMR to evaluate the potential causal effects of a broad spectrum of clinical phenotypes on CRMM within each ancestry group. In EUR, we identified numerous putative causal associations spanning digestive, genitourinary, mental health, and respiratory phenotypes, in addition to several laboratory and physical measurement traits. In contrast, substantially fewer associations were observed in AFR and EAS populations, with significant findings largely restricted to phenotypes related to metabolic syndrome and sensory organ disorders. The smaller number of significant and occasionally inconsistent associations detected in AFR and EAS likely reflects reduced statistical power arising from the limited availability of genetic instruments, as many analyses were based on a single instrumental variable, precluding the application of sensitivity analyses to assess the robustness of causal estimates.^67^

Several limitations of this study should be considered. First, AMR populations were excluded because heritability estimates were unstable, likely due to limited sample size and the challenges posed by complex admixed genetic architectures. Second, age- and sex-specific genetic effects could not be evaluated due to the limited availability of stratified GWAS summary statistics across all CRMM components. Third, smoothing procedures were required to address non-positive definite covariance matrices in EAS analyses, which may have influenced the precision of SNP effect estimates. Finally, although the hierarchical latent model explained a substantial proportion of shared genetic variance, it may not fully capture the biological complexity of CRMM. Future studies incorporating additional cardiometabolic traits, longitudinal datasets and directly measured multimorbidity phenotypes will be important for refining the proposed framework.

In conclusion, our study demonstrates that CRMM is characterized by a shared but ancestrally heterogeneous genetic architecture that spans cardiovascular, renal and metabolic disease domains. By integrating multivariate genomic analyses across diverse populations (EUR, AFR, and EAS), we identify novel genetic loci, biological pathways and therapeutic opportunities that are not detectable through conventional single-disease approaches. These findings advance our understanding of the biological basis of multimorbidity and provide a foundation for future efforts aimed at improving risk prediction, prevention and treatment through genetically informed and population-aware strategies.

## Methods

### Ethics Statement

This study utilized data from FinnGen (R12), UKB, MVP, CKB, TPMI, BBJ, and AoU (All by All v1). Most analyses were conducted using publicly available summary statistics. However, the replication analyses performed using AoU required access to individual-level data through the AoU Researcher Workbench Controlled Tier. Access was obtained after completion of the required Data User Code of Conduct and Data Use Registration Agreement (DURA), in accordance with AoU data access policies. All procedures adhered fully to the data access requirements and governance policies established by each respective biobank.

### GWAS for Latent Factor Components

To investigate CRMM, we curated GWAS summary statistics for five major cardiometabolic conditions: HP, IHD, HF, CKD, and T2D (Supplementary Note). Recognising the importance of ancestral diversity, we focused on four major ancestry groups: EUR, AFR, AMR, and EAS. A detailed overview of all contributing GWAS datasets is provided in the Supplementary Tables 1 and 2 across the four ancestries. To ensure independence between discovery and replication analyses, we constructed a replication cohort from AoU data release v8, explicitly excluding individuals included in the original GWAS summary statistics. This independent dataset was subsequently used to replicate lead SNP associations and to assess PRS performance.

### Quality Control of Summary Statistics

QC procedures were conducted independently for each biobank prior to downstream analyses. All GWAS summary statistics were harmonized to the GRCh38 reference genome; among the seven biobanks, only BBJ provided GRCh37-based coordinates, which were converted to GRCh38 using the UCSC liftOver tool.^68^ We retained only autosomal variants and applied standard QC filters, including minor allele frequency (MAF) >1%, variant missingness ≤0.05, and imputation INFO score >0.9 where applicable. Following QC, approximately 8 million variants were retained for EUR ancestry, ∼14 million for AFR, ∼9 million for AMR, and ∼7 million for EAS across each of the five traits (Supplementary Table 3).

### Meta-Analysis Using METAL

Fixed effects meta analyses for the five CRMM components (HP, IHD, HF, CKD, and T2D) were performed using METAL,^69^ with analyses conducted separately for each trait and ancestry (EUR, AFR, and EAS; AMR was excluded as data was available from only one cohort). GWAS summary statistics were combined for individuals of EUR using data from FinnGen, UKBB, MVP, and AoU. For AFR, summary statistics from MVP and AoU were meta-analysed. For EAS, data from AoU, CKB, BBJ, and TPMI were integrated. Heterogeneity statistics were computed concurrently to assess cross-cohort variability, and variants showing significant heterogeneity (*P* < 5 × 10^-8^) were removed from downstream analyses (Supplementary Table 3). Bivariate LD score regression intercepts showed no evidence of sample overlap across contributing cohorts (Supplementary Table 4). Effective sample sizes (*n_eff_*) for each meta-analyzed trait were estimated following Grotzinger, et al. ^70^ using the formula:

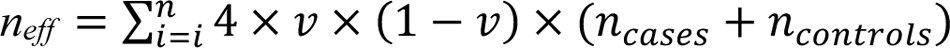

where 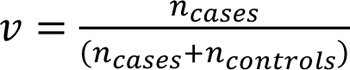 and *i* represents the number of datasets included in the meta-analysis (*i* = 4 for EUR, 2 for AFR, 4 for EAS).

### LD Score Regression

LDSC^71^ was used to generate ancestry-specific LD scores based on the AoU dataset. LD scores were constructed separately for EUR, AFR, AMR, and EAS ancestries and were restricted to autosomal SNPs with MAF >5%. In parallel, ancestry-specific reference SNP lists were created from the AoU dataset, including each variant and its population-specific MAF. Variants were retained if they had allele frequency > 1% within the corresponding ancestry or if the ancestry-specific allele count exceeded 100, following the detailed workflow described in the Supplementary Note.^27^

### Genetic Correlation and Heritability

Multivariable LD score regression was performed using the *ldsc* function implemented in the Genomic SEM R package (v0.0.5)^70^ to estimate SNP-based heritability and genetic correlations across the five CRMM components. Sample prevalence for each trait was calculated as the proportion of cases within each ancestry group (Supplementary Table 2), while population prevalence estimates were derived from previously published epidemiological studies (Supplementary Table 36). Ancestry-specific pre-computed LD scores from the AoU dataset, together with reference SNP lists excluding the Major Histocompatibility Complex (MHC) region on chromosome 6 (GRCh38: 28,510,120–33,480,577), were used for all analyses (Supplementary Note). Following QC, approximately 6 million SNPs in EUR, 8 million in AFR, 6 million in AMR, and 5 million in EAS were included in the LD score regression. Positive SNP-based heritability estimates were obtained for EUR, AFR, and EAS, allowing genetic correlations to be reliably estimated for these ancestries. However, the AMR dataset produced a negative heritability estimate, rendering genetic correlation estimation infeasible (Supplementary Table 5). Consequently, AMR was excluded from subsequent multivariable LDSC analyses.

### Factorability of Genetic Correlation

Factorability was assessed using three complementary approaches: (1) the KMO MSA, (2) Bartlett’s test of sphericity, and (3) parallel analysis. The KMO MSA^28^ is based on the squared elements of the anti-image genetic correlation matrix relative to the squared elements of the original genetic correlation matrix, yielding MSA values for each component and overall. Values above 0.5 indicate data adequacy for factor analysis. In contrast, Bartlett’s test^29^ evaluates whether the genetic correlation matrix is an identity matrix; a significant result (*P*-value <0.05) suggests the matrix is suitable for factor analysis. Parallel analysis^30^ was used to determine the number of factors to retain for EFA by comparing eigenvalues from the observed genetic correlation matrix against those from 1,000 randomly generated correlation matrices of the same size, using principal axis factoring. The resulting ancestry-specific genetic correlation matrices were then used to perform the KMO MSA, Bartlett’s test of sphericity, and parallel analysis, implemented using the *psych* R package (v2.5.3)^72^. For the EAS ancestry, the estimated genetic correlation matrix was not positive definite, as indicated by the presence of negative eigenvalues. Because both the KMO MSA and Bartlett’s test require a positive definite correlation matrix, these assessments could not be conducted for EAS. However, parallel analysis, which relies only on the eigenvalue decomposition of the observed matrix and does not require matrix inversion, was still performed (Supplementary Fig. 2).

### Confirmatory Factor Analysis

CFA was performed independently for each ancestry using the Genomic SEM R package (v0.0.5).^70^ The results from the factorability assessment and EFA were used to construct the multimorbidity factor (Supplementary Note and Supplementary Table 6–8). Traits with EFA factor loadings ≥ 0.50 were retained for inclusion in the CFA multimorbidity factor model. All CFA models were specified using unit variance identification (i.e., fixing the variance of the latent factor to one) to facilitate interpretation of model fit. Factor loadings were estimated using the diagonally weighted least squares method (DWLS). Model fit indices were obtained following the recommendations of Grotzinger, et al. ^70^, including the *χ*^2^-statistic, CFI, AIC, and SRMR. While the *χ*^2^-statistic was used to compare nested models via difference testing, AIC, CFI, and SRMR were used to assess overall model fit across competing models. Consistent with prior recommendations, models with CFI ≥0.95 were considered to demonstrate good fit, and those with SRMR ≤0.10 were considered to demonstrate acceptable fit.^70^

### Genomic Structural Equation Modelling

Multivariate GWAS was performed using the Genomic SEM R package (v0.0.5),^70^ leveraging the hierarchical factor structure derived from the CFA (Supplementary Table 9). A latent CVD factor was first constructed from HP, HF, and IHD. This CVD factor was subsequently combined with CKD and T2D to define a higher-order CRMM factor. SNP associations were estimated by regressing the CRMM latent factor on each SNP while specifying the factor model using unit loading identification, in which one indicator loading per factor was fixed to 1 for identification: HF (for CVD) and T2D (for CRMM) in EUR analyses, and HP (for CVD) and CVD (for CRMM) in both AFR and EAS analyses. SNP effect estimation was carried out using DWLS. Additional QC filters were applied, excluding SNPs with MAF < 0.01 and retaining only variants present across all ancestry-specific input GWAS. Reference SNP lists were derived from the AoU version 8 dataset (Supplementary Note). After QC, 8,509,763 SNPs in EUR, 14,602,233 SNPs in AFR, and 6,997,513 SNPs in EAS were retained for multivariate GWAS. Variants that generated warnings from *lavaan*^73^ during model fitting were excluded from analysis. To address non-positive definite covariance matrices in the EAS dataset, we additionally removed SNPs that required extreme smoothing, defined as a change in the z-statistic greater than 1.96 when comparing pre and post-smoothing values. This resulted in the removal of 227 SNPs in EUR, 7 SNPs in AFR, and 322 SNPs in EAS.

Heterogeneity statistics (Q_SNP_)^70^ were computed for individual SNPs to test deviations from the null hypothesis that each SNP influences the phenotype solely through a specified latent factor. Q_SNP_ was calculated for both the first-order CVD factor and the second-order CRMM factor. We evaluated model fit using chi-square difference tests comparing the common pathway and independent pathway models (Supplementary Fig. 10). SNPs showing evidence of heterogeneity were defined as those with Q_SNP_ *P* < 1.25 × 10^-8^, consistent with the GWS threshold (*P* < 5 × 10^-8^⁄4; four ancestry groups). For SNPs identified as heterogeneous by Q_SNP_, we additionally identified variants in LD using the AoU (v8) reference panel in PLINK v1.9^74^ (500 kb window, 50 SNPs, *r*^2^ ≥ 0.6). Both significant Q_SNPs_ and their LD proxies were excluded from all downstream analyses. For the CRMM GWAS, Q_SNP_-significant SNPs (and their LD proxies) were removed if they were associated with either the CRMM or CVD factors.

Among GWS variants, LD-based clumping was performed in PLINK^74^ to identify independent and lead SNPs. Independent significant SNPs were first obtained by clumping all GWS variants using an LD threshold of *r*^2^ < 0.6 within a 250kb window, consistent with the locus definition implemented in FUMA.^80^ Lead SNPs were then identified by applying a stricter second clumping step to the independent SNP set, using *r*^2^ < 0.1 within the same 250kb region. These lead SNPs were subsequently analyzed using GCTA COJO for conditional and joint association testing.^75^ A stepwise selection procedure with default parameters and a significance threshold of *P* < 1.25 × 10^-8^ was used to identify variants that remained conditionally and jointly associated after accounting for ancestry specific LD patterns. CRMM associated SNPs were classified as non-novel if they were previously reported in the NHGRI EBI GWAS Catalog v1.0.^76^ To further assess the potential regulatory and functional effects of the identified SNPs, variants were annotated using Combined Annotation Dependent Depletion (CADD)^77^ and RegulomeDB scores.^78^

### Cross-Ancestry Heterogeneity and Genetic Correlation

Cross-ancestry heterogeneity in SNP effect estimates was assessed using Cochran’s Q test, based on ancestry-specific effect estimates and standard errors for EUR, AFR, and EAS. Overall heterogeneity across the three ancestries and pairwise heterogeneity between each ancestry pair were evaluated. *P*-values were adjusted for multiple testing using the Benjamini-Hochberg procedure, with adjusted (*P* < 0.05) considered statistically significant.

Cross-ancestry genetic correlations for CRMM GWAS conducted in EUR, AFR, and EAS ancestry groups were estimated using Popcorn software v1.1.^31^ For each ancestry group, cross-population LD scores were generated using the PLINK binary genotype files from 10,000 randomly selected individuals per ancestry from the AoU dataset (v8). During LD score construction, the extended MHC region on chromosome 6 (28–34 Mb) was excluded due to its complex LD structure. Only SNPs with MAF >0.01 in the respective CRMM GWAS summary statistics were retained for analysis. These filtered summary statistics, together with the computed cross-population LD scores, were used as input to Popcorn^31^ to estimate cross-ancestry genetic correlation and genetic impact correlations using a maximum-likelihood estimation (MLE) framework.

### Gene Mapping and Pathway Analysis

SNPs were mapped to their nearest corresponding genes using the Ensembl Variant Effect Predictor (VEP) v115 (Supplementary Note).^79^ MAGMA v1.10^32^ was used to perform gene-based, gene-set, and gene-property analyses. For the gene-based analysis, SNPs were assigned to protein coding genes and aggregated using MAGMA’s SNP-wise mean model, accounting for LD using ancestry-specific PLINK reference files derived from the AoU v8 dataset. This mapping process covered 20,137 protein coding genes with Entrez Gene IDs (GRCh38).^32^ Statistical significance was defined as *P*< 2.48× 10^-6^ (Bonferroni-corrected threshold). Significant genes identified in each ancestry group were used for downstream pathway enrichment analysis using Metascape v3.5.20260201,^35^ to identify biological pathways and disease pathways enriched among CRMM associated genes. Enriched pathways were identified based on an FDR-corrected significance threshold of P_FDR_ < 0.05. Gene set analysis in MAGMA was performed to identify gene sets enriched for CRMM associated genes via competitive testing. Gene sets were obtained from the MSigDB v2025.1,^34^ following the FUMA workflow,^80^ including 7,561 curated gene sets and 16,228 ontology gene sets. Significance of competitive gene testing was determined using Bonferroni-corrected thresholds. Gene property analysis in MAGMA was conducted to test whether tissue specific gene expression profiles were associated with CRMM gene level association statistics. Analyses were performed using 54 detailed tissues and 30 general tissue groups from GTEx v10^33^ across the three ancestry groups. Significance of tissue associations was determined using Bonferroni-corrected thresholds. A detailed description of GTEx v10^33^ RNA-seq preprocessing is provided in the Supplementary Note.

Finally, gene prioritization for EUR was performed using the SMR Portal,^81^ identifying genes showing evidence of pleiotropic association between the CRMM factor and molecular traits including expression quantitative trait loci (eQTLs), splicing QTLs (sQTLs), methylation QTLs (mQTLs), and protein QTLs (pQTLs). The SMR framework uses genetic variants as instrumental variables to test whether a molecular trait (gene expression, splicing, methylation, or protein levels) and the outcome (here, the CRMM latent factor) share a genetic signal, which is consistent with either a causal or a pleiotropic relationship. Statistical significance was determined using Bonferroni correction for multiple testing, with corrected *P* values below the corresponding significance threshold considered significant, followed by the HEIDI test to reject associations driven by linkage disequilibrium. This approach enabled systematic prioritization of genes likely mediating CRMM risk through molecular regulatory mechanisms (Supplementary Note).

### Drug-Gene Set Analysis

Drug-gene set analysis was performed using the framework implemented in DRUGSETS^41^ to identify potential therapeutics interacting with CRMM-associated genes. Drug-gene interaction data were obtained from the Drug-Gene Interaction Database (DGIdb; accessed 16 February 2026)^82^ and the Drug Repurposing Hub (accessed 19 August 2026)^83^. For each database, drug-gene pairs were filtered to retain only drugs with at least two annotated genes, and gene entries were matched to gens associated with CRMM GWAS. This resulted in 2,816 drug-gene pairs from DGIdb and 8,445 drug-gene pairs from the Drug Repurposing Hub. Competitive gene-set analysis was conducted using MAGMA v1.10^32^ to test whether genes within each drug-defined set showed stronger associations with CRMM than genes outside the set, using gene-level association statistics derived from the CRMM GWAS. Drug-gene sets were considered statistically significant after applying Bonferroni correction to the number of tested drug sets (n= 11261; *P* < 4.4 × 10^-6^; Bonferroni-corrected threshold).

### Lead CRMM SNP Replication Analysis

To assess whether SNPs identified from the multivariate GWAS were enriched among individuals with multimorbidity, we performed replication analyses using individual-level data from the AoU v8 dataset. The available sample comprised 220,371 EUR, 72,899 AFR, and 9,584 EAS ancestry participants. To generate independent replication cohorts, individuals overlapping with the discovery cohort (AoU v7) were excluded, resulting in replication samples of 97,935 EUR, 24,966 AFR, and 4,240 EAS participants (Supplementary Note; Supplementary Table 30). Cases were defined based on the five CRMM component conditions (HP, IHD, HF, T2D, and CKD). Different case groups were constructed to examine the representation of the identified CRMM SNPs across disease profiles; specifically, all five CRMM component conditions were included as individual case groups, along with several multimorbid cohorts defined as individuals with all three cardiovascular conditions (HP, IHD, HF), those with at least one cardiovascular condition, those experiencing all five CRMM conditions, and those with T2D and CKD together with at least one cardiovascular condition. Controls were defined as participants without any of the five CRMM conditions. Association testing was performed using Firth-penalized logistic regression implemented in the *logistf* function of the *logistf* R package (v1.26.1).^84^ Lead CRMM SNPs (283 EUR, 30 AFR, and 202 EAS variants) were tested under an additive genetic model, adjusting for age, sex, and the first 10 principal components (PCs). SNPs were considered nominally significant at *P <* 0.05, while strict statistical significance was determined using FDR correction (*P_BH_* < 0.05).

### Two-Sample Mendelian Randomization

To investigate the potential causal effects of a broad range of clinical phenotypes on CRMM, we conducted 2SMR, treating CRMM as the outcome. Q_SNP_-significant variants and their LD-linked proxies were excluded from the CRMM GWAS summary statistics. Exposure GWAS datasets were sourced from the IEU OpenGWAS Project (accessed 22 December 2025).^86^ For the EUR analysis, 30,674 traits were initially available; studies derived from biobanks contributing to the construction of the multivariate CRMM factor (AoU, UKB, FinnGen, and MVP) were excluded to avoid sample overlap. Equivalent ancestry-specific filtering procedures were applied in AFR and EAS analyses, whereby exposure GWAS derived from cohorts contributing to the corresponding ancestry-specific CRMM GWAS were removed. Additional exclusions were applied to remove GWAS based on gut microbiome traits, COVID-19 outcomes, proteomics, metabolomics, and eQTL data. For traits with multiple GWAS entries, we retained the version with the largest number of valid instruments, and when tied, the GWAS with the largest sample size. Traits closely overlapping with the CRMM component outcomes (CKD, CVD, and T2D) were also removed. After filtering, 261 exposure traits were retained for EUR, 114 for AFR, and 199 for EAS. All 2SMR analyses were performed using the *TwoSampleMR* R package (v0.6.21).^86^ Instrumental variables (IVs) for each exposure GWAS were selected using the default clumping parameters implemented in *TwoSampleMR*^86^ (GWS threshold *P*<5 × 10^-8^, LD r^2^ = 0.001, clumping distance = 10 Mb), with clumping performed using ancestry-matched 1000 Genomes Project phase 3 (EUR, AFR, and EAS) reference panels. The selected IVs were then harmonized with the CRMM outcome GWAS to ensure consistent alignment of effect alleles between exposure and outcome datasets. We further excluded palindromic variants with intermediate allele frequencies, as well as instruments absent from the outcome GWAS.

Five MR methods were applied to each exposure-outcome pair, depending on the number of available IVs. When only a single IV was available, causal effects were estimated using the Wald ratio.^87,88^ For analyses with two or more IVs, the IVW method^89^ was used as the primary estimator. When more than three IVs were available, we additionally applied the WM and MR-Egger regression methods. When more than three IVs were available, MR-PRESSO was also performed, and its outlier-corrected estimate was used only when the global test was significant and enough IVs remained after outlier removal. Heterogeneity in IVW estimates was assessed using Cochran’s Q-test, horizontal pleiotropy was evaluated using the MR-Egger intercept^90^ and the MR-PRESSO global test.^91^ The most appropriate causal estimate for each exposure-outcome pair was selected following the decision framework proposed by Kim, et al. ^92^ and Park, et al. ^47^ (Supplementary Fig. 11). Causal associations were considered statistically significant at *P_BH_* < 0.05.

### Ancestry Specific Polygenic risk scores

PRS were constructed using the LDpred2-auto framework implemented in the bigsnpr R package.^85^ PRS were evaluated in an independent cohort from AoU (v8), after removing all individuals who participated in the discovery GWAS to avoid sample overlap (Supplementary Note). All analyses, including LD matrix construction and PRS estimation, were performed in an ancestry-specific manner. Ancestry-matched LD reference panels were used for LD score construction. For PRS derivation, we used ancestry-specific CRMM GWAS summary statistics after exclusion of variants flagged by Q_SNP_ filtering. The SNP set used for PRS construction consisted of HapMap3 variants lifted over to GRCh38.

To assess PRS predictive performance, we implemented a bootstrap resampling framework with 1,000 replicates. In each replicate, controls were randomly sampled at a 1:1 ratio to match the number of cases. Logistic regression models were fitted for the composite CRMM outcome (defined as individuals with all five conditions or those with diabetic kidney disease and at least one cardiovascular outcome) as well as for each CRMM component. Models were fitted only when the total sample size for the outcome was at least 40 individuals. In all models, PRS was included as the primary predictor, with age, sex, and the first 10 genetic PCs included as covariates. Model performance was evaluated using incremental Nagelkerke’s R^2^, calculated as the difference between the full model (including PRS and covariates) and a null model containing covariates only. Final performance estimates were obtained by calculating the mean R^2^ values across all bootstrap replicates.

## Code Availability

Code used for the primary analyses is available on GitHub.

## Data Availability

Multivariate GWAS summary statistics are available through the <u>GWAS Catalog</u> under accession ID XXXX. Ancestry-specific linkage disequilibrium (LD) scores and SNP lists derived from the All of Us v8 dataset are publicly available on <u>GitHub</u>. Additional GWAS summary statistics and replication datasets were obtained from publicly accessible resources, including <u>FinnGen</u>, <u>UK Biobank</u>, and <u>the Million Veteran Program</u>, <u>China Kadoorie Biobank</u> <u>PheWeb</u>, <u>BioBank Japan PheWeb</u>, <u>Taiwan Precision Medicine Initiative PheWeb</u>,, the <u>All</u> <u>by All All of Us platform</u>, and <u>the GWAS Catalog</u>. Functional annotation and downstream analyses utilized publicly available resources, including <u>MAGMA</u> auxiliary files, GTEx v10 expression reference data, <u>MSigDB gene set</u> collections, <u>OpenGWAS</u>, <u>GENCODE</u> gene annotations, <u>DGIdb</u>, and <u>the Drug Repurposing Hub</u>. Access to All of Us Controlled Tier data was obtained through the All of Us Researcher Workbench in accordance with program policies.

## Acknowledgments

This research used data from multiple large-scale biobank initiatives, and we gratefully acknowledge the participants and investigators whose contributions made this work possible. The All of Us Research Program is supported by the National Institutes of Health, Office of the Director, through multiple funding mechanisms, including Regional Medical Centers (1 OT2 OD026549; 1 OT2 OD026554; 1 OT2 OD026557; 1 OT2 OD026556; 1 OT2 OD026550; 1 OT2 OD026552; 1 OT2 OD026553; 1 OT2 OD026548; 1 OT2 OD026551; 1 OT2 OD026555), IAA #: AOD 16037, Federally Qualified Health Centers (HHSN 263201600085U), the Data and Research Center (5 U2C OD023196), the Biobank (1 U24 OD023121), the Participant Center (U24 OD023176), the Participant Technology Systems Center (1 U24 OD023163), Communications and Engagement (3 OT2 OD023205; 3 OT2 OD023206), and Community Partners (1 OT2 OD025277; 3 OT2 OD025315; 1 OT2 OD025337; 1 OT2 OD025276). The program would not be possible without the partnership and generosity of its participants. We also acknowledge the participants and investigators of the FinnGen study for their valuable contributions. We gratefully acknowledge the UK Biobank and thank its participants and research staff for their valuable contributions to this research. We are grateful to the BioBank Japan Project and its participants, as well as to all participants and investigators from the Taiwan Precision Medicine Initiative. We further acknowledge the Million Veteran Program, including former staff members and volunteers, and especially the participants who served in the military and generously contributed to this research. Finally, we thank the developers of Genomic SEM for providing essential tools that supported this analysis.

## Funding

This work was supported by the National Health and Medical Research Council (NHMRC) Ideas Grant 2025 (ID: 2038558).

## Author Contributions

SHN conceived and led the study. ADB designed and performed the analyses. VA generated SHN conceived, supervised, and led the study. ADB designed and performed the analyses and generated all figures. VA compiled, organized, and finalized the figure panels for presentation. RAL co-supervised the project and, together with VA, NKT, RM, AJ, LY, and SS, critically reviewed and refined the manuscript. SS curated the cardiovascular disease phenotype list. All authors read and approved the final manuscript.

## Competing Interests

All authors declare no competing interests.

## Supplementary Note

### Multimorbidity

Multimorbidity (MUL) is defined as the co-occurrence of two or more diseases in a single individual^93^. In this study, we focus on the multimorbidity of type 2 diabetes (T2D), cardiovascular disease (CVD), and chronic kidney disease (CKD), a combination often referred to as cardio-renal-metabolic (CRM) or cardio-kidney-metabolic (CKM) multimorbidity. As highlighted by Marassi and Fadini ^1^ these conditions represent major global health challenges of the twenty-first century. CKD has a prevalence of approximately 40% among individuals with T2D and 50% among those with heart failure (HF).^94^ Additionally, the coexistence of CVD and CKD in individuals with T2D increases disease progression, leading to increased morbidity and mortality.^94^ Furthermore, individuals with T2D have a twofold higher risk of coronary heart disease, stroke, and vascular-related mortality. Diabetes is also the leading cause of both CKD and end-stage kidney disease (ESKD).^95^

Given the complex bidirectional and compounding relationships among these conditions, where one disease can act as a risk factor for, or accelerate the progression of another, it is essential to investigate their underlying genetic etiology in conjunction with clinical and environmental factors. In this study, we define multimorbidity based on T2D, CKD, and CVD. While T2D and CKD are clinically well-defined phenotypes that can be reliably identified using ICD codes or phecodes (e.g., EM 202.2 for T2D and GU 582.2 for CKD), CVD represents a broader spectrum of conditions and is more challenging to operationalize as a single trait.

To address this, we began with the All of Us (AoU) GWAS summary statistics and considered the major CVD-related phecodes available in the AoU all-by-all results (V1). From this set, we retained traits that met the following criteria: (i) non-negative SNP-based heritability estimates, (ii) heritability standard errors less than half of the estimate, and (iii) traits present in at least two of the ancestry groups. This filtering resulted in the following traits: atherosclerosis (ASCVD; CV 436), heart failure (HF; CV 424), hypertension (HP; CV 401), ischemic heart disease (IHD; CV 404), T2D (EM 202.2), and CKD (GU 582.2) (Supplementary Table 37). Based on clinical considerations and disease categorization, ASCVD was grouped under the broader IHD, and only IHD was carried forward for analysis. Accordingly, the CVD latent construct was defined using IHD, HF, and HP. Furthermore, CVD, along with T2D and CKD, was used to construct CRM multimorbidity.

### Genetic diversity in the AoU (v8)

In AoU, we excluded 30,584 related individuals and 987 individuals flagged by the AoU program from the initial 414,830 individuals with WGS short-read sequencing data (v8). We further restricted the dataset to individuals whose sex at birth was either male or female and whose age was under 100 years. After these filters, 379,257 participants remained (Supplementary Table 38). Ancestry groups were predicted from the genetic data by the AoU program itself, based on internal analyses (Supplementary Fig. 12).^96^

### LD Score Construction Using LDSC

LD scores^71^ are required for each SNP within each ancestry group to construct genetic correlations and to calculate SNP-based heritability. To the best of our knowledge, the developers of the LD Score Regression (LDSC) package^71^ have only provided ancestry-specific LD scores for European and East Asian populations. Moreover, these resources are limited to approximately one million HapMap SNPs and are based on the GRCh37 (hg19) reference genome. In contrast, the GWAS summary statistics we obtained are aligned to the GRCh38 (hg38) reference. These constraints, which also apply to LD scores used in Genomic SEM,^70^ restrict the feasibility of cross-ancestry analyses.

Given the diverse genetic ancestry represented in the AoU dataset, with a total sample size of approximately 500,000 individuals, we performed custom LD score regression within AoU, which is based on the GRCh38 (hg38) reference genome. Analyses were conducted across all ancestry groups and restricted to autosomal SNPs. Prior to running LDSC, we generated ancestry-specific PLINK^74^ files that included centimorgan (cM) values as the genetic distance metric, derived from the hg38-based genetic map available from the Broad Institute repository.^97^ With the ancestry-specific PLINK files containing cM values prepared, the next step was to perform LD score regression using the LDSC package,^71^ which generated ancestry-specific LD scores for EUR, AFR and EAS ancestries (detailed workflow in Supplementary Fig. 13).

### Exploratory factor analysis

EFA was conducted to investigate the latent factor structure underlying the ancestry-specific genetic correlation matrices. EFA was implemented using the *fa* function in the *psych* R package (v2.5.3),^72^ and models specifying one to three factors were evaluated for each ancestry. For the EUR and AFR ancestries, factor extraction was carried out using maximum likelihood estimation, whereas for the EAS ancestry (where the genetic correlation matrix was not positive definite) minimum residual extraction was applied. To account for potential correlations among latent factors, *promax* rotation was used across all three ancestries. Components with absolute factor loadings ≥0.40 were retained, and communality estimates were computed for each trait. Additionally, to assess whether traits clustered in a manner consistent with the identified factor structure, hierarchical cluster analysis of the genetic correlation matrices was performed using the *hclust* function from the *stats* R package, applying Ward’s agglomerative clustering algorithm.^98^

### Reference SNP List for LD Score Regression

A reference SNP list was generated for use in LD score regression analyses, including variant-level information on MAF. The reference file was constructed using the AoU v8 dataset following ancestry-specific quality control procedures. Variants were retained if they met the criteria of MAF > 1% or minor allele count (MAC) > 100 within each ancestry group. Supplementary Table 38 summarizes the number of SNPs retained for each ancestry after quality control filtering, representing the set of common variants used in downstream analyses.

### Functional Annotation Using Variant Effect Predictor

Variants were functionally annotated using the Ensembl Variant Effect Predictor (VEP; v115).^79^ Annotation was performed against the GRCh38 human reference genome using the primary assembly FASTA file obtained from GENCODE.^99^ Analyses were conducted in offline mode with a locally installed Ensembl cache. Canonical transcripts were prioritized, and a single representative consequence per allele was retained. Comprehensive functional annotations were obtained using the *--everything* flag. To improve computational efficiency, variant annotation was parallelized across eight CPU threads.

### Pre-Processing of Gene Expression Data

RNA-seq data for gene property analyses were obtained from GTEx v10.^33^ Gene expression levels were quantified as transcripts per million (TPM), with Ensembl gene IDs used as primary identifiers. In total, 19,616 samples were available across tissues. From 59,033 annotated genes, filtering was applied such that genes were retained if the average expression (TPM) exceeded 1 in at least one of the 54 tissues. Expression values were winsorized at an upper threshold of 50 (i.e., TPM > 50 was set to 50) to limit the influence of extreme values. Subsequently, gene expression measures were log-transformed using log_2_(TPM+1), and the average expression per tissue (across either 54 detailed or 30 general tissue categories) was computed.

The resulting 34,447 genes were subsequently mapped to the NCBI protein-coding gene list,^32^ corresponding to the gene set used for annotation in MAGMA.^32^ Gene identifiers were then converted from Ensembl IDs to Entrez gene IDs using the org.Hs.eg.db annotation R package.^100^ In cases where a single Ensembl gene ID mapped to multiple Entrez gene IDs, a single representative Entrez ID was retained by averaging expression values across tissues for duplicated entries. These expression values were then used as covariates, conditioning on the mean expression across all tissues. After harmonization with ancestry-specific gene-level correlation matrices, 17,278 genes in the EUR, 17,333 genes in the AFR, and 17,166 genes in the EAS datasets were retained for downstream analyses.

### Replication Cohort Description

The replication cohort was constructed using data from the AoU (Supplementary Table 30). To ensure independence from the discovery GWAS, participants included in the v7 dataset were excluded from the v8 release, and only individuals not previously included in the GWAS analysis were considered. From this subset, we retained participants with available short-read whole-genome sequencing data and applied standard quality control procedures, excluding samples flagged for QC issues as well as related individuals. Among the remaining participants, five CRMM phenotypes HP, IHD, HF, T2D, and CKD, were extracted. Given the longitudinal nature of AoU electronic health record data, individuals were classified as cases for each condition based on the earliest recorded diagnosis date. Multimorbidity cohorts were subsequently defined to capture disease clustering, including individuals with all three cardiovascular conditions (HP, IHD, and HF), those with both T2D and CKD, those with all five conditions (HP, IHD, HF, T2D, and CKD), and those with at least one cardiovascular condition (HP, IHD, or HF) in combination with both T2D and CKD.

The cohort had a mean age of 55.8 years, with a predominance of women (60.8%). Individuals of EUR descent made up the majority (60.8%) of the cohort, followed by AFR ancestry. EAS participants represented the smallest proportion (n = 4,240; 2.6%). HP was the most common single phenotype across the overall cohort. When examining combinations of diseases, the largest proportion of participants fell within the CVD group, defined by the presence of at least one CVD outcome, accounting for 45,824 individuals (29.1%). In contrast, CRMM, defined as the coexistence of all relevant conditions, was relatively rare at 1,506 individuals (1.3%) of the overall cohort. Subgroup analyses highlighted important ancestry-specific differences. The highest proportion of CRMM was observed among individuals of AFR (2.8%), relative to their sample size. Conversely, CRMM cases were minimal among EAS participants (<40 individuals). A similar pattern was noted for more severe CVD presentations (i.e., individuals with all three major CVD outcomes such as HF, IHD, and HP), where fewer than 40 cases were observed in the EAS group. In comparison, these outcomes were present in 3.1% of EUR individuals and 5.6% of AFR individuals. Across all ancestry groups, hypertension remained the most prevalent condition. These findings are detailed in Supplementary Table 30.

For the replication analysis, lead SNPs identified in the discovery GWAS were selected for each ancestry, comprising 287 SNPs in EUR, 30 in AFR and 202 in EAS. These variants were extracted from the corresponding genotype datasets using PLINK for subsequent analyses. SNPs with only one case observation were excluded, resulting in 283, 30 and 202 SNPs retained for EUR, AFR and EAS, respectively. Specifically, four SNPs were excluded from the EUR analysis (chr10:58244248:G:T, chr2:35112829:A:G, chr1:156579091:G:C and chr1:144657032:T:C), and one SNP was excluded from the EAS analysis of the multimorbidity phenotype CKD&T2D&HF&IHD&HP (chr12:108236003:G:A).

### Genomic Structural Equation Modelling of CVD Latent Factor

In addition to constructing the CRMM latent factor, we derived a CVD latent factor using HP, IHD, and HF. We subsequently performed multivariate GWAS to identify SNPs associated with the CVD latent factor across EUR, AFR, and EAS ancestries (Methods; Supplementary Fig. 10). After quality control, 8,508,846 SNPs were retained in EUR, 14,602,224 in AFR, and 6,997,191 in EAS. Following GWAS, we calculated the heterogeneity statistic (Q_SNP_) for the CVD factor using independent and common pathway models. Removal of Q_SNP_ outliers and their associated LD proxies resulted in 8,457,274 SNPs in EUR, 14,600,283 in AFR, and 6,991,910 in EAS. The effective sample sizes for the CVD GWAS were 329,274 in EUR, 63,488 in AFR, and 81,131 in EAS (Supplementary Table 10).

We identified 12,529 genome-wide significant (GWS; *P*<1.25 × 10^-8^) SNPs in EUR, 278 in AFR, and 390 in EAS associated with the CVD latent factor (Supplementary Fig. 14 and Supplementary Fig. 15). Among these, 337 (EUR), 30 (AFR), and 48 (EAS) were independent lead SNPs, of which 227, 18, and 29, respectively, were retained following COJO analysis. Notably, 101 SNPs in EUR, 13 in AFR, and 19 in EAS were classified as novel based on comparison with the GWAS Catalog, even after conditional and joint analysis (Supplementary Tables 39-41). COJO-prioritized variants mapped to 116 genes in EUR, 13 in AFR, and 20 in EAS. Cross-ancestry comparison revealed that *CACNB2* was shared between EUR and AFR, while *WNT2B*, *MECOM*, *PLEKHG1*, *LSP1*, and *RGL3* were common to EUR and EAS. Two genes, *CASZ1* and *CACNA1D*, were consistently implicated across all three ancestries.

### Parameters used for SMR analysis

Summary data-based Mendelian randomization (SMR) analysis was performed to prioritize genes associated with the CRMM factor by integrating molecular quantitative trait loci (QTL) datasets, including expression QTLs (eQTLs), splicing QTLs (sQTLs), methylation QTLs (mQTLs), and protein QTLs (pQTLs) through the SMR portal (accessed 25 November 2025). To improve biological relevance, tissue-specific analyses were restricted to tissues implicated in the CRMM spectrum rather than surveying all available tissues. Specifically, eQTL analyses were conducted in adrenal gland, blood vessel, heart, kidney, and pancreas tissues. sQTL analyses were performed in adrenal gland, heart, kidney, and pancreas tissues, whereas pQTL and mQTL analyses were restricted to blood-derived datasets. To further characterize prioritized methylation signals, significant CpG sites were mapped to previously reported disease and trait associations using the EWAS Atlas database (accessed 7 September 2026).^40^

### Identification of potential CRMM drug

The potential relevance of these findings to CRMM is further informed by the established clinical profiles of the enriched drugs. Exenatide, a glucagon-like peptide-1 (GLP-1) receptor agonist, was evaluated in the EXSCEL cardiovascular outcomes trial involving 14,752 participants with T2D.^101^ Although exenatide was non-inferior to placebo with respect to cardiovascular safety, it did not demonstrate a statistically significant reduction in major adverse cardiovascular events. In addition, exenatide is not recommended in patients with end-stage renal disease or severe renal impairment, limiting its potential applicability across the full spectrum of CRMM.^102^ In contrast, repaglinide has shown favourable effects on cardiovascular risk factors, potentially through antioxidant activity,^103^ and is considered an appropriate glucose-lowering option for patients with T2D and CKD, including those with advanced renal impairment.^104^ Similarly, sitagliptin provides a cardiovascular-neutral glucose-lowering option, although dose adjustment is required according to renal function.^105,106^ Collectively, these findings suggest that the EAS drug-enrichment signals predominantly implicate established glucose-lowering therapies, while their potential relevance to the broader cardiovascular and renal components of CRMM varies substantially across individual agents.

### Cross-Ancestry Overlap of CRMM Genetic Associations

To assess the extent to which the genetic architecture of CRMM is shared across populations and to identify ancestry-specific signals, we performed cross-ancestry overlap and genetic correlation analyses. Cross-ancestry analyses revealed substantial sharing of CRMM-associated variants across populations. Cross-ancestry overlap identified 719 shared SNPs between EUR and EAS populations and 24 shared SNPs between AFR and EAS, whereas no SNPs were common across all three ancestries under this stringent filtering. Analysis of COJO variants identified three SNPs shared between EUR and EAS populations [<u>rs80196932</u> (*NUS1*), <u>rs1426371</u> (*WSCD2*), and <u>rs2925979</u> (*CMIP*)] highlighting a limited set of robust cross-ancestry signals. When including all GWS SNPs (i.e., without Q_SNP_ exclusion), substantially greater overlap was observed. Specifically, 22 SNPs were shared between EUR and AFR, 1,812 between EUR and EAS, and 4 between AFR and EAS, with 32 SNPs common across all three ancestries. Notably, these shared variants were predominantly mapped to two well-established cardiometabolic loci: *TCF7L2* (8 SNPs) and *FTO* (24 SNPs), underscoring their central role in CRMM susceptibility across diverse populations. These genes are functionally linked to metabolic regulation and neurotrophic signaling pathways (Supplementary Fig. 16).

Cross-ancestry heterogeneity was assessed among 16,203 genome-wide significant (GWS) SNPs. In the overall three-ancestry analysis, 6,993 SNPs (43.16%) showed no statistically significant evidence of heterogeneity after Benjamini-Hochberg correction (adjusted P ≥ 0.05). In pairwise comparisons, the corresponding proportions were 88.20% (14,291 SNPs) for EUR-AFR, 52.70% (8,539 SNPs) for EUR–EAS, and 70.41% (11,408 SNPs) for AFR-EAS. Among the 238 unique COJO SNPs identified across ancestries, 94 (39.50%) showed no statistically significant heterogeneity in the overall three-ancestry analysis. When examined by ancestry-specific COJO sets, 77 of 117 EUR signals (65.81%), none of the four AFR signals, and 20 of 120 EAS signals (16.67%) showed no statistically significant overall heterogeneity. In pairwise comparisons, the proportion of signals without statistically significant heterogeneity varied across ancestry-specific sets. Among EUR COJO signals, these proportions were 86.32% for EUR-AFR, 76.07% for EUR-EAS, and 88.03% for AFR-EAS. Among EAS COJO signals, the corresponding proportions were 95.00%, 17.50%, and 38.33%, respectively. Of the four AFR COJO signals, two (50%) showed no statistically significant heterogeneity for EUR-AFR, while one (25%) showed no statistically significant heterogeneity for each of the other two pairwise comparisons. Overall, these findings indicate that the evidence for cross-ancestry heterogeneity varied by ancestry comparison and by the set of COJO signals examined.

To assess the concordance of our findings with previous CRMM GWAS, GWS variants identified in the present study (11,483 in EUR, 165 in AFR, and 7,731 in EAS) were compared with loci reported by Donker, et al. ^20^, Wang, et al. ^19^, and Lu, et al. ^18^ (Supplementary Fig. 17). We identified two overlapping variants with Donker, et al. ^20^, including <u>rs3768321</u> (*PABPC4*) in EUR and <u>rs1260326</u> (*GCKR*) in EAS. Comparison with Wang, et al. ^19^ showed shared associations at <u>rs9940128</u> (*FTO*), <u>rs4665382</u> (*C2orf16*), <u>rs10938397</u>, <u>rs10244275</u> (*AUSTS2*), and <u>rs4389513</u> (*IGF2BP2*) in EAS, while <u>rs10938397</u> and <u>rs10244275</u> (*AUSTS2*) were also replicated in EUR. No overlapping variants were observed in AFR. Notably, <u>rs10938397</u> and <u>rs10244275</u> (*AUSTS2*) were common to both EUR and EAS populations and were previously reported by Wang, et al. ^19^. Comparison with Lu, et al. ^18^ identified <u>rs1421085</u> (*FTO*) as a shared locus across AFR and EAS cohorts, together with <u>rs1519480</u> (*BDNF-AS*) in AFR. In addition, 39 and 26 previously reported loci overlapped with the EUR and EAS CRMM GWAS results, respectively, further supporting the robustness and reproducibility of the identified associations.

### Mapping of CRMM-associated variants to individual component conditions

To characterise the contribution of individual cardiometabolic conditions to the genetic architecture of CRMM, we assessed the overlap between CRMM-associated variants and GWS (*P*<5 × 10^-8^) variants for HP, IHD, HF, T2D, and CKD, separately across EUR, AFR, and EAS ancestries. The analysis considered both the ancestry-specific CRMM GWS SNP sets and the conditionally independent SNPs identified through COJO. For each component and ancestry, we quantified the number of overlapping variants relative to the total number of CRMM GWS or COJO SNPs (Supplementary Table 42).

Among the 11,484 EUR CRMM GWS SNPs, the largest overlaps were observed with T2D (9,290; 80.9%) and HP (8,187; 71.3%), followed by IHD (3,452; 30.1%), HF (1,621; 14.1%), and CKD (1,592; 13.9%). Of the 165 AFR CRMM GWS SNPs, 36 (21.8%) overlapped with T2D and 31 (18.8%) with HP, while overlap with CKD was limited to two SNPs and no exact overlap was observed with IHD or HF. In EAS, 2,215 of 7,732 CRMM GWS SNPs (28.6%) overlapped with T2D, whereas 21 (0.3%) overlapped with HYP; no exact overlap was observed with IHD, HF, or CKD.A broadly similar pattern was observed for the conditionally independent CRMM SNPs. Among the 148 EUR COJO SNPs, 113 (76.4%) overlapped with T2D GWS variants and 91 (61.5%) with HP, followed by IHD (48; 32.4%), HF (25; 16.9%), and CKD (12; 8.1%). Among the 16 AFR COJO SNPs, six (37.5%) overlapped with T2D, three (18.8%) with HP, and one (6.3%) with CKD, with no overlap observed for IHD or HF. Of the 139 EAS COJO SNPs, 46 (33.1%) overlapped with T2D and one (0.7%) with HYP, with no exact overlap observed for the remaining components.

## Supplementary Figures

**Supplementary Fig. 1.**
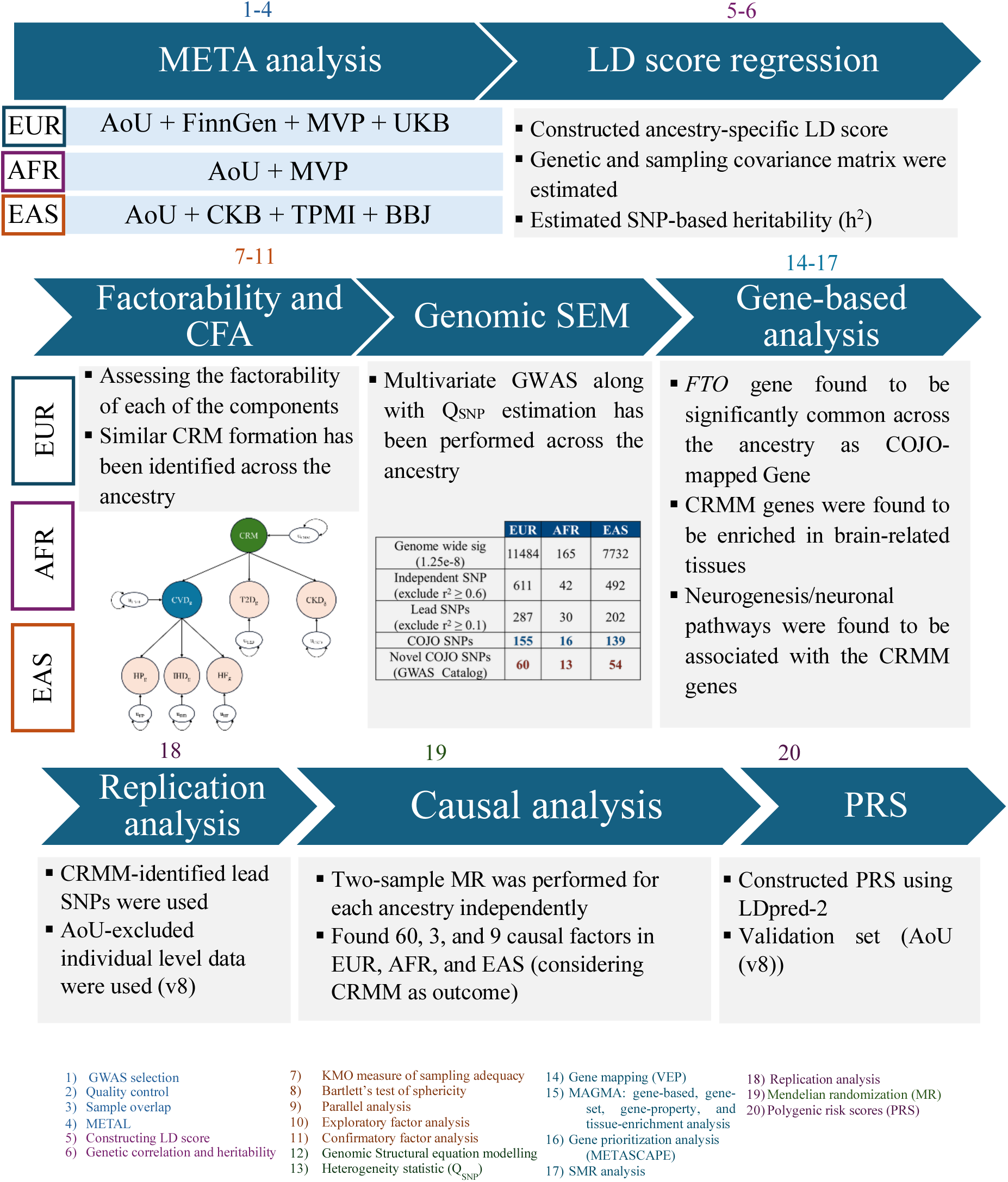
Schematic overview of the study workflow for performing the CRMM multivariate GWAS. Number listed above the headings indicate which analyses occurred during each stage of the workflow.

**Supplementary Fig. 2.**
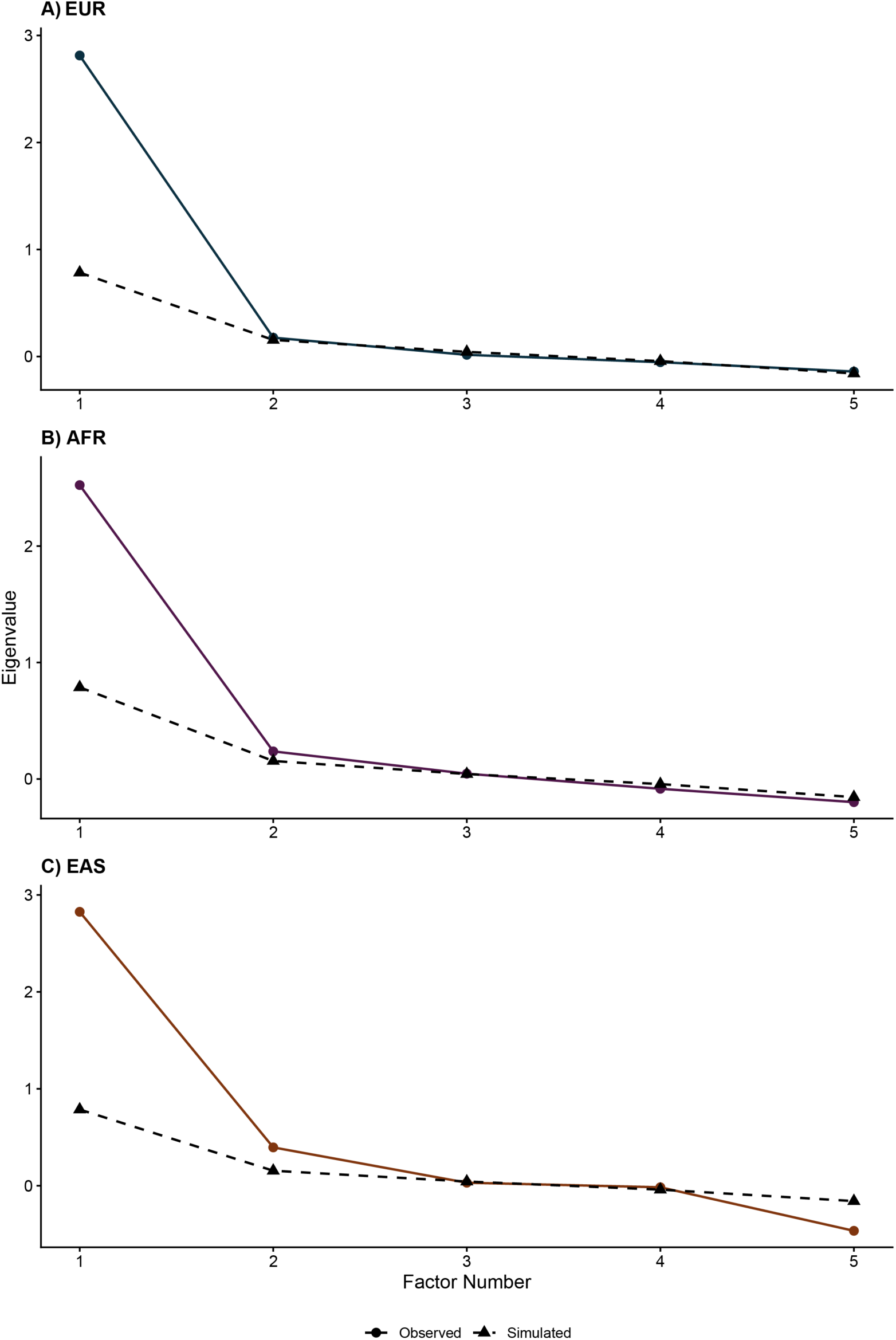
Scree plots for determining the number of latent factors. Scree plots derived from parallel analysis to infer the number of underlying latent factors across **A)** European (EUR), **B)** African (AFR), and **C)** East Asian (EAS) ancestries (top to bottom). The x-axis represents the number of factors, and the y-axis represents eigenvalues. The solid line corresponds to observed data, whereas the dashed red line represents simulated data.

**Supplementary Fig. 3.**
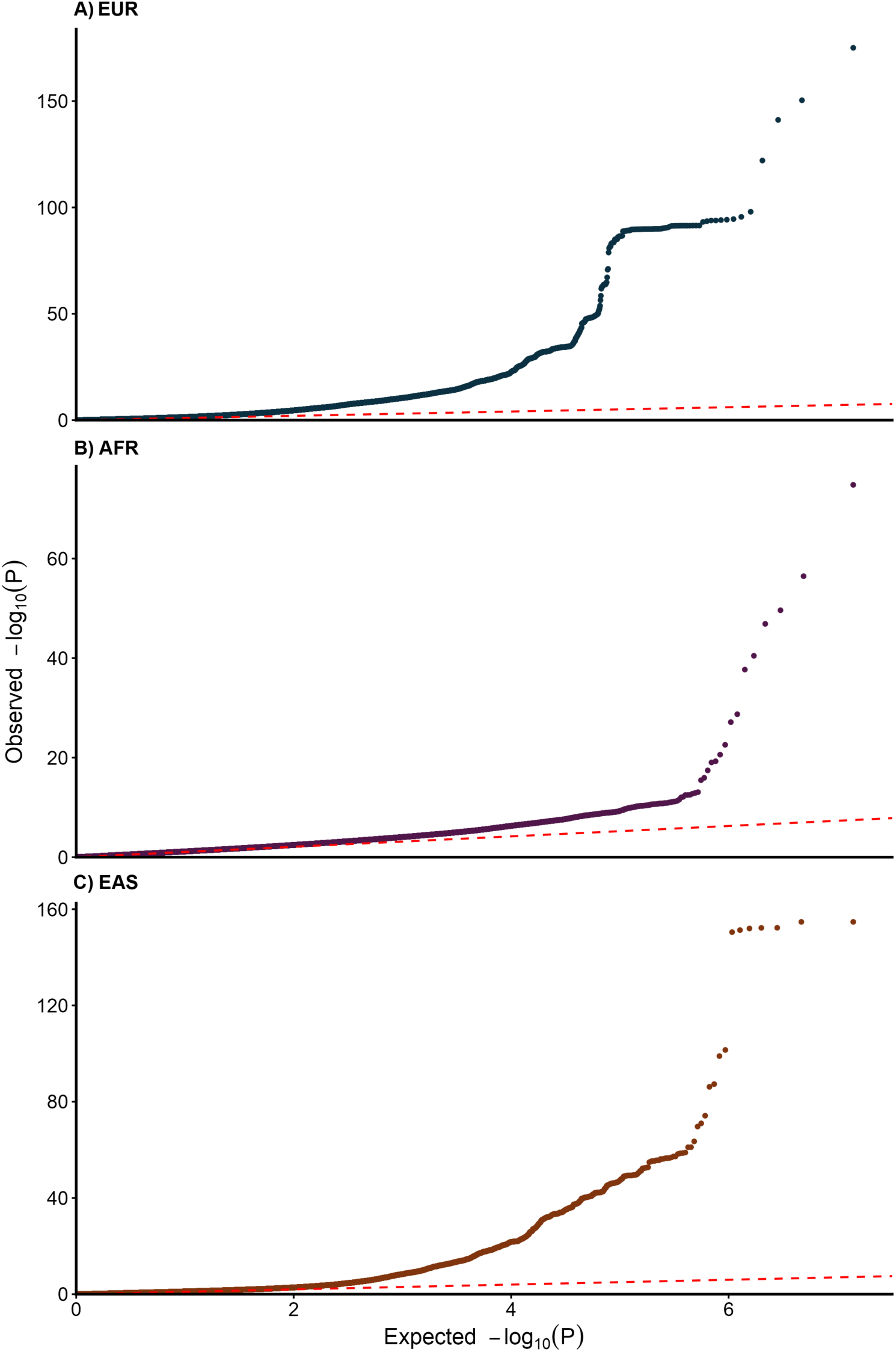
Q-Q plots of CRMM multivariate GWAS after exclusion of Q_SNP_-associated variants. Quantile-quantile (Q-Q) plots showing observed versus expected uncorrected −log_10_ (P) values for CRMM multivariate GWAS across **A)** European (EAS), **B)** African (AFR), and **C)** East Asian (EAS) ancestries (top to bottom). The x-axis represents expected values, and the y-axis represents observed values.

**Supplementary Fig. 4.**
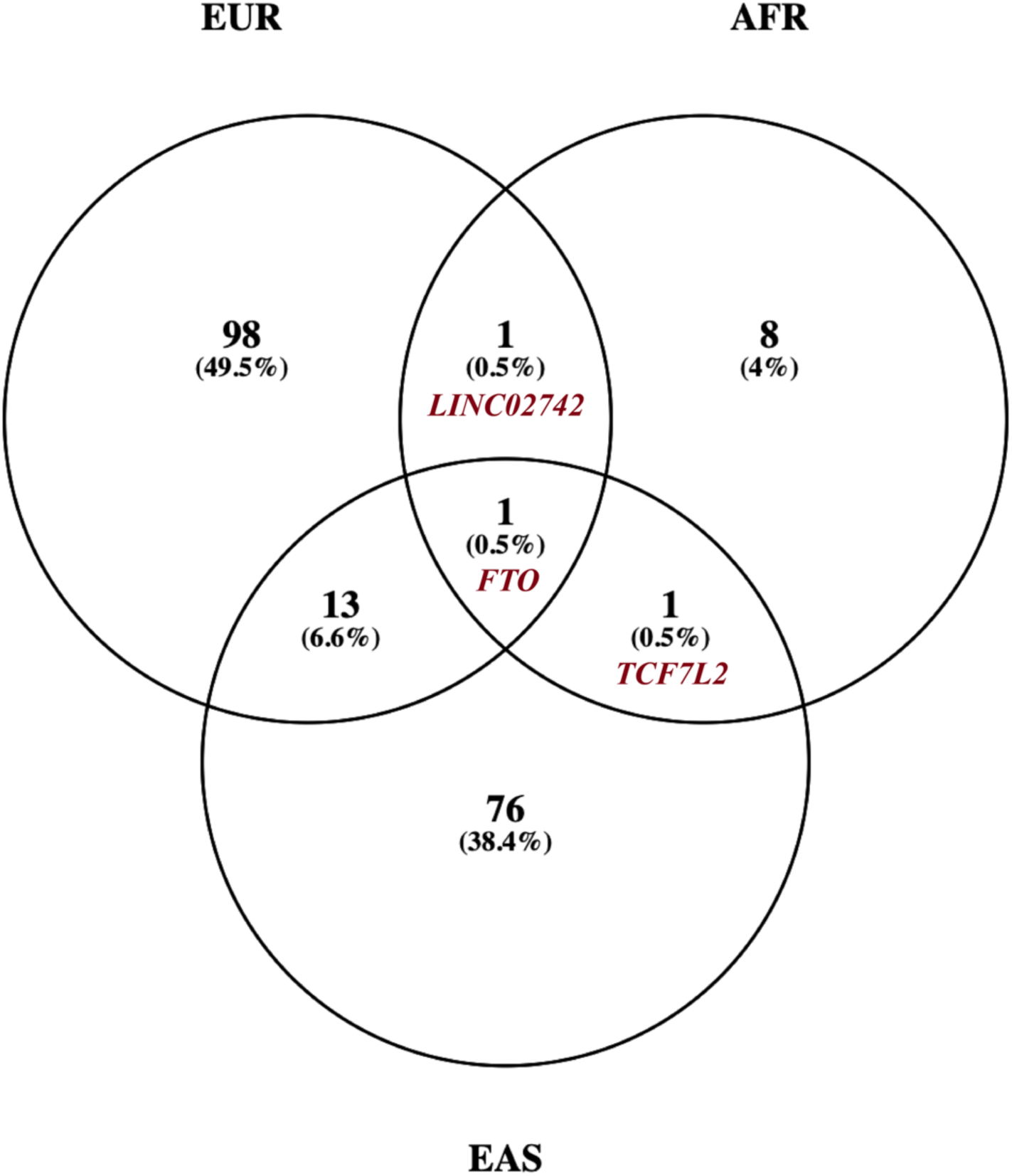
Genes identified through COJO mapping across European (EUR), African (AFR), and East Asian (EAS) ancestries. The Venn diagram illustrates genes mapped from COJO SNPs using VEP annotation, with the intersections representing the number of overlapping genes across ancestries. The numbers, along with the corresponding percentages (in parentheses), are reported.

**Supplementary Fig. 5.**
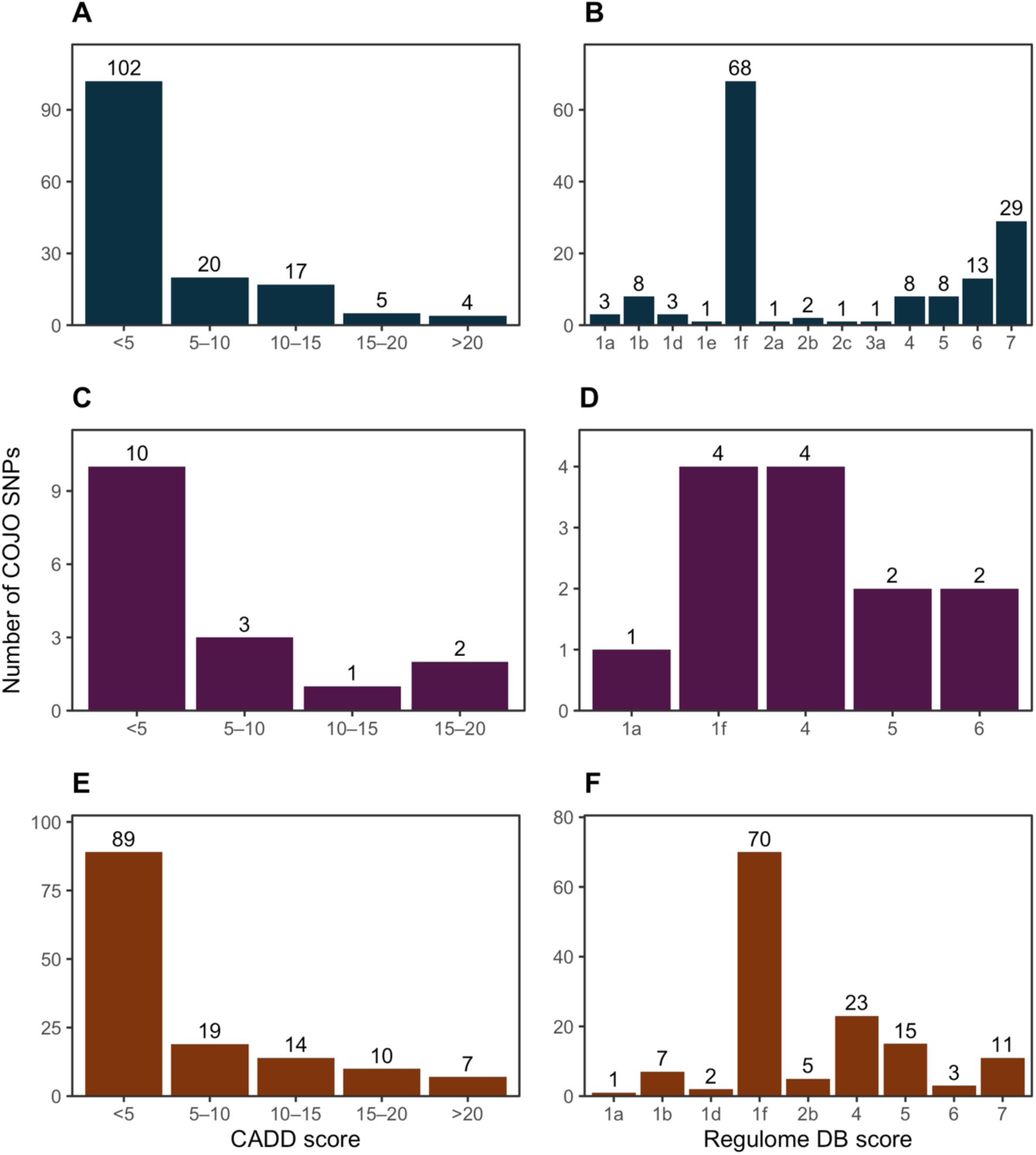
Distribution of functional annotation scores for COJO-prioritized SNPs across European, African, and East Asian populations. Bar plots showing the distribution of functional annotation scores for conditionally independent (COJO) SNPs identified in the CRMM GWAS across European (EUR), African (AFR), and East Asian (EAS) populations. Panels **A**, **C**, and **E** present the distribution of CADD scores for EUR, AFR, and EAS populations, respectively. Panels **B**, **D**, and **F** present the distribution of RegulomeDB scores for EUR, AFR, and EAS populations, respectively.

**Supplementary Fig. 6.**
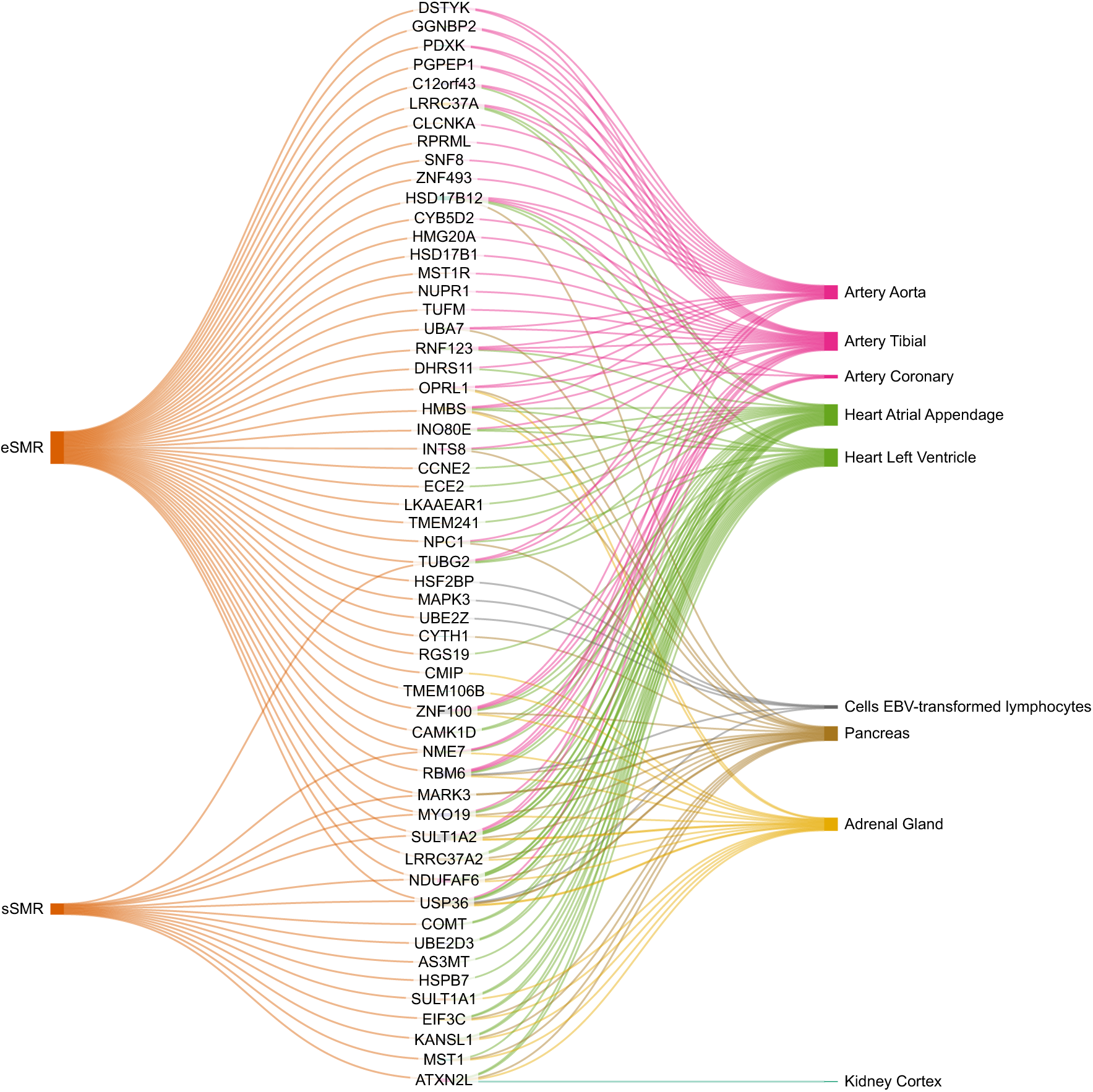
Prioritized genes identified through SMR analysis using eQTL and sQTL data. Sankey plot illustrating genes prioritized by Summary-data-based Mendelian Randomization (SMR) analysis based on expression quantitative trait loci (eQTL) and splicing quantitative trait loci (sQTL) datasets. Only genes that were significant in the SMR analysis and passed HEIDI filtering were retained.

**Supplementary Fig. 7.**
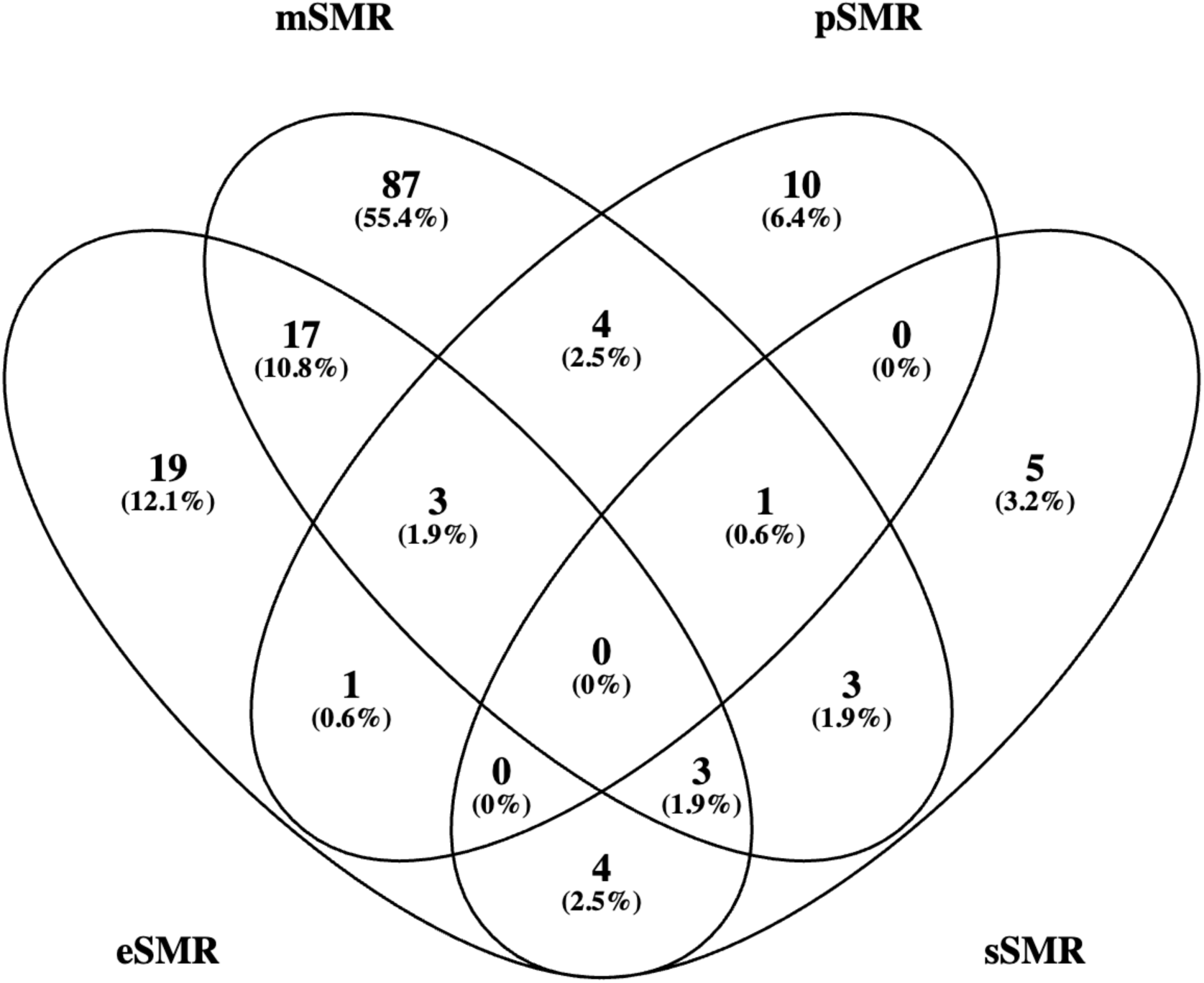
Overlap of prioritized genes identified across different SMR analyses. mSMR = methylation SMR, pSMR = protein SMR, eSMR = expression SMR, sSMR = splicing SMR.

**Supplementary Fig. 8.**
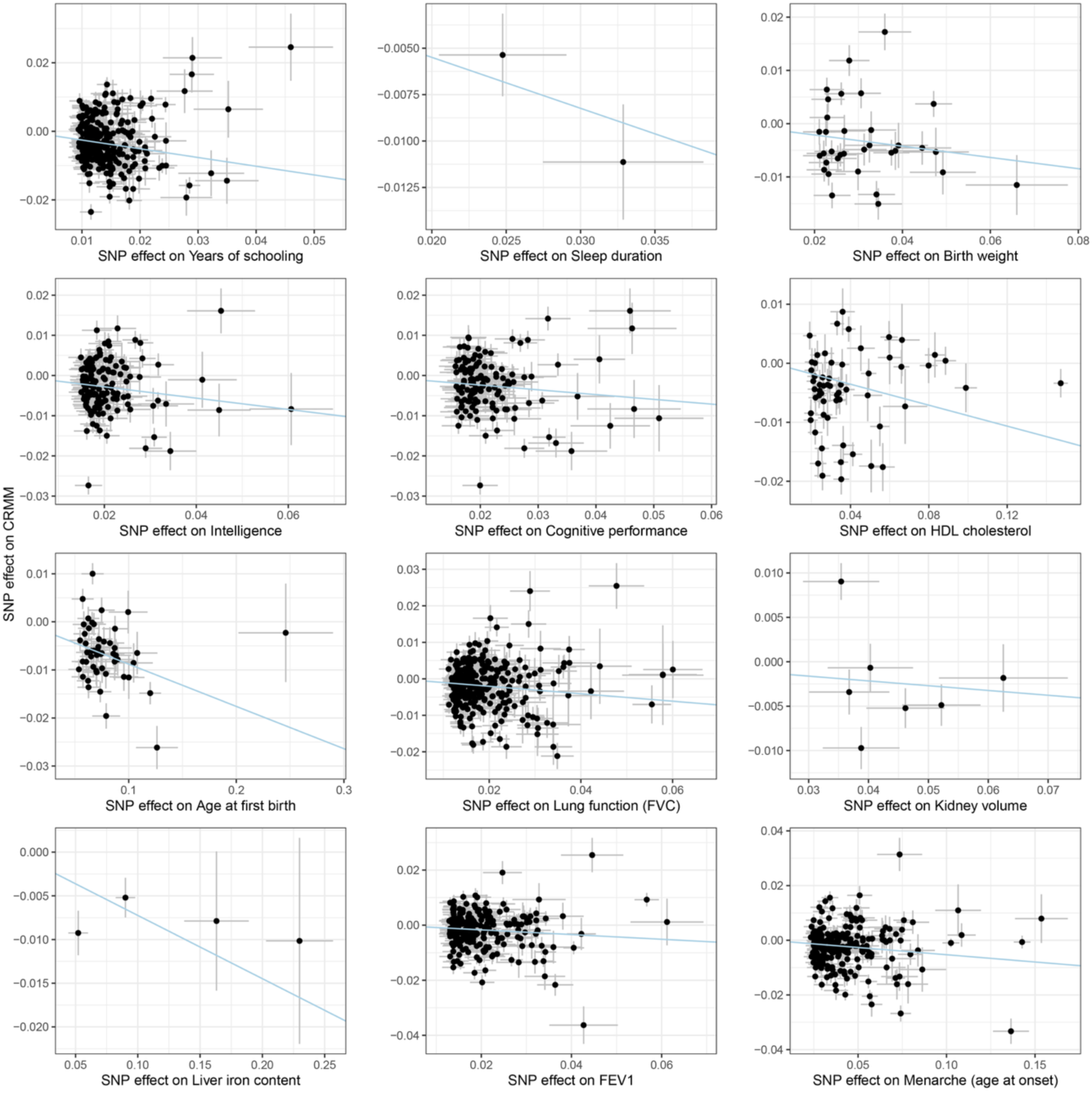
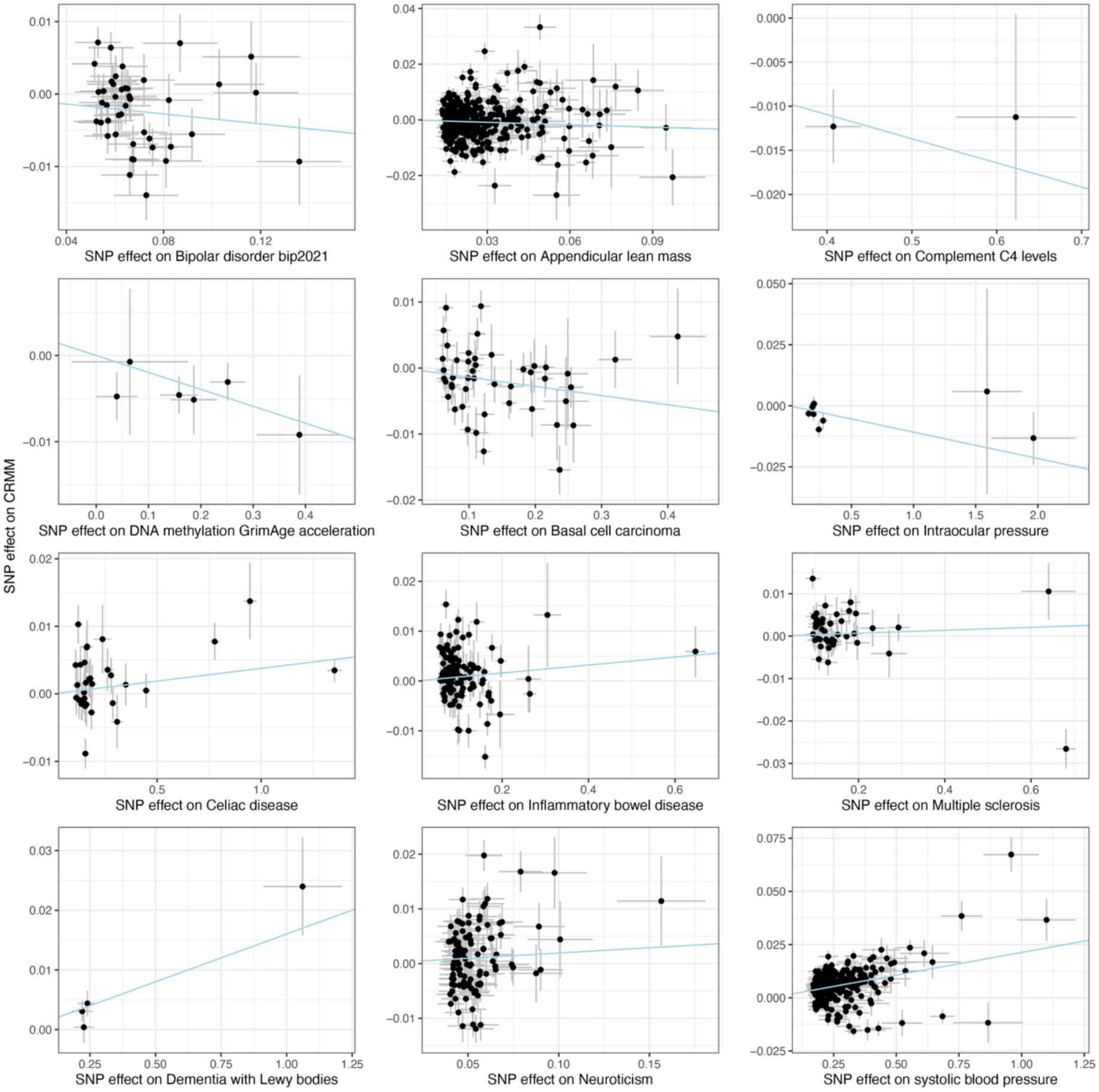
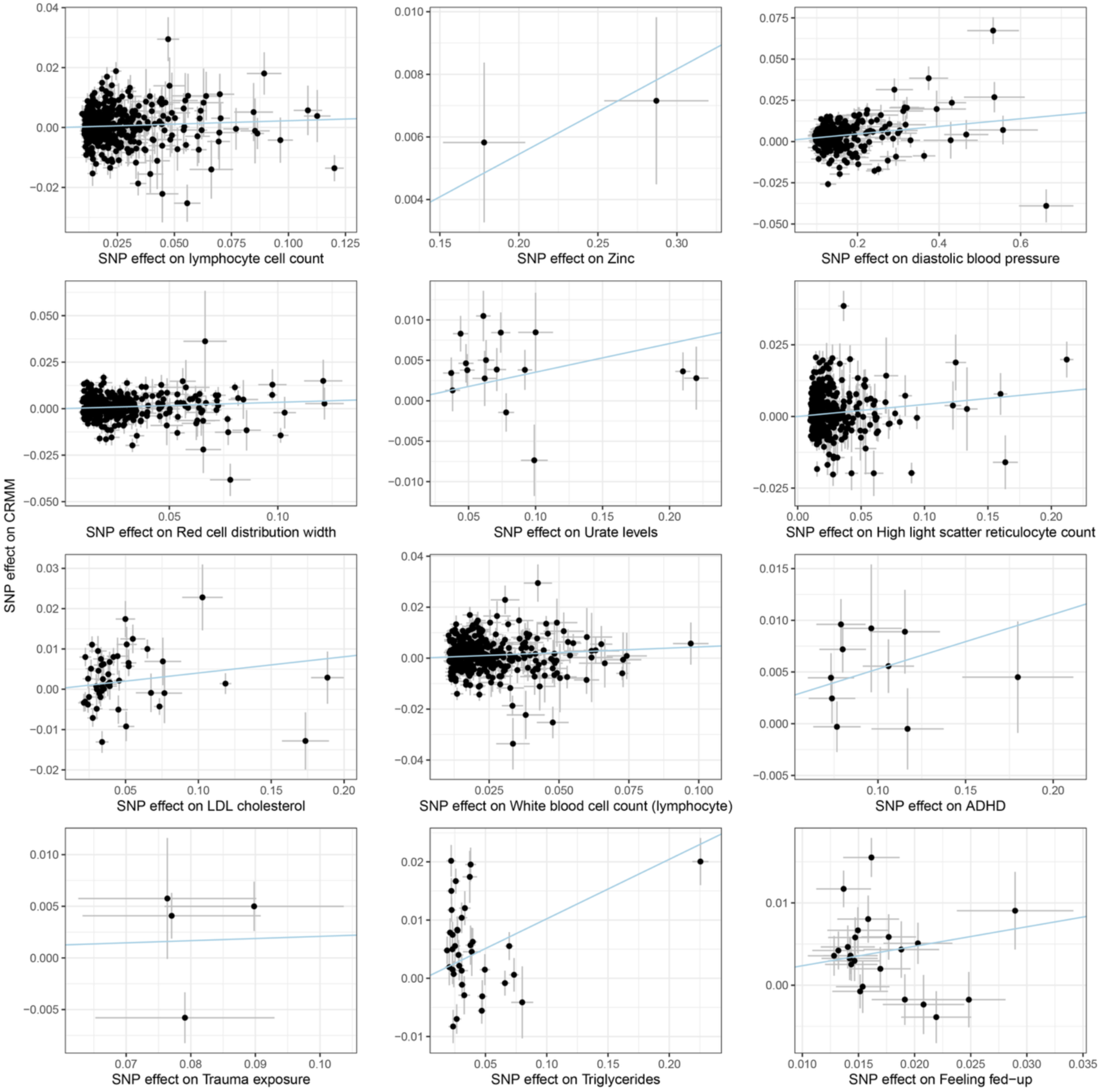
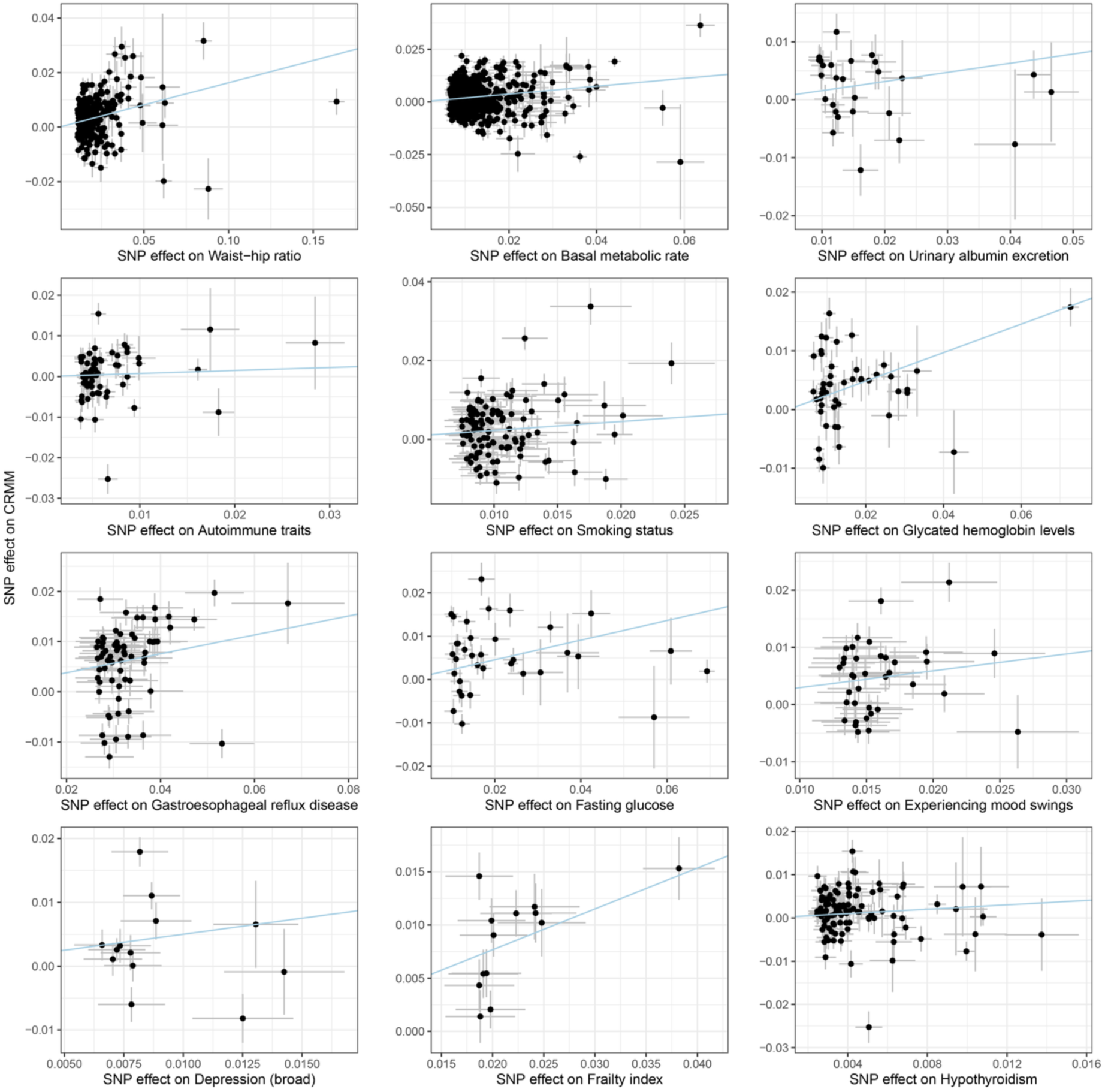
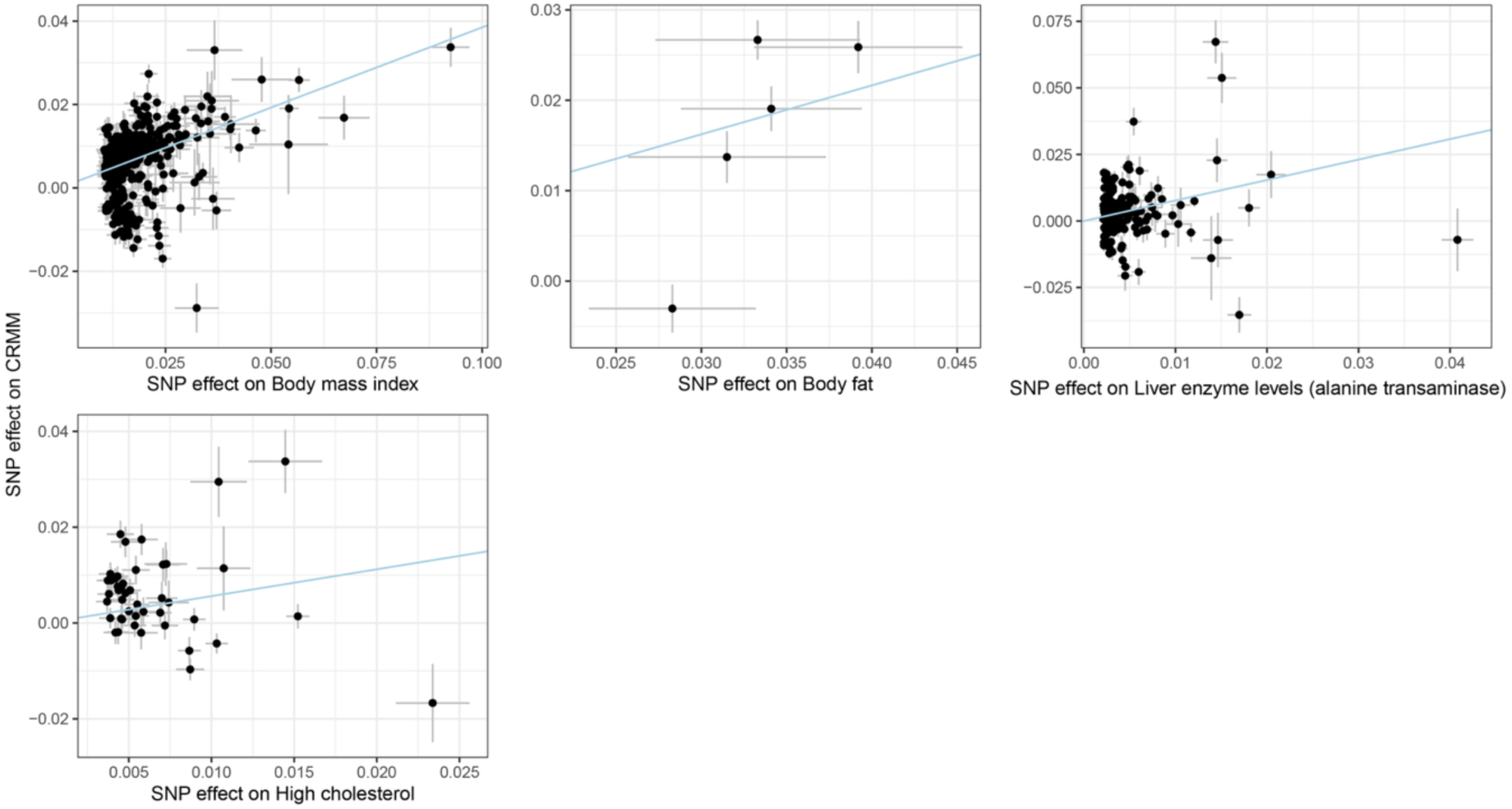
Mendelian randomization scatter plots showing genetic associations of exposures with CRMM in Europeans. Scatter plots showing SNP-specific effects of genetic instruments on each exposure (x-axis) against their corresponding effects on CRMM (y-axis) in the European ancestry population. Each point represents an instrumental variant. The blue line denotes the inverse-variance weighted (IVW) causal estimate.

**Supplementary Fig. 9.**
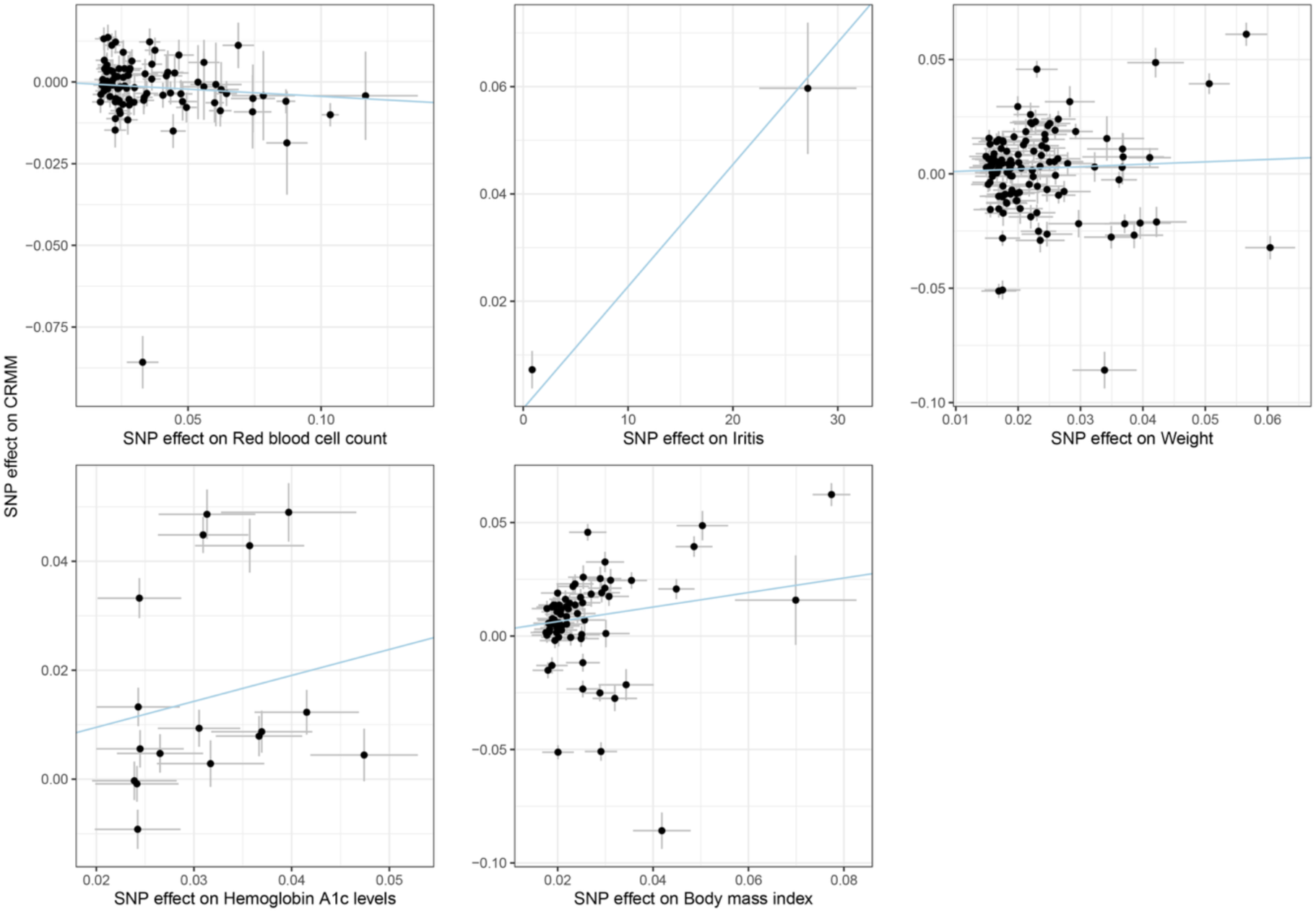
Scatter plots of SNP-specific associations between exposures and CRMM in the East Asian population. Scatter plots showing SNP-specific effects of genetic instruments on each exposure (x-axis) against their corresponding effects on CRMM (y-axis) in the East Asian ancestry population. Each point represents an instrumental variant. The blue line denotes the inverse-variance weighted (IVW) causal estimate.

**Supplementary Fig. 10.**
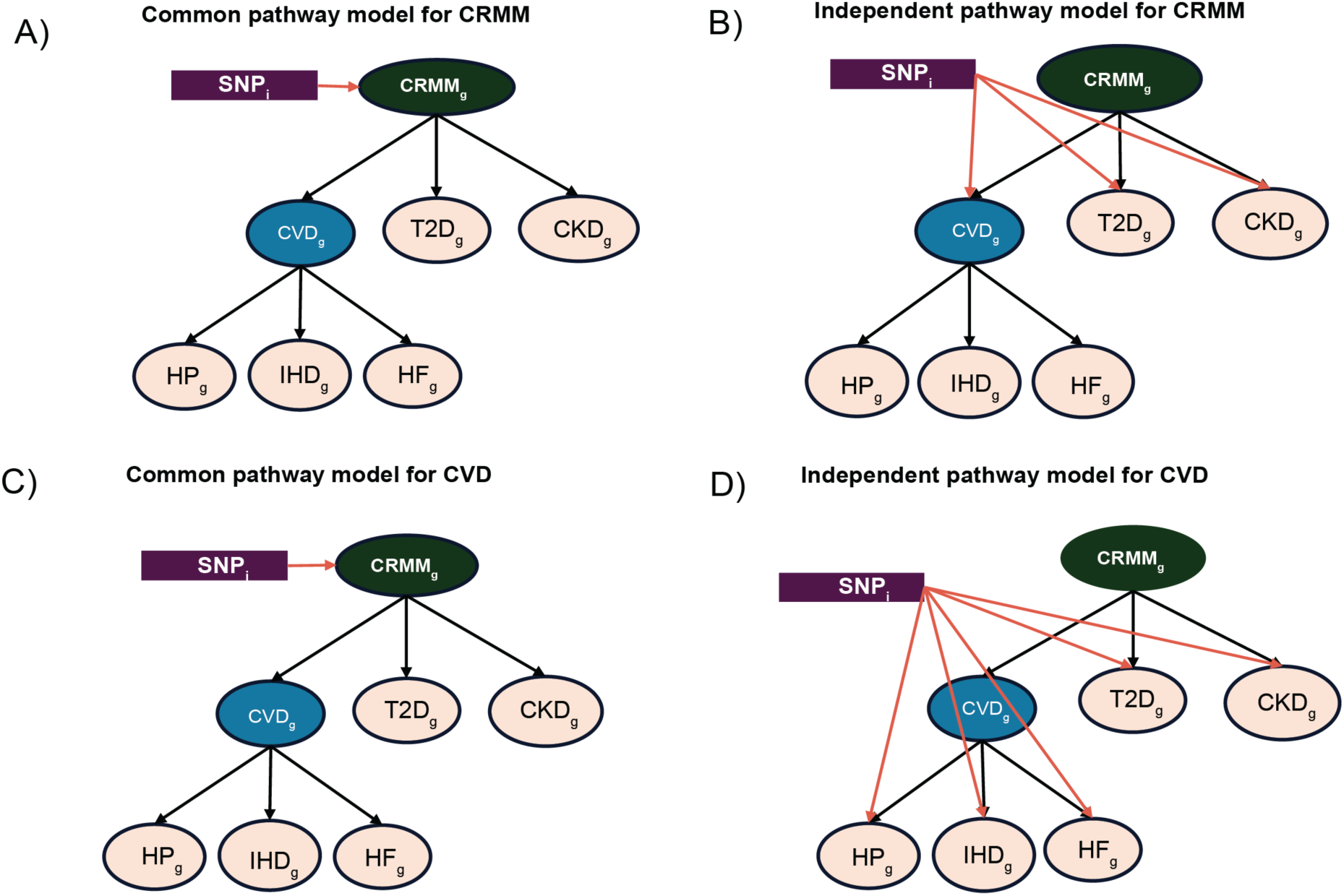
Common and independent pathway models for Q_SNP_ estimation. Schematic representation of the common and independent pathway models used to estimate Q_SNP_ for CRMM (**A** and **B**) and CVD (**C** and **D**). HP: hypertension; HF: heart failure; IHD: ischemic heart disease; T2D: type 2 diabetes; CKD: chronic kidney disease; CRMM: cardio-renal-metabolic multimorbidity; u: residual variance not explained by the latent factor; subscript g: indicates that the model is based on genetic covariances.

**Supplementary Fig. 11.**
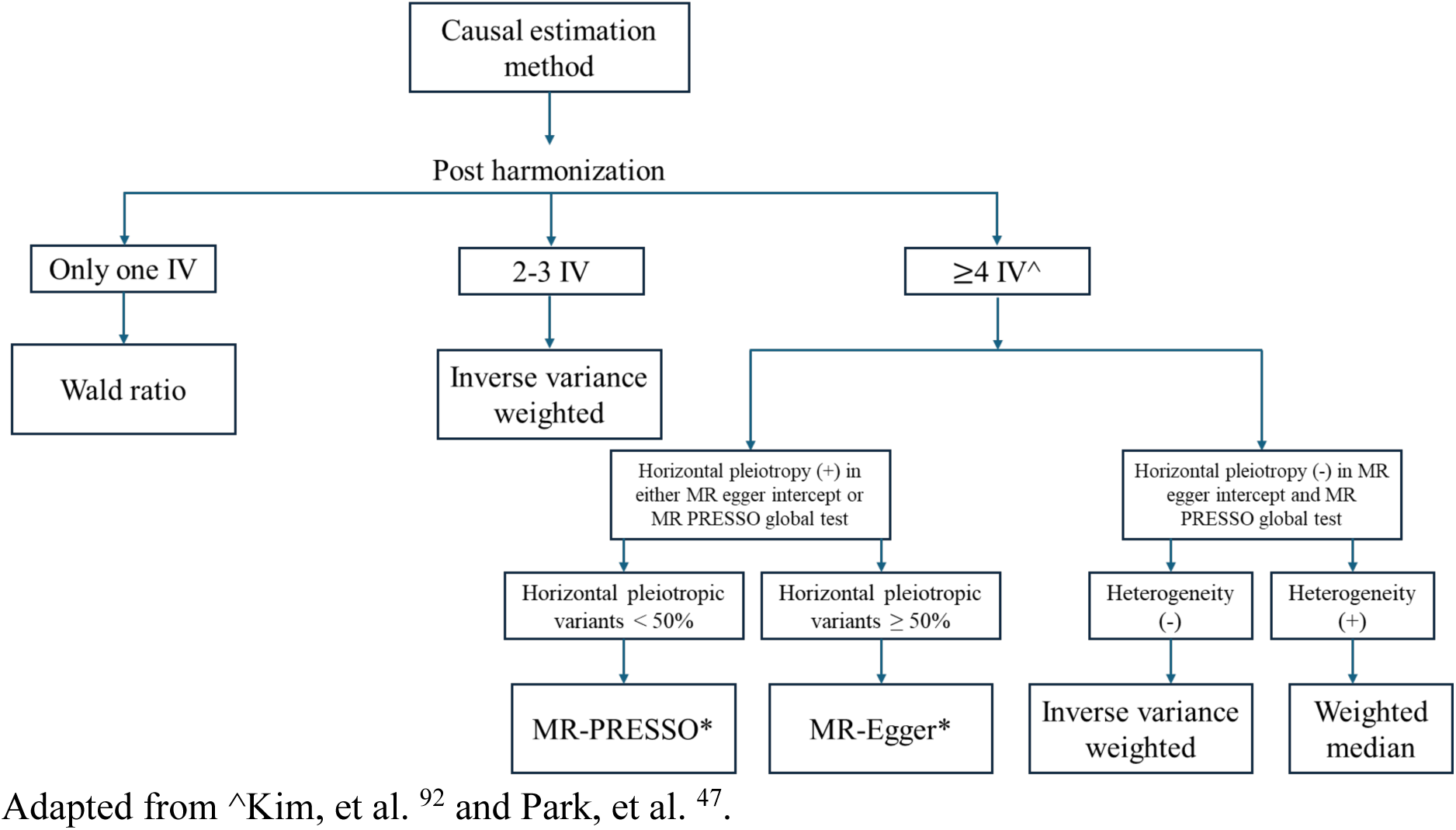
Decision Framework for Selecting Optimal Causal Estimation Methods. The flowchart below illustrates the MR framework used to determine the most appropriate causal estimates. *If no horizontal pleiotropic outliers were detected and a corrected MR-PRESSO estimate could not be generated, we used MR-Egger estimates instead; IV: Instrumental Variable. Adapted from ^Kim, et al. ^92^ and Park, et al. ^47^.

**Supplementary Fig. 12.**
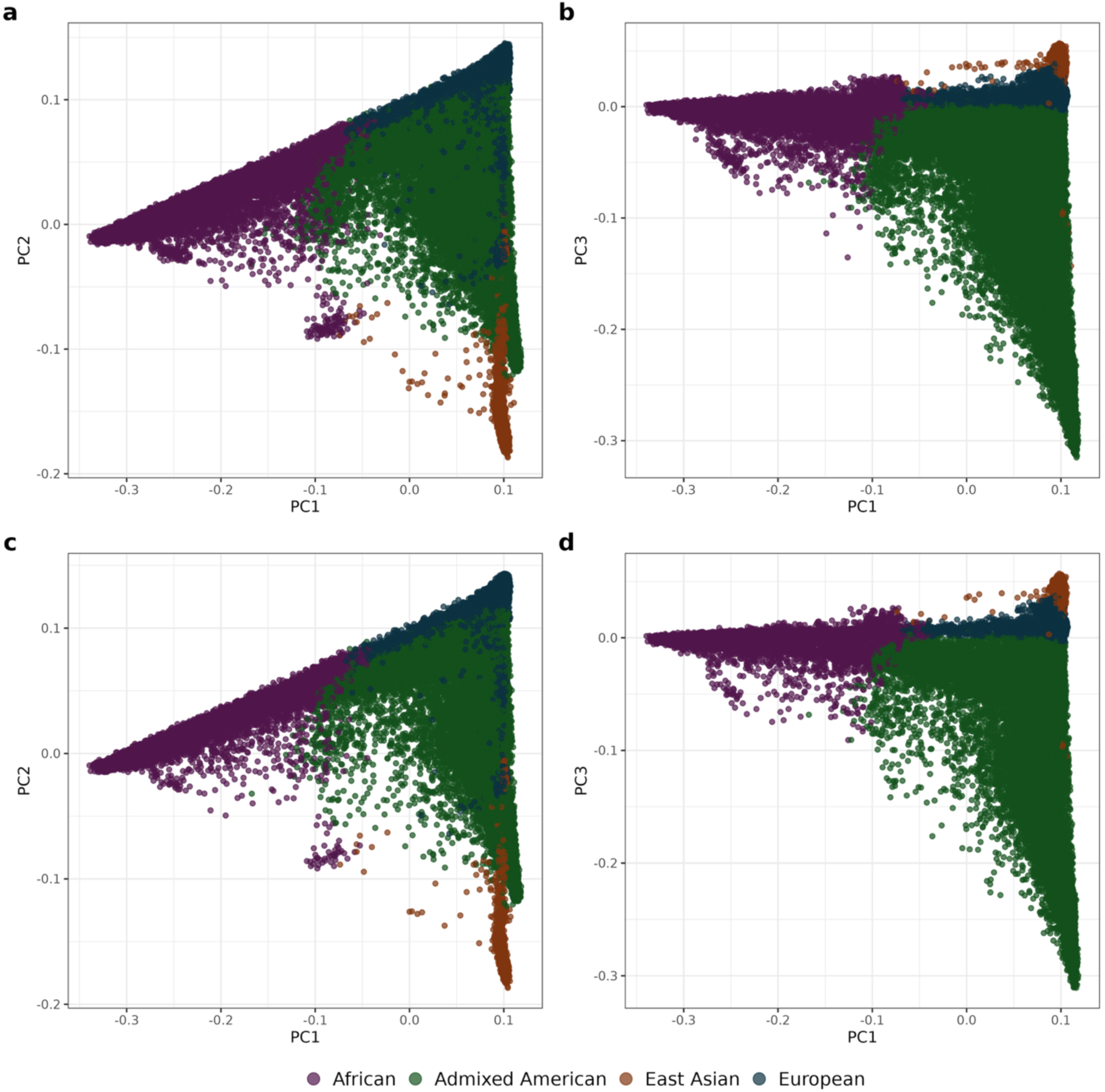
Population Structure of the All of Us v8 and Replication Cohorts. PCA plots showing population structure in the All of Us cohorts. Panels **a** and **b** represent the AoU v8 discovery cohort, and panels **c** and **d** represent the AoU replication cohort. Each point denotes an individual and is coloured by genetic ancestry.

**Supplementary Fig. 13.**
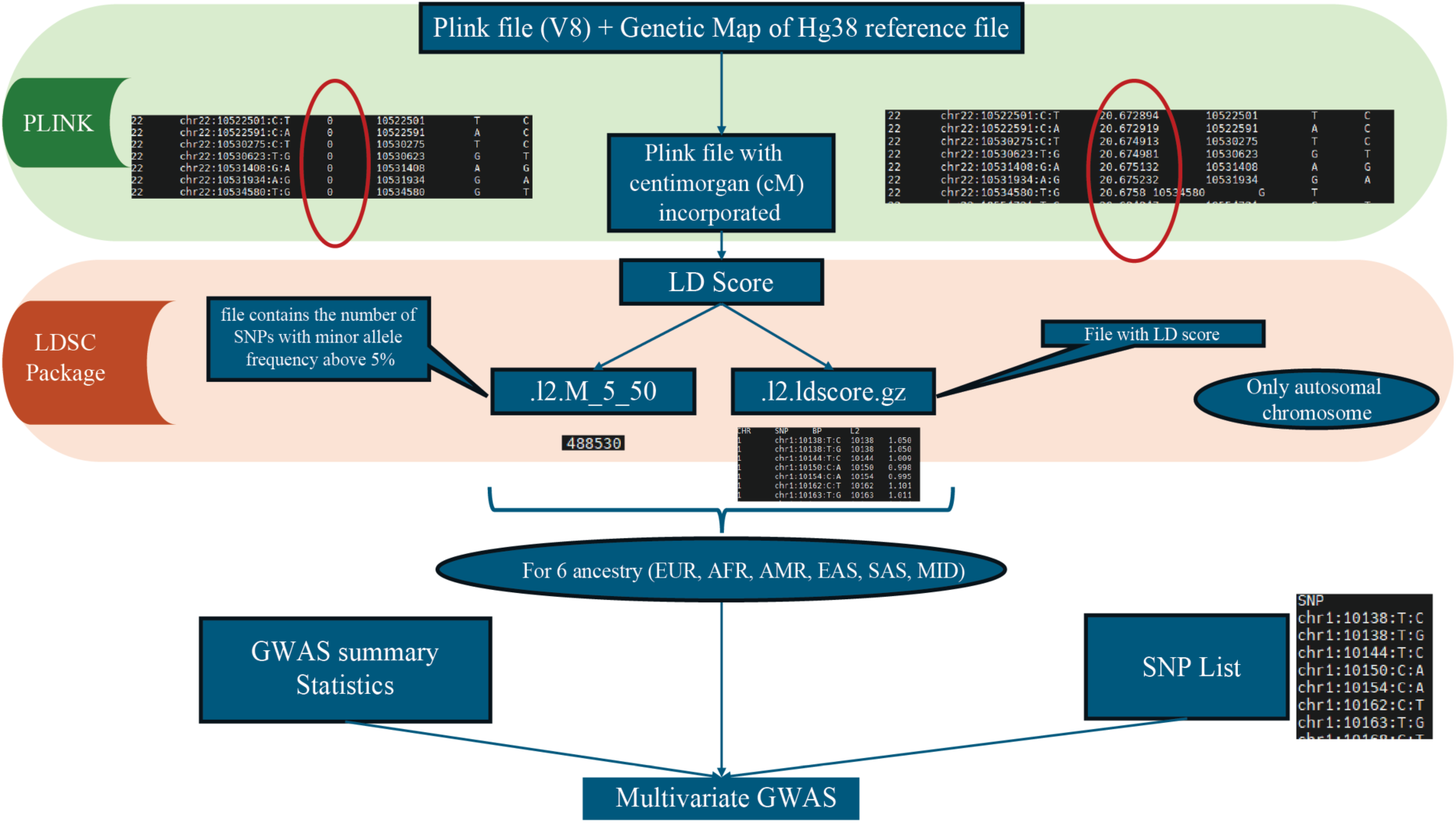
Workflow for PLINK File preprocessing and LD score regression

**Supplementary Fig. 14.**
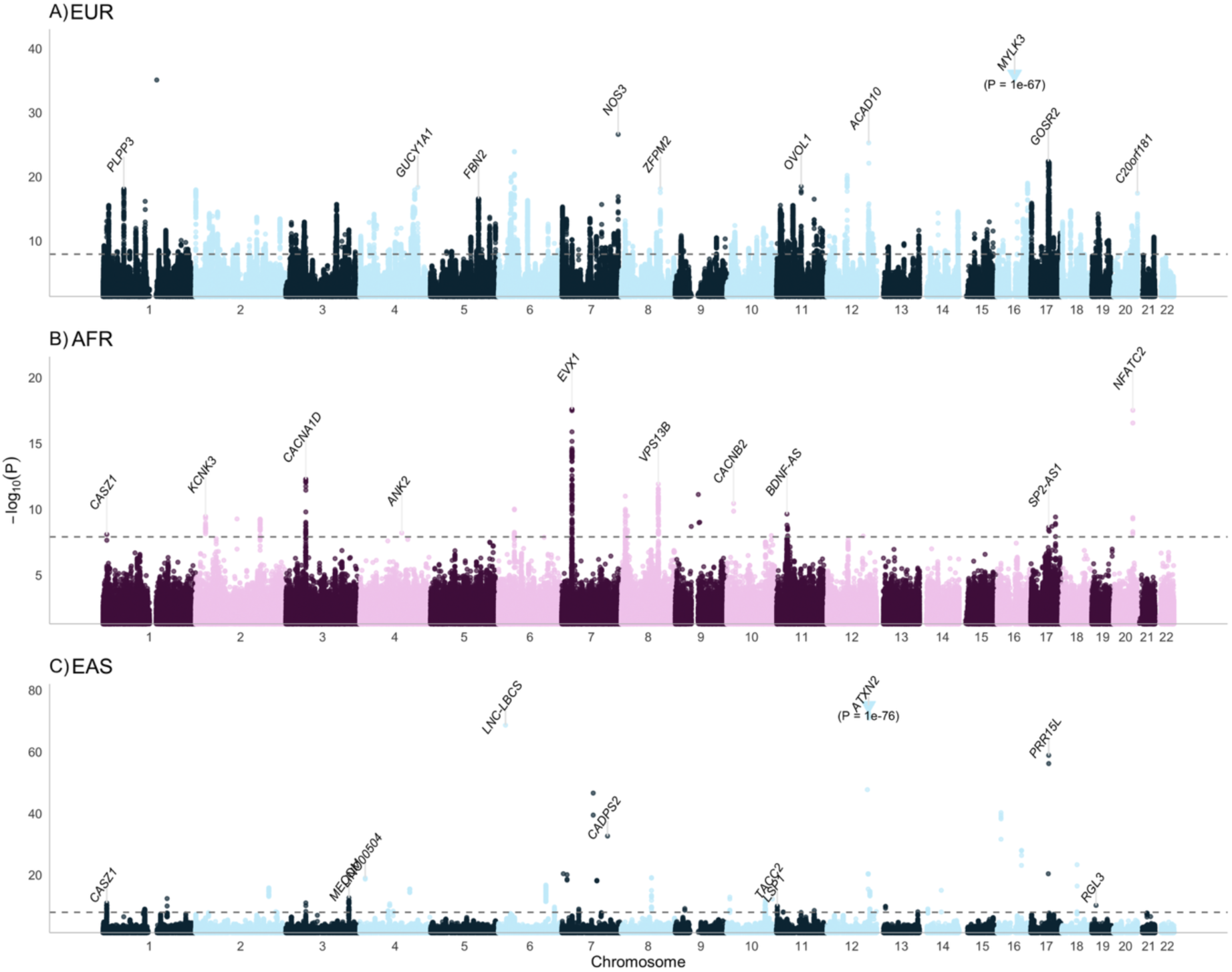
Manhattan plots of genome-wide significant loci for CVD across ancestries. Manhattan plots showing genome-wide association results for CVD in **A)** European (EUR), **B)** African (AFR), and **C)** East Asian (EAS) ancestries (top to bottom). The horizontal dashed line indicates the genome-wide significance threshold (*P* = 1.25 × 10^-8^). The top 10 lead SNPs are highlighted and annotated with their corresponding mapped genes.

**Supplementary Fig. 15.**
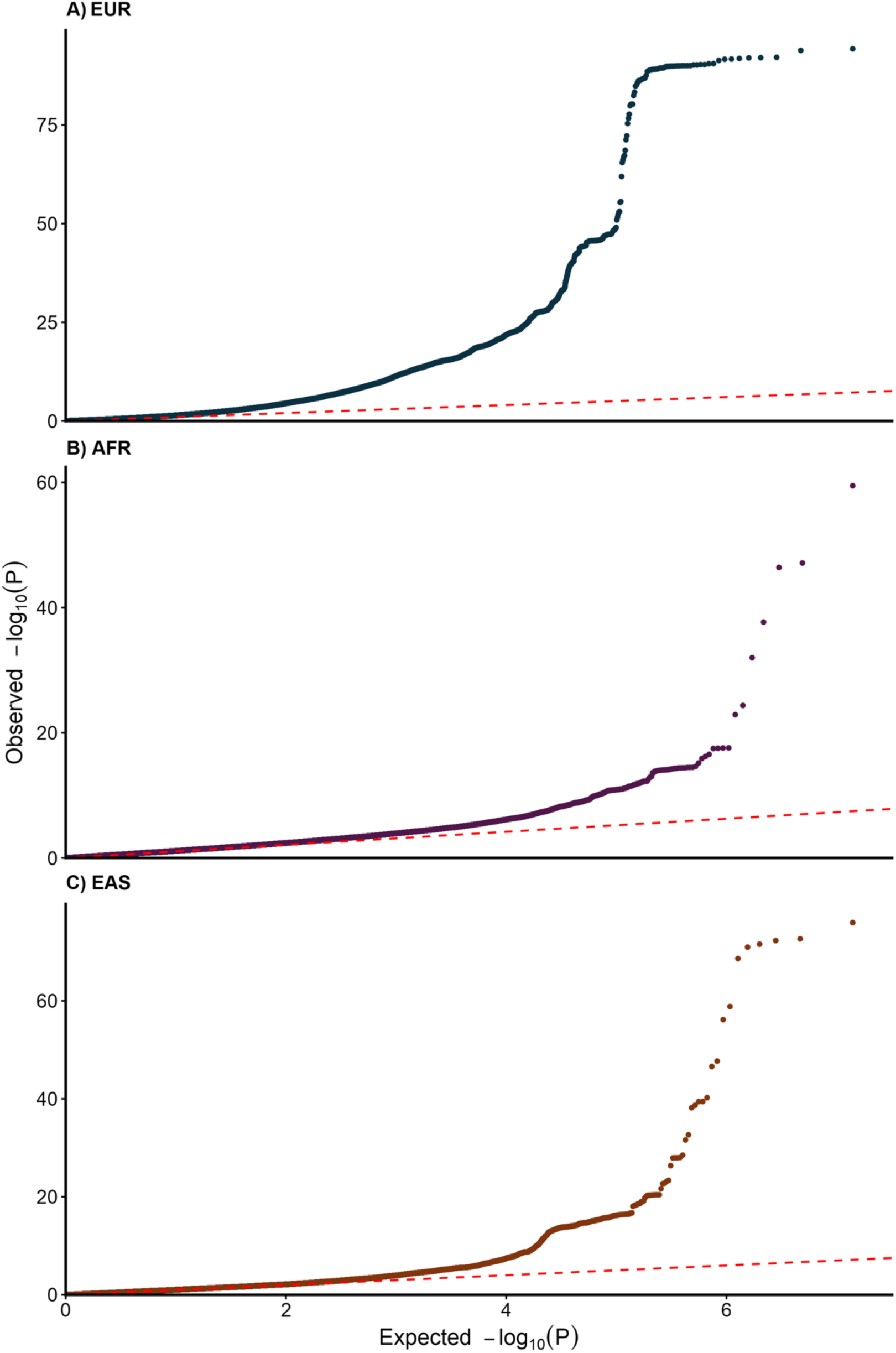
Q-Q plots of CVD multivariate GWAS after exclusion of Q_SNP_-associated variants. Quantile-quantile (Q-Q) plots showing observed versus expected uncorrected −log_10_ (P) values for CVD multivariate GWAS across **A)** European (EUR), **B)** African (AFR), and **C)** East Asian (EAS) ancestries (top to bottom). The x-axis represents expected values, and the y-axis represents observed values.

**Supplementary Fig. 16.**
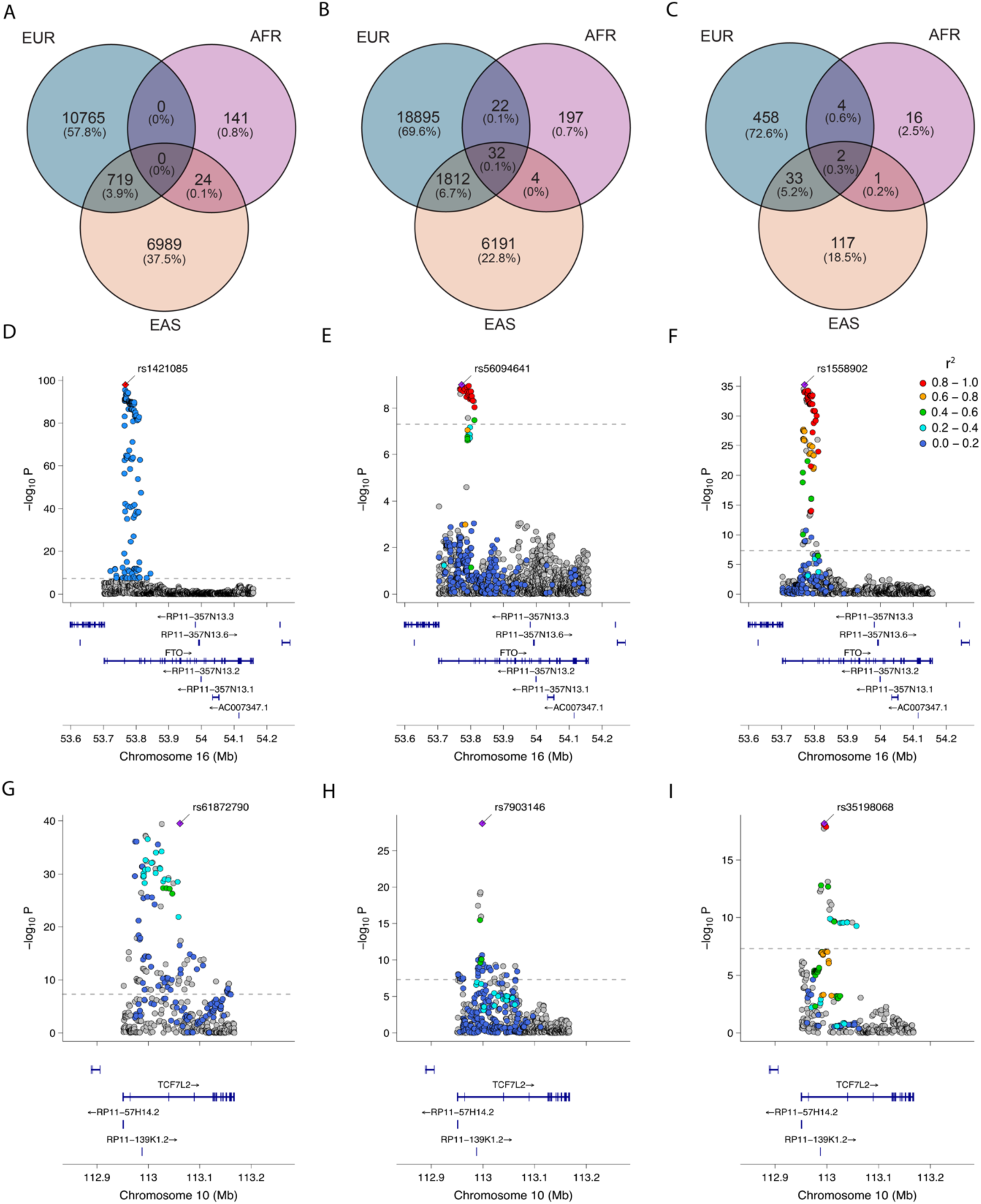
Shared genetic signals across European, African, and East Asian ancestries. **A**, Venn diagram illustrating Genome-wide significant (GWS) SNPs identified from the CRMM multivariate GWAS following Q_SNP_ filtering. **B,** Venn diagram illustrating GWS SNPs from the CRMM multivariate GWAS without Q_SNP_ filtering. **C,** Venn diagram illustrating statistically significant genes identified across ancestries using MAGMA gene-based analysis. **D-F**, Regional association (locus) plots for the *FTO* gene across European (EUR), African (AFR), and East Asian (EAS) ancestries (left to right). **G-I**, Regional association (locus) plots for the *TCF7L2* gene across EUR, AFR, and EAS ancestries (left to right).

**Supplementary Fig. 17.**
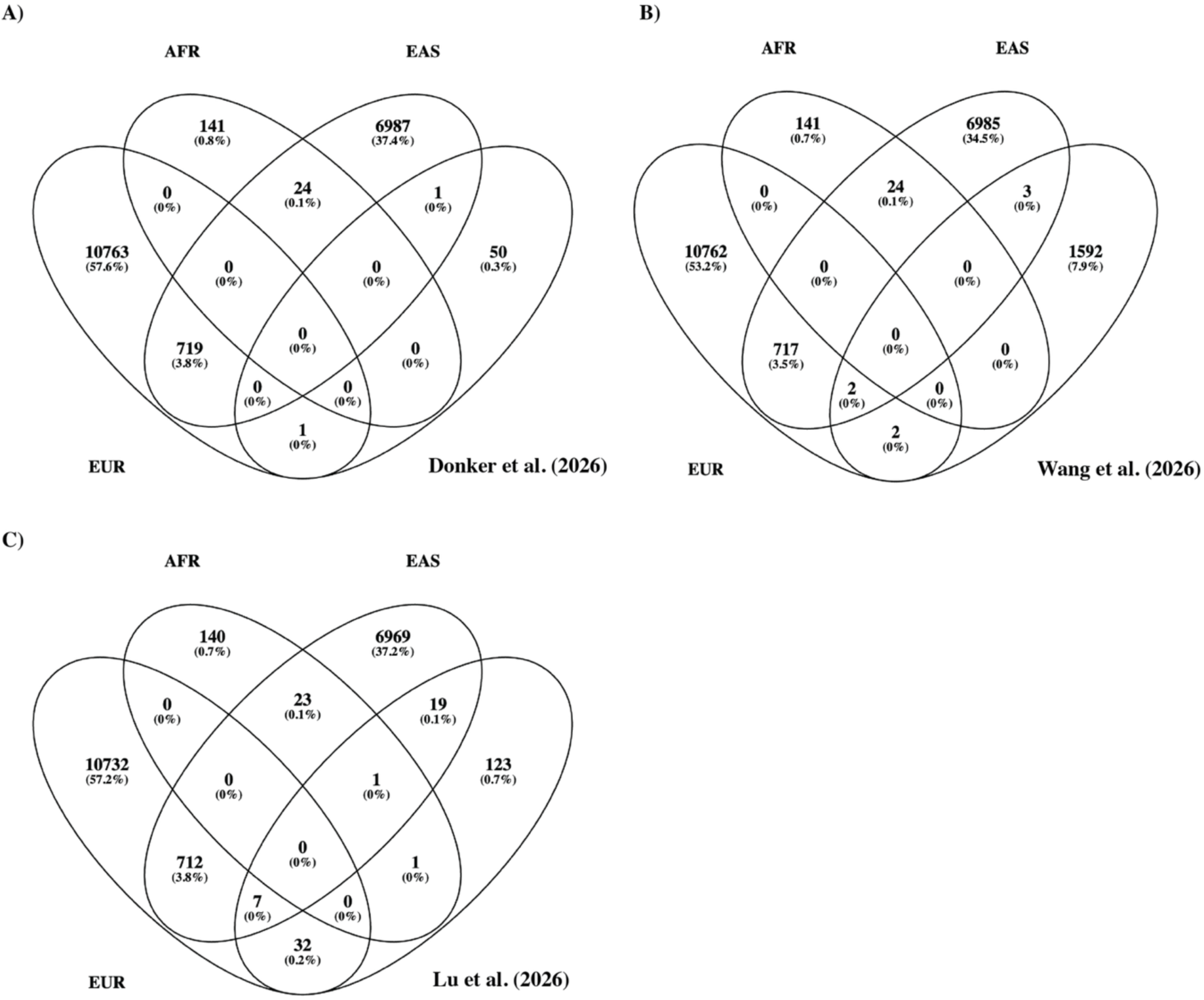
Overlap of genome-wide significant (GWS) SNPs identified in the present CRMM GWAS with previously reported CRMM GWAS loci across ancestries. Venn diagrams illustrating the overlap between GWS SNPs identified in the current chronic renal-metabolic multimorbidity (CRMM) GWAS and loci reported in previous studies: (A) Donker, et al. (2026), (B) Wang, et al. (2026), and (C) Lu, et al. (2026). Overlap was assessed separately for each ancestry using the GWS SNPs identified in the present study, comprising 11,483 SNPs in EUR, 165 SNPs in AFR, and 7,731 SNPs in EAS. Numbers within the Venn diagrams indicate shared and ancestry-specific GWS SNPs between the current study and each published CRMM GWAS.

Supplementary Reference

## Notes

### Competing Interest Statement

The authors have declared no competing interest.

### Author Declarations

This study utilized data from FinnGen, UK Biobank (UKB), Million Veteran Program (MVP), China Kadoorie Biobank (CKB), Taiwan Precision Medicine Initiative (TPMI), BioBank Japan (BBJ), and All of Us (AoU). Most analyses were conducted using publicly available summary statistics. However, the replication analyses performed using AoU required access to individual level data through the AoU Researcher Workbench Controlled Tier. Access was obtained after completion of the required Data User Code of Conduct and Data Use Registration Agreement (DURA), in accordance with AoU data access policies. All procedures adhered fully to the data access requirements and governance policies established by each respective biobank.

### Summary of Updates

Abstract section revised to improve clarity; all figures revised and updated; additional figures added to the Supplementary Note; final paragraph of the Introduction revised to improve readability; Discussion section substantially rewritten to improve clarity and interpretation of findings; Supplementary Note completed and expanded with additional methodological details and supporting results.

## References

1. Marassi, M. & Fadini, G.P. The cardio-renal-metabolic connection: a review of the evidence. Cardiovascular Diabetology 22(2023).

2. Seferovic, P.M. et al. Type 2 diabetes mellitus and heart failure: a position statement from the Heart Failure Association of the European Society of Cardiology. European Journal of Heart Failure 20, 853–872 (2018).

3. Go, A.S., Chertow, G.M., Fan, D.J., McCulloch, C.E. & Hsu, C.Y. Chronic kidney disease and the risks of death, cardiovascular events, and hospitalization. New England Journal of Medicine 351, 1296–1305 (2004).

4. Xie, Z.M. et al. Global Burden of the Key Components of Cardiovascular-Kidney-Metabolic Syndrome. Journal of the American Society of Nephrology 36, 1572–1584 (2025).

5. Aragam, K.G. et al. Discovery and systematic characterization of risk variants and genes for coronary artery disease in over a million participants. Nature Genetics 54(2022).

6. Liu, H.B. et al. Kidney multiome-based genetic scorecard reveals convergent coding and regulatory variants. Science 387(2025).

7. Suzuki, K. et al. Genetic drivers of heterogeneity in type 2 diabetes pathophysiology. Nature 627(2024).

8. Peterson, R.E. et al. Genome-wide Association Studies in Ancestrally Diverse Populations: Opportunities, Methods, Pitfalls, and Recommendations. Cell 179, 589–603 (2019).

9. Ren, H.H. et al. Association between atherosclerotic cardiovascular diseases risk and renal outcome in patients with type 2 diabetes mellitus. Renal Failure 43, 477–487 (2021).

10. Berger, M., Marx, N. & Marx-Schütt, K. Cardiovascular Risk Reduction in Patients with Type 2 Diabetes: What Does the Cardiologist Need to Know? European Cardiology Review 2025;20:e09. (2025).

11. Birkeland, K.I. et al. Heart failure and chronic kidney disease manifestation and mortality risk associations in type 2 diabetes: A large multinational cohort study. Diabetes Obesity & Metabolism 22, 1607–1618 (2020).

12. McIntyre, N.J., Fluck, R.J., McIntyre, C.W. & Taal, M.W. Risk Profile in Chronic Kidney Disease Stage 3: Older versus Younger Patients. Nephron Clinical Practice 119, C269–C276 (2011).

13. Harris, R.C. & Zhang, M.Z. The role of gender disparities in kidney injury. Ann Transl Med 8, 514 (2020).

14. Song, E.Y. et al. Effect of Community Characteristics on Familial Clustering of End-Stage Renal Disease. American Journal of Nephrology 30, 499–504 (2009).

15. Herrington, W.G. et al. Body-mass index and risk of advanced chronic kidney disease: Prospective analyses from a primary care cohort of 1.4 million adults in England. PLoS One 12, e0173515 (2017).

16. Lang, S.M. & Schiffl, H. Smoking status, cadmium, and chronic kidney disease. Renal Replacement Therapy 10(2024).

17. Sharma, K., Akre, S., Chakole, S. & Wanjari, M.B. Stress-Induced Diabetes: A Review. Cureus Journal of Medical Science 14(2022).

18. Lu, C.L., Li, L.Z., Chen, J.S., Chang, R.Z. & Dong, H.L. Genomic Structural Equation Modeling Reveals Cardiovascular-Kidney-Metabolic Syndrome Genetic Architecture. Journal of Diabetes 18(2026).

19. Wang, Z., Chen, X. & Wang, H. Integrative genomic dissection of cardio-kidney-metabolic syndrome reveals novel loci, causal genes, and tissue-specific pathways in European populations. Scientific Reports (2026).

20. Donker, H.C., et al. Cardiovascular-kidney-metabolic syndrome: candidate subtypes and genetic risk factors. Bmc Medical Genomics 19(2026).

21. Kurki, M.I. et al. FinnGen provides genetic insights from a well-phenotyped isolated population. Nature 613, 508–518 (2023).

22. Sudlow, C. et al. UK Biobank: An Open Access Resource for Identifying the Causes of a Wide Range of Complex Diseases of Middle and Old Age. Plos Medicine 12(2015).

23. Gaziano, J.M. et al. Million Veteran Program: A mega-biobank to study genetic influences on health and disease. Journal of Clinical Epidemiology 70, 214–223 (2016).

24. Chen, Z.M. et al. China Kadoorie Biobank of 0.5 million people: survey methods, baseline characteristics and long-term follow-up. International Journal of Epidemiology 40, 1652–1666 (2011).

25. Yang, H.C. et al. The Taiwan Precision Medicine Initiative provides a cohort for large-scale studies. Nature 648, 117–127 (2025).

26. Nagai, A. et al. Overview of the BioBank Japan Project: Study design and profile. Journal of Epidemiology 27, S2–S8 (2017).

27. Bick, A.G. et al. Genomic data in the All of Us Research Program. Nature 627(2024).

28. Kaiser, H.F. A second generation little jiffy. Psychometrika 35, 401–415 (1970).

29. Bartlett, M.S. The Effect of Standardization on a Chi-2 Approximation in Factor Analysis. Biometrika 38, 337–344 (1951).

30. Horn, J.L. A Rationale and Test for the Number of Factors in Factor-Analysis. Psychometrika 30, 179–185 (1965).

31. Brown, B.C., Ye, C.J., Price, A.L., Zaitlen, N. & Network, A.G.E. Transethnic Genetic-Correlation Estimates from Summary Statistics. American Journal of Human Genetics 99, 76–88 (2016).

32. de Leeuw, C.A., Mooij, J.M., Heskes, T. & Posthuma, D. MAGMA: Generalized Gene-Set Analysis of GWAS Data. Plos Computational Biology 11(2015).

33. Aguet, F. et al. The GTEx Consortium atlas of genetic regulatory effects across human tissues. Science 369, 1318–1330 (2020).

34. Subramanian, A. et al. Gene set enrichment analysis: A knowledge-based approach for interpreting genome-wide expression profiles. Proceedings of the National Academy of Sciences of the United States of America 102, 15545–15550 (2005).

35. Zhou, Y.Y. et al. Metascape provides a biologist-oriented resource for the analysis of systems-level datasets. Nature Communications 10(2019).

36. Christensen, A.A. & Gannon, M. The Beta Cell in Type 2 Diabetes. Current Diabetes Reports 19, 81 (2019).

37. Kwak, S.H. et al. Genetic architecture and biology of youth-onset type 2 diabetes. Nature Metabolism 6, 226–237 (2024).

38. Figueroa, K.P. & Pulst, S.M. Identification and expression of the gene for human ataxin-2-related protein on chromosome 16. Experimental Neurology 184, 669–678 (2003).

39. Ong, W.Y. et al. Neurodegeneration in Niemann-Pick type C disease mice. Experimental Brain Research 141, 218–231 (2001).

40. Li, M.W. et al. EWAS Atlas: a curated knowledgebase of epigenome-wide association studies. Nucleic Acids Research 47, D983–D988 (2019).

41. Bell, N., Uffelmann, E., Van Walree, E., de Leeuw, C. & Posthuma, D. Using Drug Gene-Set Analysis to Identify Drug Repurposing Candidates Based on Genome-Wide Association Results. European Neuropsychopharmacology 63, E105–E106 (2022).

42. Martinez Martin, F.J. Manidipine in hypertensive patients with metabolic syndrome: the MARIMBA study. Expert Rev Cardiovasc Ther 7, 863–9 (2009).

43. Bowden, J., Smith, G.D. & Burgess, S. Mendelian randomization with invalid instruments: effect estimation and bias detection through Egger regression. International Journal of Epidemiology 44, 512–525 (2015).

44. Baltramonaityte, V. et al. A multivariate genome-wide association study of psycho-cardiometabolic multimorbidity. Plos Genetics 19(2023).

45. Mallard, T.T. et al. Multivariate GWAS of psychiatric disorders and their cardinal symptoms reveal two dimensions of cross-cutting genetic liabilities. Cell Genomics 2(2022).

46. Foote, I.F. et al. Uncovering the multivariate genetic architecture of frailty with genomic structural equation modeling. Nature Genetics 57(2025).

47. Park, S. et al. Multivariate genomic analysis of 5 million people elucidates the genetic architecture of shared components of the metabolic syndrome. Nature Genetics 56, 2380-+ (2024).

48. Han, Y. et al. Genome-wide analysis identifies susceptibility loci for heart failure and nonischemic cardiomyopathy subtype in the East Asian populations. Plos Genetics 21(2025).

49. Bathina, S. & Das, U.N. Brain-derived neurotrophic factor and its clinical implications. Archives of Medical Science 11, 1164–1178 (2015).

50. Tews, D. et al. Teneurin-2 (TENM2) deficiency induces UCP1 expression in differentiating human fat cells. Molecular and Cellular Endocrinology 443, 106–113 (2017).

51. Lee, S., Lee, C.E., Elias, C.F. & Elmquist, J.K. Expression of the Diabetes-Associated Gene in Adult Mouse Brain. Journal of Comparative Neurology 517, 925–939 (2009).

52. Cousineau, J.P., Dawe, A.M. & Alpaugh, M. Investigating the Interplay between Cardiovascular and Neurodegenerative Disease. Biology-Basel 13(2024).

53. Speliotes, E.K. et al. Association analyses of 249,796 individuals reveal 18 new loci associated with body mass index. Nat Genet 42, 937–48 (2010).

54. Cannon, M.E. et al. Open Chromatin Profiling in Adipose Tissue Marks Genomic Regions with Functional Roles in Cardiometabolic Traits. G3-Genes Genomes Genetics 9, 2521–2533 (2019).

55. Li, L.L. et al. Ataxin-2: a powerful RNA-binding protein. Discover Oncology 15(2024).

56. Jiang, X.Q., Bahorik, A.L., Dintica, C.S. & Yaffe, K. Cardiovascular-kidney-metabolic syndrome and incidence of dementia among older adults. Jpad-Journal of Prevention of Alzheimers Disease 12(2025).

57. Meyre, D. et al. Genome-wide association study for early-onset and morbid adult obesity identifies three new risk loci in European populations. Nature Genetics 41, 157–159 (2009).

58. Puspasari, A. et al. Polymorphism of FTO rs1421085 as risk of metabolic syndrome; case-control study in two ethnics, Javanese and Malay population, Indonesia. Egyptian Journal of Medical Human Genetics 26(2025).

59. Wu, C.X. et al. Integrative exome sequencing and multi-omics analysis elucidates the regulatory role of rare variants in metabolic syndrome. Functional & Integrative Genomics 26(2026).

60. Zhang, Z.Y. et al. The rs1421085 variant within promotes brown fat thermogenesis. Nature Metabolism 5(2023).

61. Sullivan, A.I. et al. Mice harboring the obesity-associated SNP rs1421085 exhibit increased body weight and reveal an IRX3 neuronal circuit regulating body weight. Molecular Metabolism 100(2025).

62. SaizSatjes, M. & Martinez-Martin, F.J. Manidipine: an antihypertensive drug with positive effects on metabolic parameters and adrenergic tone in patients with diabetes. Drugs Context 7, 212509 (2018).

63. Black, C. et al. Meglitinide analogues for type 2 diabetes mellitus (Review). Cochrane Database of Systematic Reviews (2007).

64. Knop, F.K., Bronden, A. & Vilsboll, T. Exenatide: pharmacokinetics, clinical use, and future directions. Expert Opinion on Pharmacotherapy 18, 555–571 (2017).

65. Lee, M. & Rhee, M.K. Sitagliptin for Type 2 diabetes: a 2015 update. Expert Review of Cardiovascular Therapy 13, 597–610 (2015).

66. Ndumele, C.E. et al. A Synopsis of the Evidence for the Science and Clinical Management of Cardiovascular-Kidney-Metabolic (CKM) Syndrome: A Scientific Statement From the American Heart Association. Circulation 148, 1636–1664 (2023).

67. Nguyen, K. & Mitchell, B.D. A Guide to Understanding Mendelian Randomization Studies. Arthritis Care & Research 76, 1451–1460 (2024).

68. Casper, J. et al. The UCSC Genome Browser database: 2026 update. Nucleic Acids Research 54, D1331–D1335 (2026).

69. Willer, C.J., Li, Y. & Abecasis, G.R. METAL: fast and efficient meta-analysis of genomewide association scans. Bioinformatics 26, 2190–1 (2010).

70. Grotzinger, A.D. et al. Genomic structural equation modelling provides insights into the multivariate genetic architecture of complex traits. Nature Human Behaviour 3, 513–525 (2019).

71. Bulik-Sullivan, B.K. et al. LD Score regression distinguishes confounding from polygenicity in genome-wide association studies. Nature Genetics 47, 291-+ (2015).

72. Revelle, W. psych: Procedures for Psychological, Psychometric, and Personality Research. (Northwestern University, Evanston, Illinois, 2025).

73. Rosseel, Y. lavaan: An R Package for Structural Equation Modeling. Journal of Statistical Software 48, 1–36 (2012).

74. Purcell, S. et al. PLINK: A tool set for whole-genome association and population-based linkage analyses. American Journal of Human Genetics 81, 559–575 (2007).

75. Yang, J. et al. Conditional and joint multiple-SNP analysis of GWAS summary statistics identifies additional variants influencing complex traits. Nature Genetics 44, 369–U170 (2012).

76. MacArthur, J. et al. The new NHGRI-EBI Catalog of published genome-wide association studies (GWAS Catalog). Nucleic Acids Research 45, D896–D901 (2017).

77. Rentzsch, P., Witten, D., Cooper, G.M., Shendure, J. & Kircher, M. CADD: predicting the deleteriousness of variants throughout the human genome. Nucleic Acids Research 47, D886–D894 (2019).

78. Boyle, A.P. et al. Annotation of functional variation in personal genomes using RegulomeDB. Genome Research 22, 1790–1797 (2012).

79. McLaren, W. et al. The Ensembl Variant Effect Predictor. Genome Biology 17(2016).

80. Watanabe, K., Taskesen, E., van Bochoven, A. & Posthuma, D. Functional mapping and annotation of genetic associations with FUMA. Nature Communications 8(2017).

81. Guo, Y.Z. et al. SMR-Portal: an online platform for integrative analysis of GWAS and xQTL data to identify complex trait genes. Nature Methods 22, 220–222 (2025).

82. Cannon, M. et al. DGIdb 5.0: rebuilding the drug-gene interaction database for precision medicine and drug discovery platforms. Nucleic Acids Research 52, D1227–D1235 (2024).

83. Corsello, S.M. et al. The Drug Repurposing Hub: a next-generation drug library and information resource. Nature Medicine 23, 405-+ (2017).

84. Heinze, G. & Schemper, M. A solution to the problem of separation in logistic regression. Statistics in Medicine 21, 2409–2419 (2002).

85. Privé, F., Arbel, J. & Vilhjálmsson, B.J. LDpred2: better, faster, stronger. Bioinformatics 36, 5424–5431 (2020).

86. Hemani, G. et al. The MR-Base platform supports systematic causal inference across the human phenome. Elife 7(2018).

87. Wald, A. The fitting of straight lines if both variables are subject to error. Annals of Mathematical Statistics 11, 284–300 (1940).

88. Bowden, J., Smith, G.D., Haycock, P.C. & Burgess, S. Consistent Estimation in Mendelian Randomization with Some Invalid Instruments Using a Weighted Median Estimator. Genetic Epidemiology 40, 304–314 (2016).

89. Burgess, S., Butterworth, A. & Thompson, S.G. Mendelian Randomization Analysis With Multiple Genetic Variants Using Summarized Data. Genetic Epidemiology 37, 658–665 (2013).

90. Burgess, S. & Thompson, S.G. Interpreting findings from Mendelian randomization using the MR-Egger method. European Journal of Epidemiology 32, 377–389 (2017).

91. Verbanck, M., Chen, C.Y., Neale, B. & Do, R. Detection of widespread horizontal pleiotropy in causal relationships inferred from Mendelian randomization between complex traits and diseases (vol 50, 693, 2018). Nature Genetics 50, 1196–1196 (2018).

92. Kim, M.S. et al. Causal effect of adiposity on the risk of 19 gastrointestinal diseases: a Mendelian randomization study. Obesity 31, 1436–1444 (2023).

93. Welfare, A.I.o.H.a. Comorbidity of cardiovascular disease, diabetes and chronic kidney disease in Australia. in Heart, stroke & vascular diseases (2007).

94. Usman, M.S., Khan, M.S. & Butler, J. The Interplay Between Diabetes, Cardiovascular Disease, and Kidney Disease. in Chronic Kidney Disease and Type 2 Diabetes 13-8 (Arlington (VA), 2021).

95. Koye, D.N., Magliano, D.J., Nelson, R.G. & Pavkov, M.E. The Global Epidemiology of Diabetes and Kidney Disease. Adv Chronic Kidney Dis 25, 121–132 (2018).

96. Sharma, S. et al. Genetic ancestry and population structure in the All of Us Research Program cohort. Nature Communications 16(2025).

97. Auwera, G.v.d. & O’Connor, B.D. Genomics in the cloud: using Docker, GATK, and WDL in Terra, (O’Reilly Media, Sebastopol, CA, 2020).

98. Murtagh, F. & Legendre, P. Ward’s Hierarchical Agglomerative Clustering Method: Which Algorithms Implement Ward’s Criterion? Journal of Classification 31, 274–295 (2014).

99. Mudge, J.M. et al. GENCODE 2025: reference gene annotation for human and mouse. Nucleic Acids Research 53, D966–D975 (2025).

100. M, C. org.Hs.eg.db: Genome wide annotation for Human. R package version 3.8.2 (2019).

101. Holman, R.R. et al. Effects of Once-Weekly Exenatide on Cardiovascular Outcomes in Type 2 Diabetes. New England Journal of Medicine 377, 1228–1239 (2017).

102. Bridges A, B.K., Jacobs TF. Exenatide. StatPearls Publishing (2023).

103. Rizzo, M.R., Barbieri, M., Grella, R., Passariello, N. & Paolisso, G. Repaglinide has more beneficial effect on cardiovascular risk factors than glimepiride: data from meal-test study. Diabetes & Metabolism 31, 255–260 (2005).

104. Milner Z, A.H.R. Repaglinide. StatPearls Publishing (2023).

105. Olowofeso AM, M.F., Zinkevich NS. Sitagliptin in Type 2 Diabetes Mellitus and Cardiovascular Disease: A Public Health and Health Equity Perspective. Cardiovascular Science 2, 10010 (2025).

106. Eligar, V.S. & Bain, S.C. A review of sitagliptin with special emphasis on its use in moderate to severe renal impairment. Drug Design Development and Therapy 7, 893–903 (2013).

